# Generalizable AI predicts immunotherapy outcomes across cancers and treatments

**DOI:** 10.1101/2025.05.01.25326820

**Authors:** Wanxiang Shen, Intae Moon, Thinh H. Nguyen, Michelle M. Li, Yepeng Huang, Nitya Nair, Daniel Marbach, Marinka Zitnik

## Abstract

Immune checkpoint inhibitors are standard across cancers, yet most patients do not respond and existing biomarkers generalize poorly across tumor types, drugs and clinical settings. We present Compass, a pan-cancer foundation model that predicts immunotherapy response from bulk tumor transcriptomes using a concept-bottleneck transformer. Compass encodes gene expression through 44 biologically grounded immune concepts representing immune cell states, tumor-microenvironment interactions, and signaling pathways. Trained on 10,184 tumors across 33 cancer types, Compass outperforms 22 baseline methods in 16 independent clinical cohorts spanning seven cancers and six immune checkpoint inhibitors, increasing accuracy by 8.5% and area under the precision-recall curve by 15.7%, with minimal additional training. The model generalizes to unseen cancer types and treatments, supporting indication selection and patient stratification in early-phase clinical trials. In survival analyses, Compass-stratified responders have longer overall survival (hazard ratio = 4.7, *p <* 0.0001). Personalized response maps connect gene expression to immune concepts, revealing mechanisms of response and resistance; in immune-inflamed non-responders, Compass highlights programs including TGF-*β* signaling, endothelial exclusion, CD4+ T cell dysfunction, and B cell deficiency. By combining interpretability with transfer learning, Compass enables robust prediction and mechanistic insight to inform trial design and translational studies.

## Main

Immune checkpoint inhibitors (ICIs) have transformed cancer treatment, but clinical benefit remains uneven across tumor types, and only a minority of patients achieve durable responses^1^. While tumor mutational burden (TMB) and PD-L1 expression represent clinically validated predictive biomarkers, their limited accuracy restricts reliable patient selection^2^. Responses range from durable remission to primary resistance, reflecting differences in tumor–immune interactions. In solid tumors, ICI responders typically exhibit an immune-inflamed phenotype, marked by CD8+ T cell infiltration. Non-response is often associated with immune-desert or immune-excluded phenotypes^3^. A substantial subset of non-responders nevertheless maintains immune-inflamed characteristics, highlighting the biological complexity of resistance mechanisms. Advancing predictive capacity for ICI and delineating response and resistance mechanisms are critical for optimizing personalized treatments and improving patient outcomes^4^.

Biomarkers, including TMB, PD-L1 IHC score, CD8+ T cell infiltration, and immune gene expression signatures, provide incomplete insights into ICI response mechanisms^5^. High TMB correlates with clinical benefit to ICI in some cancers, likely as a surrogate for increased neoantigen presentation^6^, but fails to fully predict response: many high-TMB tumors remain refractory, while some low-TMB tumors respond robustly. Transcriptomic signatures, such as TIDE (T cell dysfunction)^7^ and IMPRES (immune checkpoint activity)^8^, offer mechanistic insight but show variable predictive performance across cancer types^9–11^. A pan-cancer analysis of 27,810 ICI-treated patients further underscores these limitations, revealing weak or inconsistent associations between PD-L1, CD8+ T cells, immune gene scores, and TMB with response across tumors^12^. Transcriptome-based machine learning models use gene expression and related omics signals to predict treatment response. ENLIGHT models response using transcriptome-derived gene interactions^13–15^. EaSIeR predicts response from interpretable immune and microenvironmental gene programs^16^. Graph-based approaches represent molecular dependencies as graphs and learn predictors over these structures^17^. Although valuable, many methods remain cancer-type specific or rely on fixed interaction maps and predefined gene-signature scores.

We developed Compass, a pan-cancer foundation model that learns interpretable tumor-immune concepts from transcriptomic data. Compass uses a concept bottleneck architecture^18^ that routes a patient’s transcriptome through human-readable concepts. In retinal imaging, for example, such models can first predict tissue and lesion maps and then use these intermediate outputs for diagnostic triage. We introduce this approach in cancer transcriptomics by defining tumor-immune concepts, pretraining with self-supervised contrastive learning on pan-cancer RNA-seq, and transferring to small clinical cohorts with parameter-efficient fine-tuning. Compass maps bulk RNA-seq profiles to biologically grounded concepts spanning immune cell states, tumor–microenvironment interactions, and signaling pathways (**Fig. 1a**). It pretrains on transcriptomes from 33 cancer types and fine-tunes on clinical cohorts to predict response to immune checkpoint inhibitors, including anti-PD1/PD-L1, anti-CTLA4, and combination therapies (**Fig. 1b, c**).

**Fig. 1:**
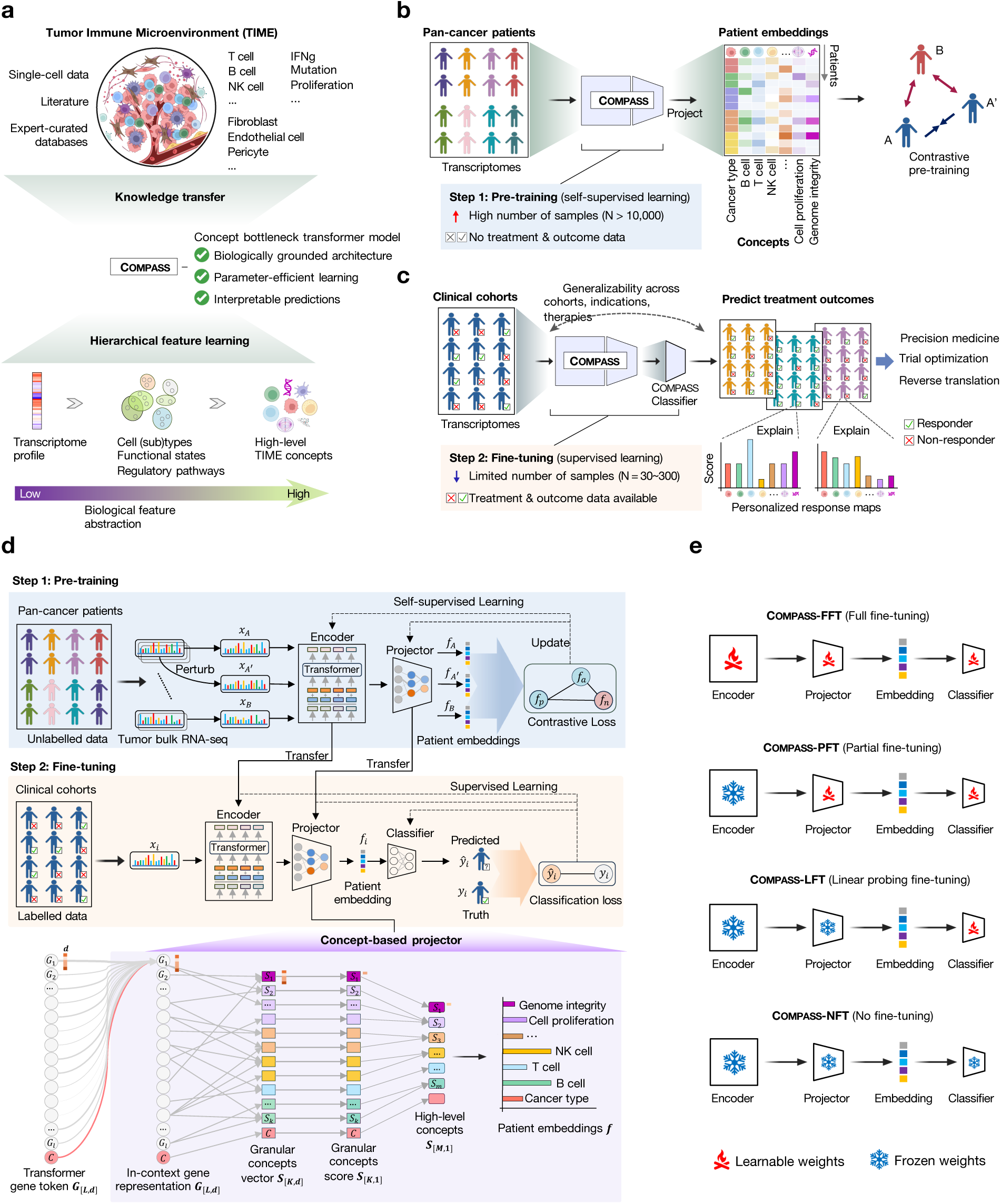
Concept-bottleneck foundation model for interpretable prediction of immunotherapy response. **(a) Transfer of immuno-oncology knowledge via hierarchical concept learning.** The Compass concept-bottleneck architecture integrates tumor immune microenvironment (TIME) gene signatures capturing cell types, functional states, and signaling pathways (top). Hierarchical feature learning transforms expression profiles into interpretable TIME representations through successive abstraction levels (bottom). This biologically grounded, parameter-efficient architecture enables generalizable and interpretable predictions of immunotherapy outcomes. **(b) Self-supervised pre-training on pan-cancer transcriptomes.** Using contrastive learning, Compass projects bulk RNA-seq profiles into biologically grounded patient embeddings that capture essential cell-type and functional pathway information. Triplet training aligns perturbed (augmented) versions of each tumor (A, A’) while distinguishing unrelated samples (B) in concept space. By leveraging large-scale, unlabeled pan-cancer cohorts, this process builds generalizable TIME representations that form the foundation for predictive tasks. **(c) Fine-tuning on clinical cohorts for explainable prediction of immunotherapy outcomes.** The pre-trained model is fine-tuned on clinical cohorts using supervised learning, where a lightweight classifier is added to predict immune checkpoint inhibitor (ICI) patient responses. Through parameter-efficient transfer learning, Compass adapts to diverse treatment regimens and patient populations without overfitting, generating interpretable predictions grounded in the learned TIME concepts. **(d) Architecture of the Compass model.** The model comprises three components: (1) a transformer-based gene encoder that transforms expression profiles into context-aware representations, (2) a hierarchical concept projector that progressively aggregates these into multi-scale TIME concepts, and (3) a task-specific classifier that outputs predictions from concept-level features. The projector’s concept-bottleneck architecture (purple box) first maps gene embeddings to granular TIME concepts, then compresses these into higher-level representations. The classifier leverages these biologically grounded features to distinguish responders from non-responders. During pre-training (blue box), triplet contrastive learning updates the encoder and projector to construct discriminative TIME representations. This is achieved through contrastive loss minimization, which reduces the cosine distance between perturbed (augmented) views of the same tumor (*f_A_*, *f_A_′* ) while increasing their separation from other samples (*f_B_*) in the concept space. Fine-tuning (yellow box) employs supervised learning on clinical cohorts, selectively adapting components (encoder, projector, or classifier) while preserving the interpretable concept hierarchy. **(e) Flexible fine-tuning strategies for clinical adaptation.** Four transfer learning modes balance stability with adaptation to new clinical cohorts of varying sizes: (1) **Full fine-tuning (Compass-FFT)**: Updates all components (encoder, projector, classifier), refining model weights for task-specific alignment. (2) **Partial fine-tuning (Compass-PFT)**: Adjusts only projector and classifier, preserving encoder knowledge while recalibrating TIME concepts. (3) **Linear probing (Compass-LFT)**: Updates classifier alone, leveraging frozen pre-trained TIME concepts for small cohorts. (4) **No fine-tuning (Compass-NFT)**: Uses cosine similarity to reference patients with known response labels in pre-trained TIME-space; **Supplementary Fig. S1**). Trainable components are denoted by fire icons (learnable weights) versus ice icons (frozen weights).

We evaluate Compass on 1,133 patients from 16 clinical cohorts spanning seven cancer types, using pre-treatment tumor RNA-seq profiles to predict response to ICIs. In leave-one-cohort-out evaluation, Compass outperforms 22 existing models, achieving 8.5% higher accuracy and 15.7% greater area under the precision-recall curve. Across settings, Compass generalizes to new cohorts, cancer types, and treatments, and supports parameter-efficient transfer to small clinical cohorts. In a held-out phase II trial of metastatic urothelial cancer, patients predicted as responders by Compass show significantly longer survival (HR = 4.7, *p* = 1.7 × 10^−7^), outperforming TMB and PD-L1 immunohistochemistry biomarkers.

Compass generates personalized response maps that connect gene expression to immune concepts and provides mechanistic interpretation of treatment response for individual patients. The maps highlight key drivers (T cell exhaustion, myeloid programs, immunoregulatory signaling) and resolve cases where canonical immune phenotypes (inflamed, excluded, desert) are misleading. For example, inflamed non-responders exhibit TGF-*β*-driven suppression, vascular exclusion, or CD4+/B-cell dysfunction, while non-inflamed responders display residual cytotoxic activity or TMB-associated pathways without immunosuppressive signals. These results show that Compass identifies functional immune states that complement the coarse stratification provided by PD-L1 expression, TMB, or bulk immune phenotypes. By combining transcriptomic data with tumor-immune modeling, Compass achieves cross-cancer and cross-therapy prediction and yields mechanistic insight into determinants of treatment response.

## Results

### Compass concept bottleneck transformer for immunotherapy response prediction

Predicting response to immunotherapy remains difficult due to small cohort size, patient heterogeneity, and complexity of tumor-immune interactions. To address this, we developed Compass, a concept bottleneck-based model that combines pan-cancer pre-training on bulk tumor transcriptomes with a biologically structured architecture grounded in immune and stromal concepts (**Fig. 1a**). This design improves generalizability and supports parameter-efficient, interpretable adaptation to clinical settings.

To ground the model in biologically meaningful features, we curated 132 gene signatures from the literature (**Supplementary Data S1**). The signatures capture immune cell types, functional states, pathways, and non-immune tumor biology (e.g., stromal programs and DNA damage response), which define Compass’s interpretable representation of tumor-immune dynamics.

Compass uses a transformer-based gene language model to encode expression profiles of 15,672 protein-coding genes (**Fig. 1d**, **Methods 2**). It projects gene embeddings onto 132 gene signatures, which are then aggregated into 43 high-level TIME concepts using a hierarchical projector. A cancer-type token is included as an additional concept. This process generates 44-dimensional patient embeddings that reflect patient-specific transcriptional programs.

We pre-trained Compass on transcriptomes from 10,184 tumors in The Cancer Genome Atlas (TCGA) using self-supervised contrastive learning (**Methods 3**). Contrastive learning pulls perturbed (augmented) views of the same tumor together and pushes different tumors apart in the 44-dimensional concept space (**Fig. 1d**). This approach allows the model to learn patient representations of TIME. After pre-training, Compass is fine-tuned on clinical cohorts to predict immunotherapy response (**Methods 4**). A classifier adaptor takes the 44-dimensional concept vector as input and outputs a response probability, with parameter-efficient updates depending on cohort size (**Fig. 1d, e**): full fine-tuning for large cohorts, partial updates for medium cohorts, and adaptor-only training for small cohorts; when outcomes are unavailable, Compass supports zero-shot prediction via similarity in concept space (**Supplementary Fig. S1**).

### Compass achieves state-of-the-art performance across 16 immunotherapy cohorts

We consider pre-treatment bulk RNA-seq and clinical outcomes for 1,133 patients from 16 clinical cohorts (**Fig. 2a, Extended Data Table 1**, **Supplementary Table S1**). These cohorts span seven cancer types and multiple ICI regimens (anti-PD1/PD-L1, anti-CTLA4, and combinations), with cohort sizes ranging from 16 to 298 patients. Patients are classified as responders (*n* = 346, 30.5%; complete or partial remission) or non-responders (*n* = 787, 69.5%; progressive or stable disease) (**Methods 1.2**). This dataset captures heterogeneity across indications, treatments, platforms, and study designs.

**Fig. 2:**
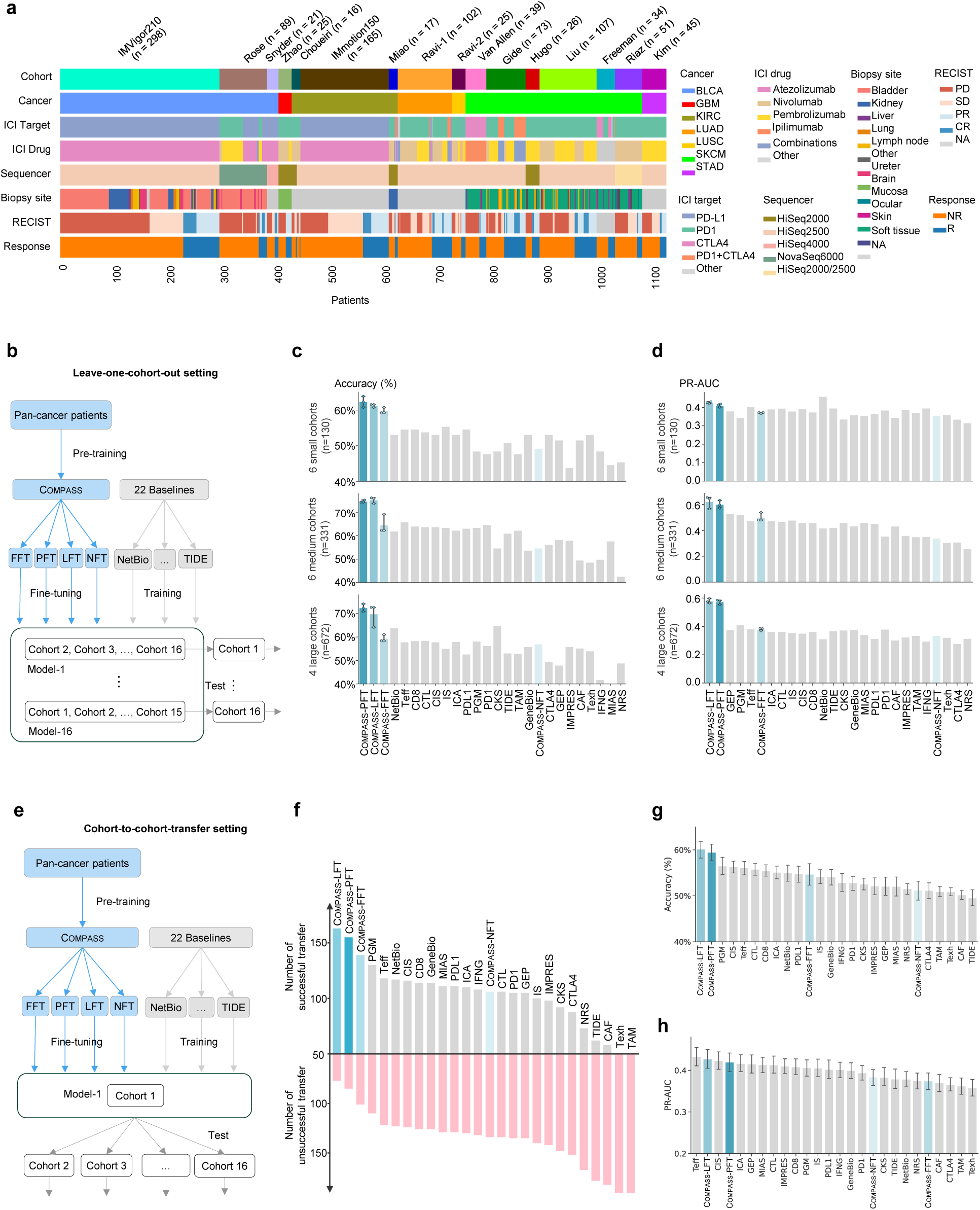
Evaluation of Compass across clinical cohorts for immunotherapy response prediction. **(a) Overview of clinical cohorts.** Each column represents one patient. The dataset is diverse in cohort size, patient indication, drug treatment, and response outcomes. It includes 16 cohorts across seven cancer types, categorized by size into large (four cohorts, *n >* 100: IMvigor210^22^, IMmotion150^35^, Ravi-1 (SU2C-MARK LUAD cohort)^40^, and Liu^43^), medium (six cohorts, *n* = 30 to 100: Freeman (MGH cohort)^47^, Van Allen^45^, Kim^39^, Riaz^6^, Gide^44^, and Rose^42^), and small (six cohorts, *n <* 30: Choueiri^36^, Miao^38^, Snyder^41^, Zhao^37^, Ravi-2 (SU2C-MARK LUSC cohort)^40^, and Hugo^46^) (**Extended Data Table 1, Supplementary Table S1**). All RNA-seq data originate from pre-treatment samples, with variability in biopsy sites, sequencing platforms, and analytical pipelines. This heterogeneity enables rigorous benchmarking of Compass and baseline methods. **(b-d) Leave-one-cohort-out evaluation.** Models are trained on 15 cohorts and tested on the held-out cohort. This process is iterated across all cohorts for 22 baseline methods (**Supplementary Table S2**) and Compass ’s four fine-tuning modes. Performance is aggregated by cohort size groups (large/medium/small) to improve statistical power. Panel **(c)** presents accuracy (%), and panel **(d)** shows PR-AUC. For Compass’s fine-tuning modes, experiments are repeated three times with different random seeds; error bars show standard deviations. The numerical results are provided in **Supplementary Table S3**. **(e-h) Cohort-to-cohort-transfer evaluation.** Models initially trained on one cohort are tested on the remaining 15 cohorts to evaluate generalization across different populations, indications, and treatment regimens. A cohort-to-cohort transfer is considered successful when accuracy is greater than the baseline accuracy for the held-out target cohort (**Method 5**). The overall success rate across 240 pairwise transfers (16×15) serves as an indicator of model robustness (panel **f**). Panels **(g)** and **(h)** show average accuracy and PR-AUC, respectively, aggregated across the 240 independent pairwise transfers. Bars show the mean performance, and error bars represent the 95% confidence interval. PD1/PDL1/CTLA4: drug target markers; NetBio^48^: Network-Based ICI Treatment Biomarkers; PGM^47^: Paired Gene Markers; GeneBio^48^: combination of PD1, PDL1, and CTLA4; CIS^49^: Cytotoxic Immune Signature; Teff^50^: T-effector-IFNg Signature; NRS^51^: Neoadjuvant Response Signature; IFNG^9^: IFNG Signature Score; CTL^7^: Cytotoxic T Lymphocytes Markers; TAM^7, 52^: Tumor-Associated Macrophages; Texh^7^: T-cell Exhaustion; CKS^53^: Chemokine Signature Score; CAF^54^: Cancer-Associated Fibroblasts Signature Score; IS^55^: Roh Immune Score; ICA^56^: Immune Cytolytic Activity Score; CD8^48^: CD8 Signature Score (CD8A + CD8B); MIAS^10^: MHC I Association Immune Score; GEP^10, 11^: the T Cell-Inflamed Gene Expression Profile Score (GEP); IMPRES^8^: the Immuno-Predictive Score; TIDE^7^: Tumor Immune Dysfunction and Exclusion Score.

To benchmark predictive performance, we compared Compass against 22 widely used ICI response prediction methods, spanning single-gene predictors, immune signature scoring approaches, and network/ML models (**Supplementary Table S2**). We used a leave-one-cohort-out evaluation strategy to assess generalizability in clinically relevant scenarios (**Fig. 2b**). Models were trained on all but one cohort and evaluated on the held-out cohort. We evaluated performance using accuracy, area under the precision-recall curve, area under the ROC curve, and Matthews correlation coefficient (MCC) (**Methods 5**).

Compass substantially improved upon established methods across all cohort sizes and metrics (**Fig. 2c-d**, **Supplementary Fig. S2**, **Supplementary Table S3**). Partial fine-tuning (Compass-PFT) and linear probing (Compass-LFT) consistently delivered the highest overall predictive performance. Compared to the second-best-performing models, Compass-PFT and Compass-LFT improved accuracy by an average of 8.5%, AUPRC by 15.7%, and MCC by 12.3%, demonstrating their superior capability for response prediction. Performance remained robust across large, medium, and small cohorts, with Compass-PFT and Compass-LFT showing consistently strong results across settings (**Fig. 2c-d**).

Among the fine-tuning strategies evaluated, partial fine-tuning (Compass-PFT) and linear probing (Compass-LFT) provided the best balance between model adaptability and stability. In contrast, full fine-tuning (Compass-FFT), which updates all model parameters during training, performed worse when large cohorts were held out during leave-one-cohort-out evaluation, likely due to overfitting. The biologically informed architecture and parameter-efficient fine-tuning of Compass thus offer a practical approach to capture clinically relevant tumor-immune interactions for robust and generalizable predictions of immunotherapy outcomes.

### Compass generalizes within and across immunotherapy cohorts

We next assessed how accurately Compass predicts immunotherapy responses when training data are limited to a single cohort. This mimics early-phase settings where models are trained on one study. We used two benchmarking strategies: intra-cohort leave-one-patient-out validation and cohort-to-cohort transfer across studies.

In intra-cohort validation, Compass consistently outperformed previously developed methods across all cohort sizes (**Supplementary Figs. S3-S6**). Partial fine-tuning (Compass-PFT), linear probing (Compass-LFT), and full fine-tuning (Compass-FFT) achieved the highest accuracy on medium and large cohorts. For small cohorts (fewer than 30 patients), the no fine-tuning variant (Compass-NFT) performed best (**Supplementary Fig. S3b**). This likely reflects limited data, where additional fine-tuning can overfit and Compass-NFT leverages the pre-trained concept space. Among previously published approaches, NetBio performed best on medium and large cohorts. For small cohorts, PGM, Texh, and TIDE demonstrated better accuracy.

In the cohort-to-cohort transfer analyses, models trained on one cohort were used to predict responses in a different cohort, resulting in 240 pairwise transfer evaluations (16 cohorts × 15 possible transfers per cohort; **Fig. 2e-f**, **Supplementary Figs. S7-S8**). We defined successful transfer as achieving a prediction accuracy greater than the reference accuracy for the target cohort (**Methods 5**). Linear probing (Compass-LFT) performed best, with successful transfer in 163 of 240 cases, followed closely by partial fine-tuning (Compass-PFT) with 155 successful transfers. These results substantially exceeded the top-performing previously published methods, including PGM (130/240), Teff (118/240), and NetBio (117/240), with aggregated prediction accuracy shown in (**Fig. 2g**). The strong performance of Compass-LFT, which updates only the final classification layer, suggests improved robustness under limited training data. We also evaluated area under the precision-recall curve (PR-AUC), a metric that is informative in imbalanced settings because it focuses on positive-class retrieval, and observed results consistent with the other performance measures (**Fig. 2h**).

### Compass predicts treatment response across cancer types, treatments, and drug targets

To be clinically useful, predictive models must generalize beyond individual studies across cancer types, treatment regimens, and immune checkpoint targets despite biological and technical heterogeneity^19, 20^. We evaluated Compass’s generalizability along these clinically important axes by stratifying patients by cancer type, therapy, and immune checkpoint targets (**Fig. 3**, **Supplementary Fig. S9**).

**Fig. 3:**
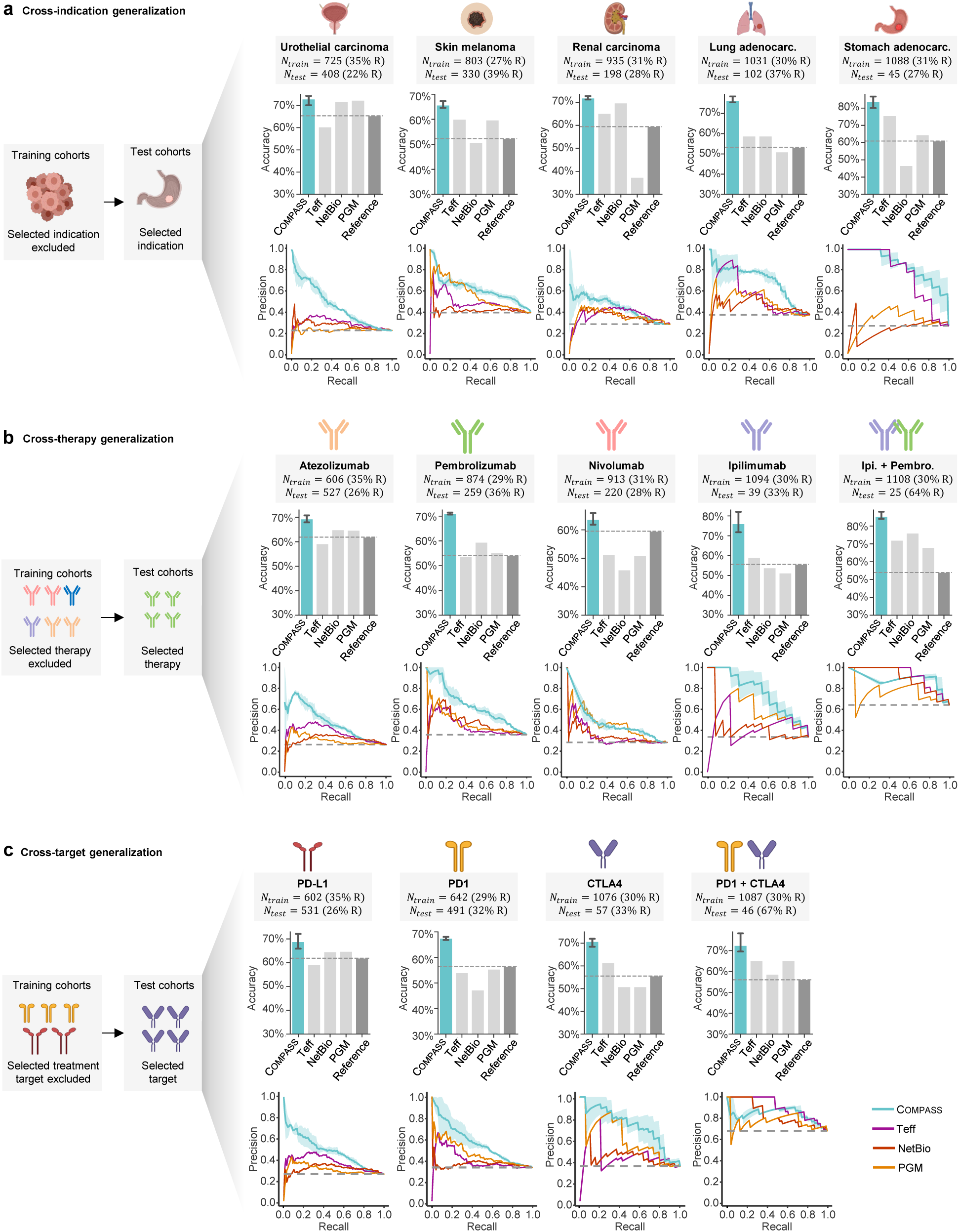
Evaluation of model generalization across biological contexts. To assess the generalizability of the Compass-PFT model relative to best-performing methods (Teff, NetBio, and PGM), we evaluate the models under biologically meaningful data splits stratified by indication, drug, or ICI target. In the figure, *t* denotes the number of training samples, n represents the number of test samples, with the percentage of responders in each set annotated. For each evaluation, the models are trained on cohorts excluding a specific category and tested on the excluded cohort. Top panels show accuracy (%), bottom panels precision-recall curves labeled with average precision (AU-PRC). (a) **Cross-disease generalization.** Models are trained on ICI cohorts excluding one specific indication and then tested on the excluded indication. For example, the stomach adenocarcinoma cohorts are excluded during training and used for testing. The results show accuracy and precision-recall curves for urothelial carcinoma, skin melanoma, renal carcinoma, lung adenocarcinoma, and stomach adenocarcinoma (**Supplementary Table S4**). (b) **Cross-therapy generalization.** Models are trained on cohorts excluding those treated with a specific drug and then tested on the excluded therapy. For instance, the pembrolizumab treatment cohorts are excluded during training and used for testing. The results include accuracy and precision-recall curves for atezolizumab, pembrolizumab, nivolumab, ipilimumab, and a combination of ipilimumab and pembrolizumab (**Supplementary Table S5**). (c) **Cross-target generalization.** Models are trained on cohorts excluding those targeting a specific immune checkpoint and then tested on the excluded target. For example, the CTLA4 treatment cohort is excluded during training and used for testing. The results cover accuracy and precision-recall curves for PDL1, PD1, CTLA4, and a combination of PD1 and CTLA4 (**Supplementary Table S6**).

We compared partial fine-tuning (Compass-PFT) against top-performing baselines (PGM, NetBio, Teff) and found consistently higher accuracy and precision–recall performance across settings. In cross-indication prediction (**Fig. 3a**), a task critical for indication selection in clinical drug development, Compass-PFT identified responders in cancer types excluded from training. For example, when lung adenocarcinoma was excluded from training, Compass-PFT achieved 76.5% accuracy on the held-out cohort (**Supplementary Table S4**). These results demonstrate that Compass can distinguish generalizable features of antitumor immunity from indication-specific transcriptional variation.

In cross-therapy and cross-target evaluations (**Fig. 3b-c**), Compass-PFT generalized across regimens and checkpoint targets (70.8% accuracy for anti-CTLA4 when trained on PD1/PD-L1 cohorts; **Supplementary Table S5**), indicating shared immune mechanisms. The cancer-type token ablation studies on cross-indication and cross-target generalization show that the performance is not driven by type-specific prevalence of response (**Supplementary Fig. S10**). Another clinically relevant problem is predicting responses to combination therapies (ipilimumab plus pembrolizumab) using models trained on non-combination cohorts. Here, Compass-PFT achieved 85.3% accuracy when trained only on monotherapy cohorts (**Fig. 3b**, **Supplementary Fig. S11**).

We also tested generalizability across technical factors, including sequencing platforms and biopsy sites (**Supplementary Fig. S12**). Compass-PFT remained robust, whereas signature-based models were sensitive to platform and site differences.

### Multi-stage fine-tuning improves prediction for new therapies and cancer types

In clinical development, early-stage studies provide limited target-specific data, making indication selection and patient enrichment challenging. To address this, Compass uses a multi-stage fine-tuning (MSFT) strategy to build robust, treatment-specific models from small clinical cohorts (**Fig. 4a**). MSFT sequentially (i) pre-trains on large transcriptomic data, (ii) fine-tunes on pan-cancer ICI cohorts, and (iii) refines on the target drug or combination, transferring shared immune-response features to therapy-specific settings.

**Fig. 4:**
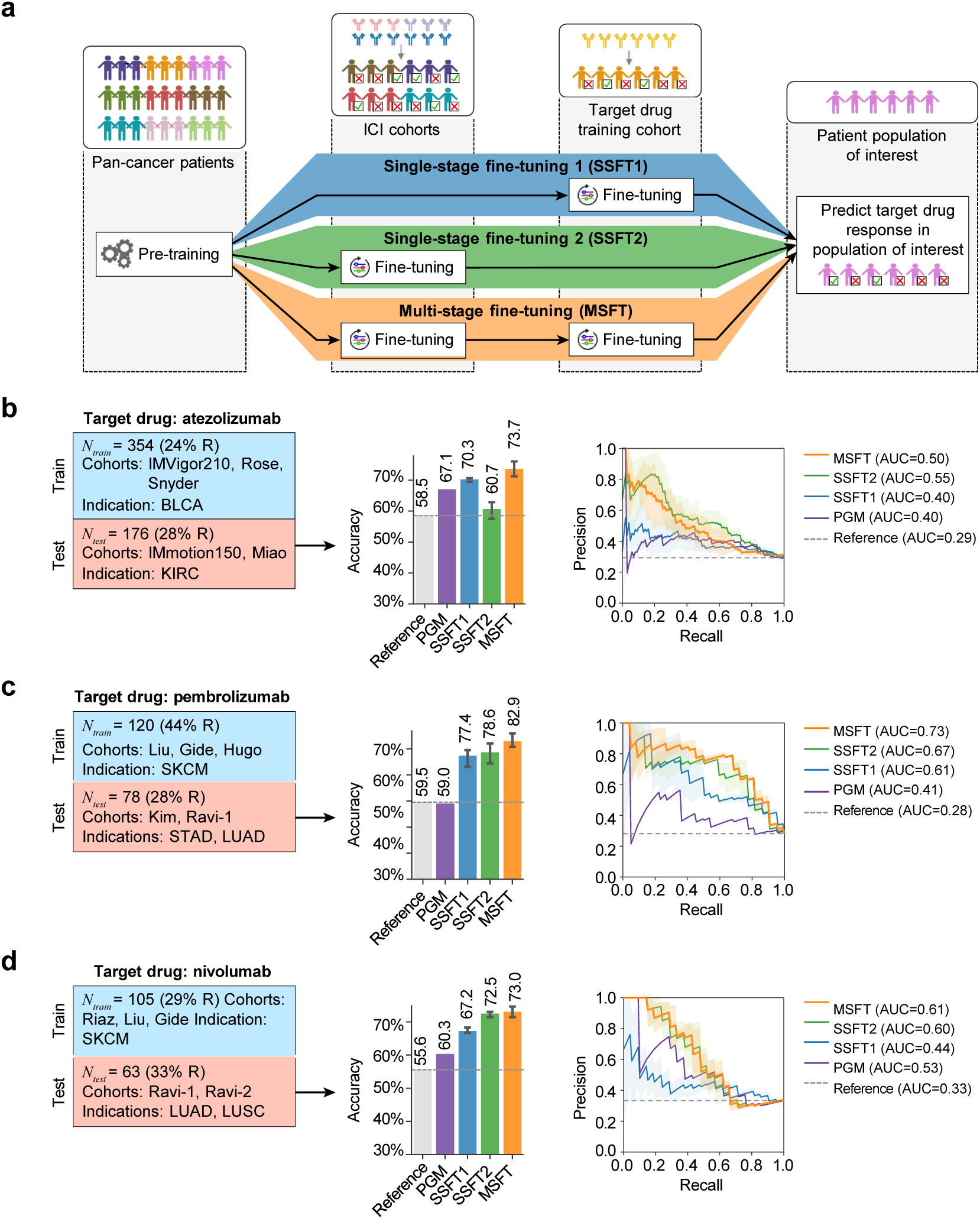
Development and evaluation of drug-specific models using single-stage and multi-stage fine-tuning strategies. **(a) Fine-tuning strategies based on the pre-trained Compass model.** Drug-specific models are developed using three strategies: single-stage fine-tuning 1 (SSFT1), which fine-tunes directly on the drug-specific cohort; single-stage fine-tuning 2 (SSFT2), which fine-tunes on general ICI-treated cohorts; and multi-stage fine-tuning (MSFT), which sequentially fine-tunes on both (i.e., general then drug-specific cohorts). PGM and reference baseline models are trained using only the drug-specific cohorts. Patient cohorts are mutually exclusive between SSFT1 and SSFT2 stages. **(b–d) Model performance across three drug settings.** Drug-specific models were developed and evaluated for: (b) atezolizumab (anti-PD-L1), (c) pembrolizumab (anti-PD-1), and (d) nivolumab (anti–PD-1). Each subplot summarizes training and test cohort composition, including sample sizes (*N*_train_ and *N*_test_) and responder proportions (**Supplementary Table S7**). Training schemes differ by fine-tuning strategy: SSFT1 uses only drug-specific cohorts (e.g., bladder cancer for atezolizumab); SSFT2 uses broader ICI cohorts, excluding treatments that target the same checkpoint pathway; MSFT combines both. All models are evaluated on held-out cancer types. Bar plots present accuracy, and line plots show precision–recall curves along with the corresponding area under the curve (AUC) values. MSFT consistently outperforms SSFT1, SSFT2, PGM, and reference baselines. Complete results in **Supplementary Table S8**.

We evaluated MSFT by developing drug-specific models for three immune checkpoint inhibitors: atezolizumab (anti-PD-L1), pembrolizumab, and nivolumab (both anti-PD1) (**Methods 6**). Models were pre-trained, fine-tuned on ICI cohorts excluding the target drug/class, and refined on a single target-drug cohort, then tested on held-out cohorts treated with the same drug (**Supplementary Table S7**).

MSFT outperformed single-stage fine-tuning strategies and existing drug-specific models (**Fig. 4b-d**). In predicting atezolizumab response in kidney cancer (KIRC, *n* = 89), MSFT achieved 73.7% accuracy, compared to 70.3% for single-stage fine-tuning on the drug-specific cohort alone (SSFT1) and 60.7% when fine-tuning models using only pan-cancer data (SSFT2).

We also applied MSFT to disease-specific prediction tasks (**Methods 6**). For lung adenocarcinoma (LUAD), we predicted pembrolizumab responses (*n* = 33) (**Supplementary Fig. S13**). MSFT reached 91% accuracy. In contrast, SSFT1 trained only on the LUAD cohort achieved 67% accuracy. Thus, MSFT improves prediction in small treatment- or disease-specific settings.

### Compass predicts survival and reveals resistance mechanisms beyond immune phenotypes

We characterized the 44 interpretable TIME concepts learned by Compass, including cross-cohort stability, associations with response, and comparisons to fixed gene-set scoring approaches (**Fig. 5a; Supplementary Figs. S14–S19, Extended Data Fig. 1; Extended Data Fig. 2; and Extended Data Fig. 3**). We next evaluated the clinical relevance of these concepts by applying Compass to IMvigor210, a held-out phase II trial of atezolizumab (anti-PD-L1) in metastatic urothelial cancer^21, 22^, to assess whether it predicts long-term outcomes and reveals resistance mechanisms. To prevent data leakage, we used a Compass-PFT model fine-tuned on all other ICI-treated cohorts, excluding IMvigor210.

**Fig. 5:**
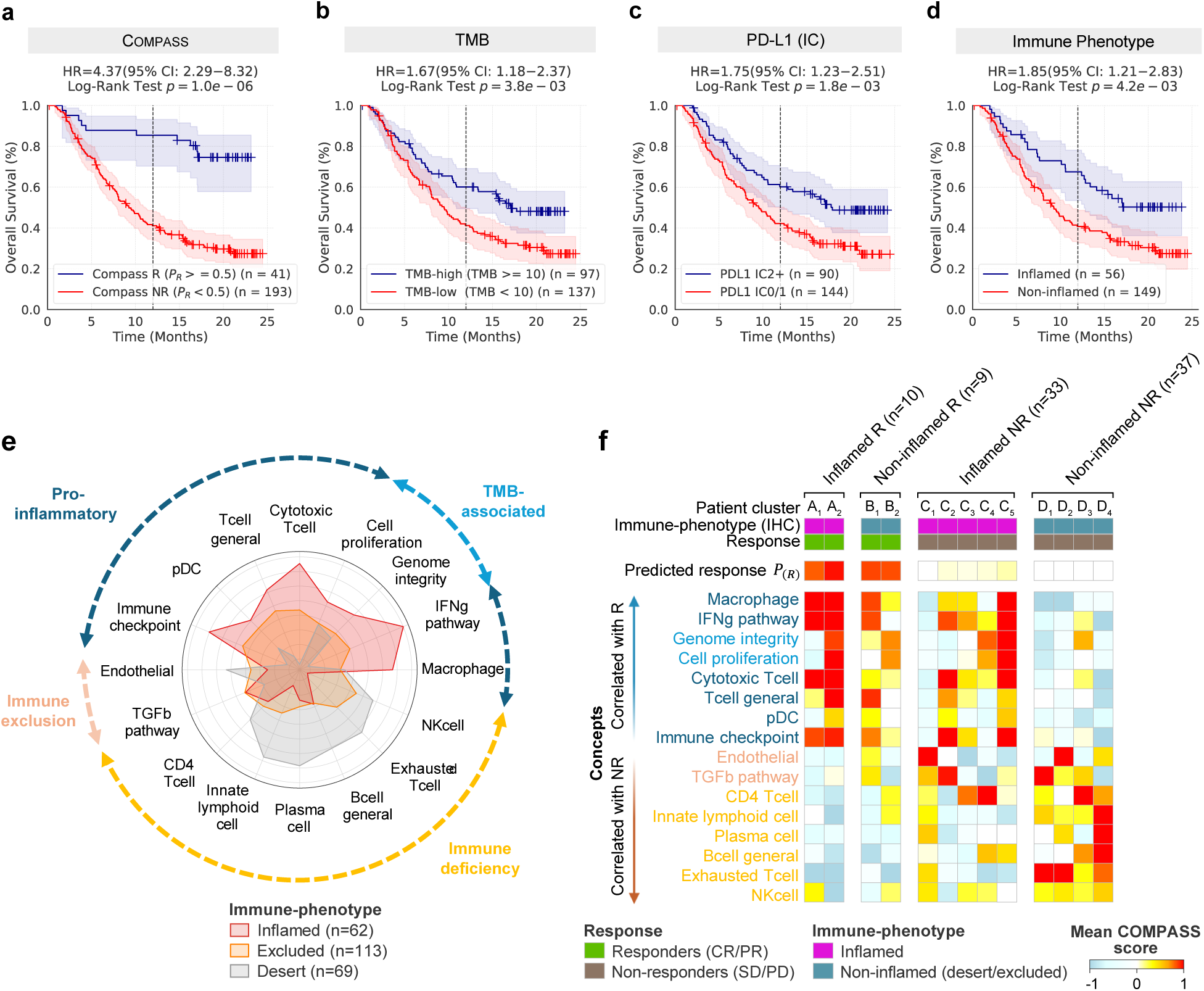
Survival prediction and tumor-immune profiling in atezolizumab-treated urothelial carcinoma. Compass evaluation of atezolizumab-treated urothelial carcinoma patients (IMvigor210 cohort), with partial fine-tuning on external ICI-treated cohorts to prevent data leakage. **(a–d) Kaplan–Meier survival analysis.** Overall survival stratified by: (a) Compass-predicted response probability (*P_R_* ≥ 0.5 vs *P_R_* < 0.5), (b) tumor mutation burden (TMB-high [≥10 mut/Mb] vs TMB-low), (c) PD-L1 immune cell score (IC2+ vs IC0/1), and (d) immune phenotype (inflamed vs excluded/desert). Analysis restricted to patients with available TMB data (n=234). Compass-predicted responders showed superior survival benefit (HR = 4.37, 95% CI: 2.29–8.32, *p* = 1.0 ×10*^−^*^6^) compared to conventional biomarkers. **(e) Immune phenotype-specific concept profiles**. Radar plot displays average scores of 16 key Compass-derived TIME concepts across immune phenotypes: inflamed (red, *n* = 62), excluded (orange, *n* = 113), and desert (beige, *n* = 69). Concepts are grouped by functional category (**Method 9**): pro-inflammatory (dark blue), TMB-associated (blue), immune exclusion (peach), and immune deficiency (yellow). Inflamed tumors show elevated pro-inflammatory and TMB-associated concept scores, while excluded/desert phenotypes exhibit immunosuppressive features. **(f) Concept score patterns across immune phenotype and response subgroups.** Heatmap shows average scores of the top 16 Compass-derived TIME concepts across four patient subgroups: inflamed responders, non-inflamed responders, inflamed non-responders, and non-inflamed non-responders. Patients are clustered within each subgroup by concept score similarity (**Extended Data** Fig. 5), revealing distinct TIME concept profiles. The top row indicates mean Compass-predicted response probability (*P_R_*) per cluster, demonstrating the relationship between concept activation patterns and clinical outcomes.

Patients classified by Compass as responders had improved overall survival (OS) compared to non-responders (**Fig. 5a**). Response probabilities outperformed concept-based risk scores in stratifying outcomes (**Methods 8**, **Supplementary Fig. S20**). Among n = 298 patients, those with *P_R_* ≥ 0.5 showed a 1-year OS rate of 86%, compared to 40% for those with *P_R_ <* 0.5, yielding a hazard ratio of 4.7 (log-rank *p* = 1.7 × 10^−7^). Compass outperformed TMB (HR = 1.67, *p* = 0.0038), PD-L1 IC2+ scoring (HR = 1.75, *p* = 0.0018), and IHC-based immune phenotype (HR = 1.85, *p* = 0.0042) (**Fig. 5b-d**)^23^.

To uncover the immunological features that inform Compass’s predictions, we analyzed TIME concept scores across immune phenotypes defined by CD8+ T cell infiltration patterns (inflamed, excluded, desert)^23^ (**Methods 9**). Inflamed tumors showed high activation of pro-inflammatory concepts, including Cytotoxic T Cell, IFN-*γ* Pathway, Immune Checkpoint, and Macrophage, as well as elevated Genome Integrity and Cell Proliferation (**Fig. 5e, Extended Data Fig. 4**). These concept scores positively correlated with the expression of genes underlying the learned concepts. Desert tumors lacked activation of pro-inflammatory concepts (Cytotoxic T Cell) and exhibited elevated scores for dysfunctional or deficient immune components, including NK Cell, Innate Lymphoid Cell, B Cell General, and Plasma Cell (**Fig. 5e**). These concept scores were negatively correlated with the expression of underlying genes, reflecting reduced infiltration or impaired functionality of the corresponding immune cells (**Supplementary Figs. S21-S23**). Immune-excluded tumors displayed intermediate features, with lower activation of inflammatory programs than inflamed tumors and weaker immunodeficiency signals than desert tumors (**Fig. 5e**). These tumors showed activated TGF-*β* Pathway and Endothelial concepts, consistent with immunosuppressive stromal remodeling and vascular exclusion.

Next, we examine how Compass resolves the heterogeneity of ICI responses within and between immune phenotypes. Patients were stratified into four groups based on their response and phenotype: inflamed responders and non-inflamed non-responders (groups showing the expected clinical outcome based on conventional immune phenotype), and non-inflamed responders and inflamed non-responders (groups showing an unexpected clinical outcome based on immune phenotype classifications). We cluster concept profiles for correctly predicted cases (**Fig. 5f, Extended Data Fig. 5**, **Methods 9**).

Inflamed responders formed two major clusters, both characterized by a strong activation of pro-inflammatory concepts (Cytotoxic T cell, IFN-*γ* Pathway) and absence of immuno-suppression or immuno-deficiency signals. These clusters differed in TMB-associated concept activity, particularly Genome Integrity and Cell Proliferation (**Extended Data Fig. 4b**), with one cluster showing strong activation of these concepts and the other lacking it. Non-inflamed responders clustered into two groups, exhibiting intermediate pro-inflammatory activation but lacking immunosuppressive signals, which distinguished them from non-responders. These patients comprised predominantly immune-excluded tumors, phenotypes generally associated with a lack of response to ICI, exemplifying how RNA-seq provides information on functional immune states beyond traditional IHC classification.

A greater degree of heterogeneity in grouping was observed among non-responders. Inflamed non-responders are of particular interest, because according to conventional immune phenotypes, they would be predicted to be responders, yet Compass correctly identified these patients as non-responders. These patients displayed variable pro-inflammatory levels but were unified by the presence of distinct resistance mechanisms. A first cluster showed strong activation of the Endothelial concept, reflecting angiogenesis and vascular remodeling processes that create a physical barrier to T cell infiltration, impairing immune cell trafficking. A second cluster exhibited high TGF-*β* Pathway concept activation; TGF-*β* signaling promotes stromal remodeling and fibrosis via cancer-associated fibroblast activation, excluding T cells from the tumor microenvironment^23, 24^. The remaining three clusters were characterized by CD4+ T cell immunosuppression and B cell deficiency. These clusters exhibited Th17-like CD4 programs with concurrent TGF-*β* signaling, consistent with a suppressive Th17 phenotype^25^ (**Supplementary Fig S22**). The deficiency of B cells, a cell type associated with the presence of tertiary lymphoid structures and survival benefit to ICI^26^, further suggests a loss or dampening of adaptive immune responses critical for anti-tumor immunity. Non-inflamed non-responders also exhibited heterogeneity, forming four distinct clusters. The largest cluster comprised patients without pro-inflammatory activation. Two clusters showed minimal pro-inflammatory signals with strong TGF-*β* activation, while another cluster exhibited strong endothelial concept-driven immune exclusion. These results show that Compass stratifies patients by survival outcomes and suggests resistance mechanisms between and within immune phenotypes.

### Compass generates personalized response maps to explain individual patient predictions

We developed personalized response maps that trace how Compass links patient-specific gene expression to predicted outcomes through its concept bottleneck across five levels: gene expression, encoder representations, granular immune concepts, aggregated TIME concepts, and the final response probability. Personalized response maps trace how individual gene activations in the personalized transcriptome propagate through biological pathways to immunological concepts, where they are integrated and transformed to produce the final prediction. We generated personalized response maps for representative patients from each immune-response cluster in **Fig. 5f**. Four illustrative examples are shown in **Fig. 6**, with additional cases provided in **Supplementary Figs. S24-S27**.

**Fig. 6:**
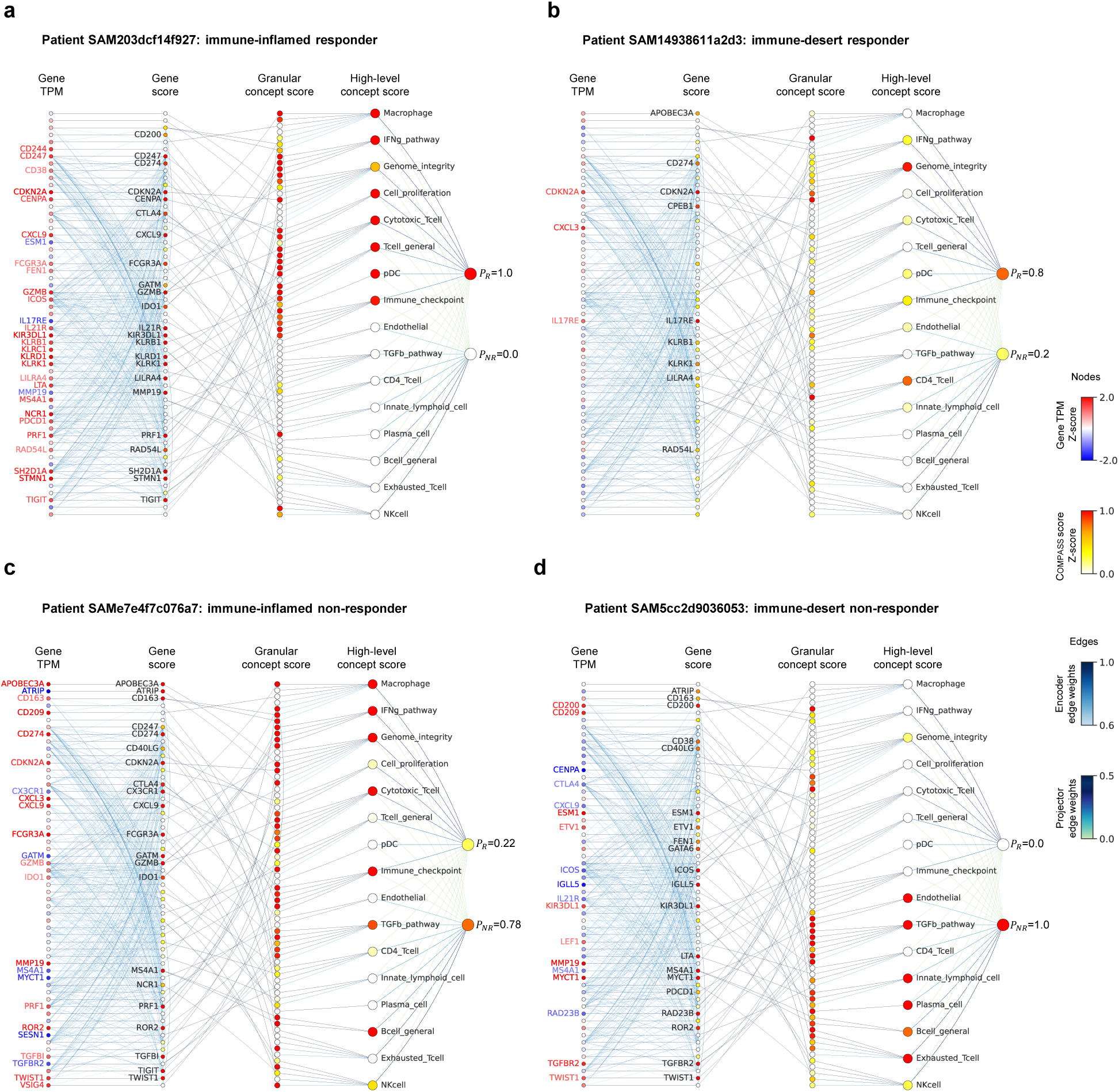
Personalized response maps explain ICI outcome predictions for individual patients. The maps trace how Compass connects input gene expression profiles to immunotherapy predictions through its interpretable concept hierarchy. Four representative patients from Fig. 5f clusters are shown (**a-d**, additional examples in **Supplementary Fig. S24–S27**). Each map displays: Z-score normalized gene expression levels (TPM), Z-score normalized gene and concept scores, predicted probabilities (response [*P_R_*] and non-response [*P_NR_*]), and model attention weights. For clarity, only the top-16 high-level concepts and the dominant contributing gene of each granular concept are shown (zoomed views: **Extended Data** Fig. 6). An interactive tool for exploring personalized response maps is available at the Compass website. **(a)** Inflamed responder — Patient SAM203dcf14f927 (cluster A_3_, *P_R_*= 1.0); **(b)** Desert responder — Patient SAM14938611a2d3 (cluster B_2_, *P_R_* = 0.8); **(c)** Inflamed non-responder — Patient SAMe7e4f7c076a7 (cluster C_5_, *P_R_* = 0.22); **(d)** Desert non-responder — Patient SAM5cc2d9036053 (cluster D_2_, *P_R_* = 0.0).

The response maps reveal how diverse immune profiles influence predicted outcomes. An inflamed responder shows broad IFN-*γ* and cytotoxic activation with minimal immunosuppression (**Fig. 6a**; response probability *P_R_* = 1.0), whereas an immune-desert responder shows strong Genome Integrity and moderate IFN-*γ* concept activations, consistent with a TMB-associated mechanism (**Fig. 6b**; *P_R_* = 0.80). In contrast, an inflamed non-responder shows co-activation of TGF-*β* signaling and B cell deficiency (**Fig. 6c**; *P_R_* = 0.22), and a desert non-responder shows dominant immunodeficiency features (**Fig. 6d**; *P_R_* = 0). Activation response maps link genes to concept activations, providing mechanistic context for predictions and supporting hypothesis generation.

## Discussion

Immune checkpoint inhibitors have improved outcomes across multiple cancer types, but response rates remain low and biomarkers such as tumor mutational burden and PD-L1 immunohistochemistry often fail to stratify patients accurately. Here, we present Compass, a concept bottleneck model that predicts immunotherapy response from transcriptomic data and provides mechanistic insight into response variability. Compass enables accurate prediction with concept-level interpretation to highlight resistance mechanisms and support trial-oriented hypothesis generation.

Compass complements emerging prognostic models that leverage real-world data^27^, clinico-genomic profiles^20, 28^, imaging^29^, and routine blood tests^30^ by providing mechanistically interpretable transcriptomic features. Unlike population-level risk predictors that rely primarily on systemic host factors (e.g., neutrophil-to-lymphocyte ratio, albumin), Compass encodes tumor transcriptomes into 44 immune concepts that capture tumor–immune biology linked to response and resistance. Recent transformer-based models^31^ integrate clinical and genomic features, including standard gene-signature scores. These models can incorporate Compass concept scores; in a Clinical Transformer predictor, Compass concepts improved survival prediction (**Supplementary Methods**, **Supplementary Figs. S29–S34**, **Supplementary Tables S11–S12**), supporting a modular strategy in which transcriptomic encoders augment multimodal predictors. Additional benchmarking against transcriptomic-based machine learning methods (ENLIGHT^13^, EaSIeR^16^ and IRnet^17^) further confirmed the state-of-the-art performance of Compass (**Supplementary Fig. S28**).

Building on its mechanistic foundation, Compass enables key applications in clinical development. Through multi-stage fine-tuning, the model can be rapidly tailored to specific indications and drug regimens without losing interpretability. Compass stratifies patients by mechanistically distinct features of response, including cytotoxic T cell activity, interferon-*γ* signaling, and TGF-*β* pathway activation. Through personalized response maps that trace the contributions of individual genes and immune concepts to predicted outcomes, Compass provides interpretable patient-level insights to support mechanistic interpretation of model predictions. The maps may support biomarker-driven enrichment and hypothesis generation, and concept trajectories could be used to monitor pharmacodynamic changes in trials.

Compass integrates pan-cancer transcriptomic pre-training with a concept bottleneck architecture that encodes 44 immune concepts derived from curated gene signatures. This design improves generalizability across cancers and checkpoint therapies while maintaining interpretability, including in small datasets via parameter-efficient adaptation. By modeling functional immune states, Compass resolves heterogeneity that conventional immune phenotypes cannot. It distinguishes responders within both inflamed and non-inflamed tumors and reveals resistance mechanisms in patients who fail treatment despite high immune infiltration. These mechanisms include TGF-*β* signaling, endothelial exclusion, CD4+ T cell dysfunction, and B cell deficiency. The findings extend prior results from the IMvigor210 study^23^, which linked TGF-*β* activity and tertiary lymphoid structures to immunotherapy outcomes.

Limitations of Compass include reliance on bulk RNA-seq, which lacks spatial resolution and may obscure signals from rare immune cell populations. Integrating single-cell or spatial transcriptomic data could improve resolution of cell-specific and spatially restricted immune states^32, 33^. Limited harmonized metadata, including biopsy site and clinical covariates, constrained site-specific and subgroup analyses. In addition, the lack of non-ICI comparator arms prevented us from separating prognostic from predictive signals. Addressing these limitations will require harmonized cohorts, covariate-adjusted analyses, and prospective controlled studies.

Predictions by Compass or similar models should never be used alone to deny immunotherapy. A further limitation is that the mechanistic concepts used by Compass have not yet been experimentally validated, and their current role is to generate testable hypotheses rather than serve as established biomarkers. Clinical deployment remains challenging and will require assay validation, cross-platform calibration, and reproducible inference pipelines. While promising, Compass remains an exploratory tool requiring prospective validation through clinical trials to define its appropriate use in therapeutic decision-making.

Compass links tumor transcriptomes to interpretable immune representations and supports biomarker discovery, mechanistic hypothesis generation, and patient stratification in immunotherapy trials. Its performance across cancer types and checkpoint therapies supports the use of mechanistically interpretable immune modeling in translational research and clinical development.

## Data availability

The pan-cancer TCGA datasets, including gene expression and mutation data, are available from the Genomic Data Commons data portal (https://portal.gdc.cancer.gov/, version 37). Clinical data for TCGA patients are provided in Liu *et al.*^34^. The datasets for the IMmotion150 cohort (EGA accession: EGAS00001002928)^35^, IMvigor210 cohort (EGA accession: EGAS00001002556)^22^ and IMvigor210CoreBiologies (v1.0.1, Ref^23^), Choueiri *et al.*^36^, Zhao *et al.* (SRA accession: PRJNA482620)^37^, Miao *et al.* (dbGAP accession: phs001493.v1.p1)^38^, and Kim *et al.* (ENA accession: PRJEB25780)^39^ are available from the Cancer Research Institute iAt-las (https://cri-iatlas.org/) and Synapse (https://www.synapse.org/, accession: syn10337516). The Ravi *et al.* cohort 1 (LUAD) and cohort 2 (LUSC) are non-small cell lung cancer (NSCLC) cohorts from the SU2C-MARK study^40^, available through dbGaP under accession number phs002822.v1.p1. The Snyder*et al.* cohort^41^ is available on Zenodo (https://doi.org/10.5281/zenodo.546110). The Rose *et al.*^42^ dataset is available on the Gene Expression Omnibus under accession ID GSE176307. The melanoma cohorts from Liu *et al.*^43^ (dbGAP accession: phs000452.v3.p1), Gide *et al.*^44^ (ENA accession: PRJEB23709), Riaz *et al.*^6^ (BioProject accession: PRJNA356761), Van Allen *et al.*^45^ (dbGAP accession: phs000452.v2.p1), Hugo *et al.*^46^ (GEO accession: GSE78220), and Freeman *et al.* (the MGH cohort, dbGAP accession: phs002683.v1.p1)^47^. Additional information, including patient metadata, gencode annotations, cancer codes, and Compass-concepts, are available on the Compass website (https://www.immuno-Compass.com/download).

## Code availability

The Python implementation of 22 baseline methods for immunotherapy response prediction, Compass code, model training, response prediction, and feature extraction used in this study is available on GitHub at https://github.com/mims-harvard/Compass/. An interactive online response prediction server based on various Compass models is at https://www.immuno-Compass.com/predict. The Compass-based gene, geneset, and concept feature extraction online tool is accessible at https://www.immuno-Compass.com/extract. The data processing pipeline for preparing Compass input from raw FASTQ or raw count mRNA TPM data is at https://github.com/mims-harvard/Compass-web/tree/main/mRNA pipeline. Compass tool to generate personalized response maps is available at www.immuno-Compass.com/explore/IMvigor210/.

## Data Availability

All publicly available data generated in this study are available from the corresponding author upon reasonable request. Access to controlled datasets requires direct application to the original study investigators, in accordance with their data sharing policies.

https://www.immuno-Compass.com/download/index.html

## Acknowledgements

We thank M. Cong for helpful discussions on immune checkpoint treatments. We thank S. Gao for advice and helpful suggestions on machine learning analyses.

## Funding

We gratefully acknowledge the support by NSF CAREER Award 2339524, ARPA-H Biomedical Data Fabric (BDF) Toolbox Program, Amazon Faculty Research, Google Research Scholar Program, AstraZeneca Research, GlaxoSmithKline Award, Roche Alliance with Distinguished Scientists (ROADS) Program, Sanofi iDEA-iTECH Award, Boehringer Ingelheim Award, Merck Award, Optum AI Research Collaboration Award, Pfizer Research, Gates Foundation (INV-079038), Chan Zuckerberg Initiative, Collaborative Center for XDP at Massachusetts General Hospital, John and Virginia Kaneb Fellowship at Harvard Medical School, Biswas Computational Biology Initiative in partnership with the Milken Institute, Harvard Medical School Dean’s Innovation Fund for the Use of Artificial Intelligence, and the Kempner Institute for the Study of Natural and Artificial Intelligence at Harvard University. M.M.L. is supported by The Ivan and Francesca Berkowitz Family Living Laboratory Collaboration at Harvard Medical School and Clalit Research Institute. T.H.N. acknowledges the support of NIH-2T32AI007512. W.S. acknowledges the support of Information Technology Center, Zhejiang University, and China Mobile Zhejiang Co., Ltd. Hangzhou Branch. Any opinions, findings, conclusions or recommendations expressed in this material are those of the authors and do not necessarily reflect the views of the funders. The funders had no role in study design, data collection and analysis, decision to publish or preparation of the manuscript.

## Authors contributions

W.S., D.M., and M.Z. designed the study. W.S. and M.Z. conceptualized the Compass models and algorithm. D.M., W.S., and T.H.N. provided and processed gene signatures. W.S., T.H.N., and M.M.L. collected and preprocessed the datasets. W.S. implemented the Compass code and conducted the benchmarking. I.M. performed reproducibility and robustness evaluations of the Compass models. W.S. and T.H.N. performed the patient survival analysis. Y.H. provided suggestions on Compass model. W.S., T.H.N., N.N., D.M. and M.Z. contributed to biological interpretation of the results. All authors contributed to writing the manuscript.

## Competing interests

D.M. and N.N are currently employed by F. Hoffmann-La Roche Ltd.

## Methods

The Methods describe: (1) dataset curation and pre-processing, (2) the Compass model, (3) self-supervised pre-training of Compass, (4) supervised fine-tuning for response prediction, (5) benchmarking Compass models against established methods, (6) multi-stage fine-tuning for drug- and disease-specific models, (7) SHAP analysis of important features, (8) overall survival analysis, (9) TIME concept analysis in the IMvigor210 cohort, and (10) personalized response maps generation.

## 1 Dataset curation and processing

### 1.1 The Cancer Genome Atlas (TCGA) datasets

Pre-training datasets are acquired from The Cancer Genome Atlas (TCGA) via the Genomic Data Commons (GDC) portal (version 37; GDC Portal), using TCGAbiolinks for data retrieval. To ensure cross-cohort compatibility with downstream ICI analyses, all RNA-seq data are uniformly processed through our standardized pipeline. Read alignment is performed against the GRCh38/hg38 reference genome using STAR (v2.7.5c), with gene features annotated according to GENCODE v36.

Raw counts are normalized by gene effective length and converted to TPM:

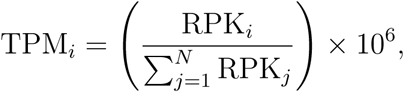

where *N* is the number of genes and

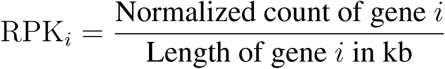

This normalization facilitates cross-sample comparability. Initial data included 60,660 genes across 11,274 samples. After excluding normal tissue samples, 10,534 samples remained. Further exclusions were applied for prior treatment samples and non-FFPE samples, resulting in 10,305 samples. Finally, aggregation to the patient level using the ”bcr patient barcode” key yielded 10,184 unique patient tumor samples. Protein-coding genes are selected (15,672 genes) based on overlap with the gene expression data from the clinical cohorts (see next section).

### 1.2 Immune checkpoint inhibitor (ICI) clinical cohorts

We curate 16 cohorts (**Fig. 2a**) spanning 7 cancer types, categorized into three groups: large cohorts (*>*100 patients), medium-sized cohorts (30-100 patients), and small cohorts (*<*30 patients). Publicly available RNA-seq data underwent uniform processing through the same standardized pipeline that was used to process the TCGA data (see previous section), converting raw sequencing data (FASTQ) to counts and TPM values. For cohorts with available raw data, FASTQ files were reprocessed; otherwise, TPM values were derived from the counts using TCGA-aligned gene lengths. To ensure cross-cohort consistency, all data were mapped to the same reference genome, correcting for potential differences in original genomic builds. To ensure reproducibility, researchers may download raw data using accession IDs in **Extended Data Table 1** and reprocess it via our publicly available code (https://github.com/mims-harvard/Compass-web/tree/ main/mRNA pipeline), which includes parameters for alignment, quantification, and TPM conversion. All steps rely on the GRCh38 reference genome and GENCODE v36 annotations to maintain cross-cohort consistency. Only pre-treatment samples are included. Responders are defined as patients achieving partial response (PR) or complete response (CR), while non-responders include those with stable disease (SD) or progressive disease (PD), per RECIST v1.1 BOR criteria as reported in source studies, unless otherwise noted. See **Extended Data Table 1** for cohort overview.

### Large immunotherapy cohorts

The IMvigor210 cohort (n=298) includes atezolizumab-treated bladder cancer patients (68 responders, 230 non-responders)^22^, with data sourced from the IMvigor210CoreBiologies (v1.0.1) R package and CRI iAtlas. The IMmotion150 cohort (n=165) comprises atezolizumab-treated renal cell carcinoma patients (48 responders, 117 non-responders)^35^. For the cohort from Liu *et al*. (n=107), post-treatment samples are excluded, retaining 41 melanoma patients (nivolumab/pembrolizumab) classified as responders and 66 as non-responders^43^. The Ravi-1 cohort (n=102) is a sub-cohort of the SU2C-MARK NSCLC study^40^, focusing on lung adenocarcinoma (LUAD) patients receiving PD-(L)1 ± CTLA4 inhibitors.

### Medium-sized immunotherapy cohorts

The Rose *et al*. cohort (n=89) includes bladder cancer patients treated with PD-(L)1 inhibitors (16 responders and 73 non-responders)^42^. The Gide *et al*. (n=73)^44^ cohort comprises melanoma patients receiving anti-PD-1 ± anti-CTLA4 (40 responders and 33 non-responders). Additional cohorts include: Riaz *et al*. (n=51) with nivolumab-treated melanoma patients (10 responders and 41 non-responders)^6^; Kim *et al*. (n=45) with pembrolizumab-treated stomach adenocarcinoma patients (12 responders and 33 non-responders)^39^; Van Allen *et al*. (n=39) with ipilimumab-treated melanoma patients (26 responders [CR/PR or SD with overall survival *>* 1 year] and 13 non-responders [PD or SD with OS *<* 1 year])^45^; and Freeman *et al*. (n=34) with melanoma patients from the MGH cohort treated with nivolumab, pembrolizumab, ipilimumab, or combination therapies (12 responders and 22 non-responders)^47^.

### Small immunotherapy ohorts

The Hugo *et al*. cohort (n=26) involves pembrolizumab-treated melanoma patients (14 responders and 12 non-responders by irRECIST)^46^. The Ravi-2 cohort (n=25), represents a SU2C-MARK NSCLC sub-study^40^ of squamous cell carcinoma (LUSC) patients treated with PD-1 or PD-L1 inhibitors (8 responders and 17 non-responders). For the Zhao *et al.* cohort (n=25), glioblastoma patients receiving nivolumab or pembrolizumab are classified as responders based on either: (1) post-treatment histopathology showing inflammatory response with minimal/no residual tumor cells, or (2) radiographic evidence of stable/shrinking tumor volume over six months^37^. The Snyder *et al*. cohort (n=21) includes atezolizumab-treated bladder cancer (BLCA) patients (7 responders and 14 non-responders)^41^. For renal cell carcinoma (KIRC) patients: the Choueiri *et al*. cohort (n=16) contains nivolumab-treated cases (3 responders and 13 non-responders)^36^, while the Miao *et al*. cohort (n=17) includes patients receiving PD-(L)1 ± CTLA4 inhibitors (5 responders and 12 non-responders)^38^, both sourced from CRI iAtlas. No new datasets are generated in this study.

## 2 Compass model

The Compass model comprises three key components. The first component, a transformer-based Gene Language Model (GLM), serves as the **encoder** to generate contextualized representations of individual genes. Next, a hierarchical **projector** transforms these gene-level embeddings into high-level biological concepts, including immune cell types and pathways. The final component, a **classifier**, performs immunotherapy response prediction from the concept representations, employing either a multilayer perceptron (MLP) or a non-parametric, similarity-based method for zero-shot prediction.

### 2.1 GLM encoder

The Gene Language Model (GLM) adapts natural language modeling techniques to transcriptomic data^32^, where each gene is treated as a token. Unlike natural language where tokens follow a clear sequential structure, gene expression profiles are inherently unordered and represented in tabular format. This difference renders positional encodings, such as fixed sinusoidal encodings used in NLP, suboptimal for capturing gene-gene relationships. Drawing inspiration from FT-Transformer^57^, which is designed for tabular data, we introduce a learnable gene-specific positional bias that enables the model to capture contextual interactions between genes in a biologically informed, data-driven manner.

**Gene abundance embedding.** Let **X**_gene_ ∈ R*^B^*^×^*^L^* denote the input gene expression matrix, where *B* is the batch size and *L* is the number of genes. Each gene is embedded into a *d*-dimensional latent space using a learnable embedding matrix **W** ∈ R*^L^*^×^*^d^*, initialized from a uniform distribution:

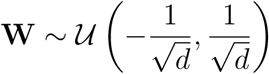

To generate expression-aware embeddings, we scale each gene’s embedding vector by its corresponding expression value. Specifically, the embedding for the *l*-th gene in the *b*-th sample is given by:

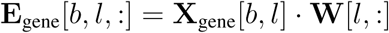

for *b* = 1*, … , B* and *l* = 1*, … , L*, resulting in the embedding tensor **E**_gene_ ∈ R*^B^*^×^*^L^*^×^*^d^*. This design enables the model to capture both gene identity (via **W**) and sample-specific abundance (via **X**_gene_) in the representation.

### Learnable positional encoding

To inject gene-specific inductive biases into the model, we introduce a learnable positional encoding matrix **P** ∈ R*^L^*^×*d*^, initialized in the same manner:

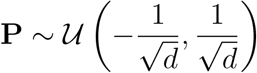

Each gene receives a unique, trainable positional vector **P**[*l,* :], which acts as a contextual bias. The final input embedding for each gene in each sample is computed by element-wise addition of the positional encoding:

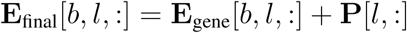

resulting in **E**_final_ ∈ R*^B^*^×^*^L^*^×^*^d^*. Unlike fixed encodings, this learnable scheme allows the model to adaptively encode gene-level functional relevance during training, thereby serving as a gene-aware bias that enhances the transformer’s capacity to model context-specific gene interactions.

### Cancer type token embedding

To account for pan-cancer heterogeneity, Compass integrates a cancer type token that interacts with gene tokens through attention mechanisms and is separately projected as a Cancer Type concept in the model’s hierarchy. To generate the cancer type token embedding, the 33 cancer types are first encoded as integers (0-32). This integer encoding serves as an index for looking up a learnable embedding matrix:

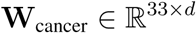

Given a batch of cancer type labels **X***_c_* ∈ R*^B^*, we perform a lookup to obtain their embeddings:

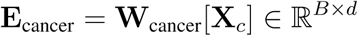

These embeddings are reshaped as **E**_cancer_ ∈ R*^B^*^×1×^*^d^*, and prepended to the gene embeddings along the sequence axis. This cancer type token interacts with gene tokens via self-attention and is later projected into a dedicated Cancer Type concept node in the concept hierarchy. As a robustness check, we ablated the Cancer Type concept during fine-tuning and re-evaluated leave-one-indication-out and leave-one-target-out generalization; performance decreased moderately but remained substantial (**Supplementary Fig. S10**).

### Transformer encoder for contextual learning

The full input to the transformer encoder is constructed by concatenating the cancer type token and gene embeddings:

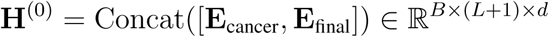

This tensor passes through a multi-layer transformer encoder composed of stacked self-attention and feedforward layers:

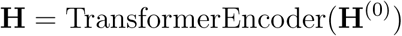

Within each layer, the self-attention mechanism enables each token to dynamically attend to all others, including gene-gene and gene–cancer-type interactions. The attention weights are computed via:

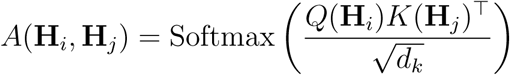

Given the large number of genes typically used in the model, full attention becomes computationally expensive. To address this, we adopt the Performer architecture^58^, which replaces standard attention with a linear approximation while preserving expressiveness. The architecture supports flash attention as an optional alternative to further reduce memory overhead and runtime.

The output tensor **H ∈** R*^B^*^×(^*^L^*^+1)×^*^d^* encodes the contextualized representations of both the gene expression profile and cancer type context. Rather than directly using these latent features for prediction, we project them into an explicit, biology-grounded concept bottleneck via a hierarchical projector (**Methods 2.2**). The resulting interpretable concept representation serves as the exclusive input to the downstream response classifier (**Methods 2.3**).

### 2.2 Concept-based hierarchical projector

Compass follows the concept bottleneck paradigm, in which inputs pass through a layer of human-interpretable concepts rather than mapping directly from latent features to predictions. In concept bottleneck models (CBMs), inputs map to a concept vector, and predictions are computed from these concepts, which supports interpretability and concept-level intervention^18^. Compass introduces this architecture for cancer transcriptomics by leveraging immunological gene sets and a hierarchical gene gene-set concept mapping to represent each patient’s tumor immune microenvironment. To transform gene embeddings into interpretable features, Compass uses a hierarchical projector with two outputs: (i) gene-set scores, *S*_Geneset_, where each score corresponds to a curated gene set, such as a pathway or immune cell signature; and (ii) concept scores, *S*_Concept_, which aggregate gene sets into functional modules, such as immune activation or immune suppression.

### Granular concept (gene set) aggregation

Given a curated gene set *G* = {*g*_1_*, g*_2_*, … , g_k_*} with *k* genes, we extract their contextualized embeddings from the GLM output 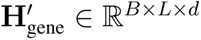:

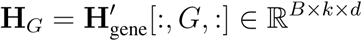

To aggregate the *k* gene vectors into a unified representation, we introduce a learnable attention vector **a***_G_* ∈ R*^k^*, initialized from a normal distribution and normalized via softmax:

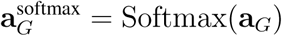

The attention-weighted gene set embedding is then computed as:

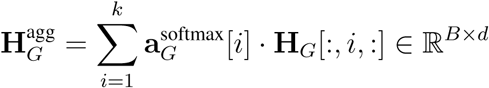

This aggregated vector is passed through a linear layer to produce the scalar score for gene set *G*:

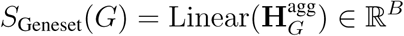

The full set of gene sets yields a tensor *S*_Geneset_ ∈ R*^B^*^×^*^K^*, where *K* denotes the total number of gene sets in the model. This mid-level representation captures modular biological information across curated pathways and cell types.

### High-level concept aggregation

Each high-level concept *C* consists of a subset of gene sets {*G*_1_*, G*_2_*, … , G_kC_* }, where *k_C_* is the number of gene sets associated with concept *C*. The corresponding gene set scores {*S*_Geneset_(*G*_1_)*, … , S*_Geneset_(*G_kC_* )} are aggregated using a second-level attention mechanism. A learnable attention vector **a***_C_* ∈ R*^kC^* is normalized via softmax:

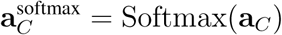

The high-level concept score is then computed as:

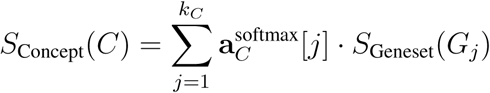

This attention-based aggregation allows each high-level concept to dynamically weight its constituent gene sets, enabling flexible and interpretable summarization of complex biological programs.

### Final concept representation

The Compass framework defines *M* = 43 high-level concepts derived from immune-related pathways, cell types, and functional groups. One additional concept represents the cancer type. The final concept representation for each sample is:

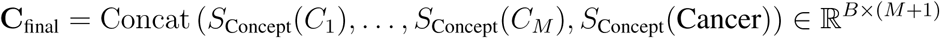

Here, the cancer type score is computed by projecting the cancer token embedding **H**_cancer_ ∈ R*^B^*^×*d*^ through a linear layer:

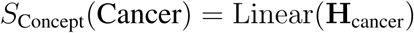

Together, the 44-dimensional vector **C**_final_ provides a biologically grounded and interpretable embedding of each patient’s tumor microenvironment, suitable for downstream prediction and mechanistic analysis.

### 2.3 Prediction module

Prediction module in the Compass model performs the conversion of high-level concept representations into probabilistic predictions of immunotherapy response. To accommodate both standard supervised learning and generalization to new cohorts or cancer types, Compass supports two distinct classifier types: a parametric multilayer perceptron (MLP) and a non-parametric cosine similarity-based Prototypical Network. The latter is referred to as the NFT (Non-Fine-Tuned) classifier, as it operates without gradient-based optimization during inference. These classifier heads provide complementary strengths and can be selected based on the availability of training labels and the desired generalization behavior.

### Parametric classifier using an MLP

The MLP classifier transforms high-level concept vectors into binary predictions via a trainable feedforward network. Given an input matrix **X ∈** R*^B^*^×44^, where *B* is the batch size and 44 is the number of concepts (43 biological concepts plus 1 cancer type), the inputs are first standardized:

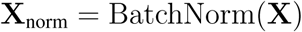

The normalized vectors are then passed through fully connected layers, producing output logits

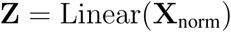

To calibrate output confidence, the logits are scaled by a learnable temperature parameter *τ* , defined as *τ* = exp(log temperature):

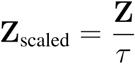

These scaled logits are transformed into probabilities using the softmax function:

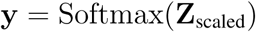

The learnable temperature *τ* allows the model to adjust the sharpness of its predictions and improves its ability to distinguish ambiguous cases, such as borderline responders.

### Non-parametric classifier using prototypes

The NFT (no fine-tuning) classifier adopts a prototypical network architecture^59^ that performs inference through similarity comparisons with labeled support examples, completely eliminating the need for model fine-tuning. This non-parametric approach is advantageous when limited training data prevents effective model adaptation. As il-lustrated in **Supplementary Fig. S1a**, the classifier begins with a support set of labeled patient embeddings, each represented by a high-level concept vector **f** R^44^ and a binary response label. The support examples are grouped by class *c* responder, non-responder , and a class prototype is computed by averaging the normalized vectors in each group:

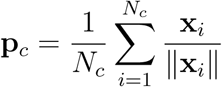

where **x***_i_ ∈* R^44^ is the concept vector of the *i*-th support sample in class *c*, and *N_c_* is the number of support examples in that class. The resulting prototypes **p***_c_* are unit-normalized to enable cosine-based comparison.

Given a query patient with concept vector **q**, the classifier computes the cosine similarity between the query and each class prototype:

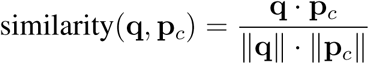

The similarity scores are scaled by a fixed temperature *τ* (typically 0.1), then passed through a softmax layer: logits*_c_*

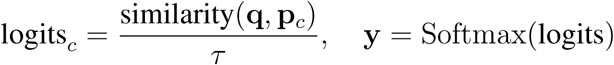

## 3 Pre-training Compass on TCGA

The Compass model was pre-trained on bulk RNA-seq data from 10,184 patients across 33 TCGA cancer types using a self-supervised triplet contrastive learning approach. This framework learns to map tumor transcriptomes (TPM values) into a 44-dimensional concept embedding space that captures TIME features. During training, each triplet consists of an anchor sample (a patient’s transcriptome), a positive sample (an augmented version of the same transcriptome), and a negative sample (a transcriptome from a different patient within the same cancer type). The model optimizes the embedding space to minimize cosine distance between anchor-positive pairs while maximizing separation from negative samples.

To address imbalance in TCGA cohort sizes across cancer types, we implemented balanced sampling with replacement, upweighting underrepresented cancer types during training. This ensures all cancer types contribute proportionally to the learned representations.

### Data augmentation

For contrastive learning, each anchor transcriptome undergoes stochastic transformation via one of two augmentation methods. Random masking independently zeros each gene’s expression value *x_i_* with probability *p_mask_* = 0.1 according to:

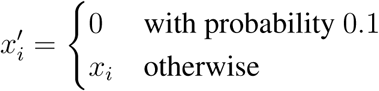

Alternatively, Gaussian jitter adds normally distributed noise to each value:

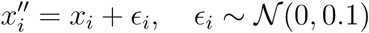

The augmentation method is selected randomly for each transformation event. We use random masking and Gaussian jitter, two widely used augmentations for contrastive learning on high-dimensional expression data. These augmentations promote robustness to technical variability, such as measurement noise. By analogy, computer vision uses domain-relevant augmentations such as rotation, cropping, and brightness perturbation. Future work could develop biologically grounded augmentation strategies for RNA-seq.

### Self-supervised training

The model is trained using a margin-based triplet loss function:

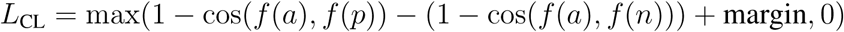

where *f* (*a*), *f* (*p*), and *f* (*n*) represent anchor, positive, and negative embeddings respectively, cos denotes cosine similarity, and a default margin of 1 is used.

Training was conducted on NVIDIA Tesla A100 80GB GPUs with batch size 128 and learning rate 1e-3 using the Adam optimizer. We reserved 1% of samples as a validation set for early stopping, with training halted if validation loss failed to improve for 10 consecutive epochs. For robustness, we performed three independent training runs with random seeds 24, 42, and 64, selecting the checkpoint with lowest validation loss for downstream use.

## 4 Fine-tuning Compass for response prediction

Following pre-training on TCGA, the Compass model is fine-tuned on ICI-treated cohorts to predict clinical response. To accommodate varying dataset sizes and quality across cohorts, we implemented four complementary fine-tuning strategies with differing levels of parameter adaptation: non-parametric zero-shot inference (Compass-NFT mode), linear probing (Compass-LFT mode), partial fine-tuning (Compass-PFT mode), and full model fine-tuning (Compass-FFT mode). These approaches provide a spectrum from maximal parameter efficiency (Compass-NFT) to full model adaptability (Compass-FFT).

All parametric modes (Compass-LFT, Compass-PFT, Compass-FFT) process the 44-dimensional concept vector through a single-layer dense classifier with 16 hidden units, generating logit outputs for binary response prediction. The trainable parameters vary substantially across modes: Compass-FFT updates all model parameters (approximately 1,018,784 total), including the GLM encoder and projector; Compass-PFT adapts only the classifier and projection layers (2,144 parameters); Compass-LFT modifies solely the classifier head (182 parameters); while Compass-NFT maintains frozen pre-trained weights with no trainable parameters, instead using prototypical inference based on cosine similarity in concept space (detailed in **Method 2.3**).

For parametric modes, models are optimized using cross-entropy loss with learning rates between 10^−3^ and 10^−2^ (slightly higher than the pre-training learning rate to promote faster adaptation to new domains), batch sizes of 8-16 (scaled to cohort size and GPU memory), and weight decay (10^−2^ and 10^−4^) tuned per mode and dataset. Internal cross-validation determined optimal training epochs, with FFT typically converging faster but being more prone to overfitting compared to the more stable LFT and PFT approaches in small datasets. All experiments ran on NVIDIA Tesla V100 GPUs, with final model selection based on cross-validated validation performance.

The Compass-NFT (no fine-tuning) mode represents a distinct non-parametric approach where labeled samples from the training cohort serve as a support set to compute responder/non-responder prototypes in the frozen 44-dimensional concept space. New samples are classified by cosine similarity to these prototypes (detailed in Method 2.3). This enables generalization to new domains without any additional gradient updates, making Compass-NFT ideal for low-data or zero-shot transfer scenarios.

## 5 Benchmarking immunotherapy response prediction models

### Overview of existing methods

We evaluated Compass against 22 established ICI response prediction methods (**Supplementary Table S1**), all implemented in our Python module. These methods enCompass three main categories: (1) target gene markers (PD1, PDL1, CTLA4, and their combination [GeneBio]^48^); (2) immune cell and functional signatures including Cytotoxic Immune Signature (CIS)^49^, T-effector-IFNg Signature (Teff)^50^, Neoadjuvant Response Signature (NRS)^51^, IFN-*γ* Signature Score (IFNG)^9^, Cytotoxic T Lymphocytes Markers (CTL)^7^, Tumor-Associated Macrophages (TAM)^7, 52^, T-cell Exhaustion (Texh)^7^, Chemokine Signature Score (CKS)^53^, Cancer-Associated Fibroblasts Signature Score (CAF)^54^, Roh Immune Score (IS)^55^, Immune Cytolytic Activity Score (ICA)^56^, CD8 Signature Score (CD8)^48^, MHC I Association Immune Score (MIAS)^10^, and the T Cell-Inflamed Gene Expression Profile Score (GEP)^10, 11^; and (3) comprehensive integrative methods including Tumor Immune Dysfunction and Exclusion Score (TIDE)^7^, Immuno-Predictive Score (IMPRES)^8^, Paired Gene Markers (PGM)^47^, and Network-Based ICI Treatment Biomarkers (NetBio)^48^.

For benchmarking purposes, we implemented standardized logistic regression models using the marker gene or predictive score from each baseline method (detailed in **Supplementary Table S1**) as input features. For each model, we performed hyperparameter optimization via scikit-learn’s GridSearchCV, employing L2 regularization with the LBFGS solver. All models incorporated balanced class weighting and were configured with a maximum of 10^10^ iterations to guarantee convergence. Through 5-fold cross-validation grid search across the regularization strength range (*C* ∈ [0.1, 1]), we identified the optimal parameter value using the area under the ROC curve (AUC) as scoring metric. This optimized C value was subsequently used to finalize each logistic regression model for comparative performance evaluation across all baseline methods.

### Cross-cohort and within-cohort model evaluation

To rigorously assess model performance across clinically relevant scenarios, we implemented three complementary validation strategies. First, leave-one-cohort-out validation evaluates cross-cohort generalization by training models on all available cohorts except one held-out cohort, then testing performance exclusively on the excluded cohort. Second, cohort-to-cohort transfer prediction provides a more stringent assessment of cross-cohort generalizability by training models on a single complete cohort and directly predicting outcomes for patients from an entirely different cohort. Third, within-cohort leave-one-patient-out validation assesses intrinsic predictive performance through iterative training on all patients except one within individual cohorts, followed by testing on each excluded patient - this approach provides robust performance estimates in homogeneous clinical settings while effectively controlling for overfitting. These strategies serve distinct purposes: both leave-one-cohort-out validation and cohort-to-cohort transfer prediction examine external validity across diverse clinical populations, while leave-one-patient-out validation focuses on optimizing and evaluating within-study performance.

Two baseline methods, NetBio and TIDE, were originally designed with specialized prediction approaches tailored to specific therapies and cancer types. NetBio selects the top 200 therapy-specific genes corresponding to the treatment regimen (‘PD1‘, ‘PDL1‘, ‘PD1 CTLA4‘, or ‘PD1 PDL1 CTLA4‘)^48^, whereas TIDE employs distinct scoring models for melanoma and non-small cell lung cancer (NSCLC), with all other cancer types processed through a generalized ’Other’ category^7^. To ensure consistency and transparency across all baselines, we further summarized in **Supplementary Table S1** the originally designed cancer type(s) and therapy/target contexts for NetBio, TIDE, and the remaining 20 published methods. During cross-cohort transfer evaluations, each method was applied in configurations aligned with the characteristics of the corresponding test cohort, following its original published implementation.

### Evaluation metrics and reference performance

We evaluated model performance using three complementary metrics: accuracy, Matthews correlation coefficient (MCC), and precision-recall area under the curve (PR-AUC). MCC and PR-AUC were included as robust measures for class-imbalanced datasets. Accuracy was computed by binarizing predicted response probabilities using a fixed decision threshold of 0.5 across all cohorts and analyses, rather than selecting cohort-specific thresholds; using a fixed threshold avoids cohort-dependent calibration and threshold-optimization effects and supports comparability across settings. To contextualize accuracy under class imbalance and facilitate interpretation of cross-cohort transfer results, we report cohort-specific baseline/reference metrics derived from the test cohort’s response distribution. These baselines represent the expected performance if predictions were made knowing only the responder prevalence in the test cohort (information unavailable to the models during prediction):

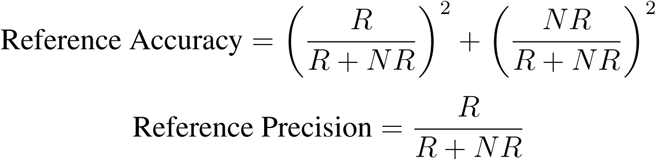

where R and NR denote the counts of responders and non-responders in the test cohort, respectively. These formulations naturally account for class imbalance, with perfectly balanced cohorts (R = NR) yielding reference values of 50% accuracy and 0.5 precision (see cohort-specific distributions in **Supplementary Fig. S8**). For our 240 cohort-to-cohort transfer evaluations, we defined successful transfer as cases where model accuracy surpassed the target cohort’s reference accuracy.

## 6 Multi-stage fine-tuning for drug- and disease-specific model development

### Overview of multi-stage fine-tuning

The multi-stage fine-tuning (MSFT) approach enables Compass to adapt to new therapeutic contexts with limited data availability through a hierarchical training strategy. This process begins with coarse fine-tuning on large, heterogeneous ICI-treated cohorts spanning multiple cancer types, followed by precise fine-tuning on smaller drug-or disease-specific datasets (**Fig. 4a**). The first fine-tuning stage establishes general ICI response prediction capabilities, capturing pan-cancer TIME features, while the second fine-tuning stage optimizes these features for specific drug mechanisms or clinical populations. This sequential approach enhances model transferability and robustness in data-limited settings where direct fine-tuning on small cohorts would risk overfitting.

We evaluated MSFT against two single-stage baselines (SSFT1: direct fine-tuning on drug- or indication-specific cohorts; SSFT2: fine-tuning on pan-cancer ICI cohorts) along with two reference models (PGM trained on SSFT1 data and baseline/reference performance for the test cohort as described in **Methods 5**).

For each assessment, clinical cohorts were split into two mutually exclusive groups: (1) pan-cancer ICI cohorts for coarse fine-tuning stage 1, and (2) drug- or disease-specific cohorts. The drug- or disease-specific cohorts were further split into disjoint training and test cohorts for rigorous cross-cohort transfer assessment. See **Fig. 4** and **Supplementary Table S7** for an overview of dataset configurations.

Importantly, when developing drug-specific models, drugs with the same target were excluded from the pan-cancer ICI cohorts. For example, when developing pembrolizumab-specific models, all other anti-PD1 drugs were excluded from the pan-cancer ICI dataset.

### Dataset splits

To evaluate drug-specific adaptation, we applied MSFT to three immune checkpoint inhibitors: atezolizumab (anti-PD-L1), pembrolizumab (anti-PD-1), and nivolumab (anti-PD-1). For atezolizumab, the drug-specific training cohort comprised 354 patients (IMvigor210: n=298 bladder cancer; Rose: n=35; Snyder: n=21), with testing performed on 176 kidney cancer patients (IMmotion150: n=165; Miao: n=2). Pembrolizumab-specific models were trained on 120 melanoma patients (Liu: n=62; Gide: n=32; Hugo: n=26) and evaluated on 78 gastric/lung cancer cases (Kim: n=45; Ravi-1 LUAD: n=33). Nivolumab models utilized 105 melanoma patients for training (Riaz: n=51; Liu: n=45; Gide: n=9) and 63 non-melanoma patients for testing (Ravi-1 LUAD: n=49; Ravi-2 LUSC: n=14). Consistent with our exclusion criteria, the pan-cancer ICI cohorts used for initial coarse fine-tuning excluded any cohorts treated with drugs sharing the same target mechanism (e.g., all anti-PD-1 therapies were excluded for pembrolizumab/nivolumab studies). For population-specific adaptation in lung adenocarcinoma (LUAD; **Supplementary Fig. S13**), the training data consisted of 69 Ravi-1 cohort patients treated with non-pembrolizumab therapies, while testing used a held-out set of 33 pembrolizumab-treated LUAD patients from the same cohort. As with drug-specific models, the pan-cancer ICI fine-tuning stage excluded all LUAD patients to prevent data leakage.

## 7 SHAP concept importance analysis

We employed SHAP (SHapley Additive exPlanations) analysis^60^ to quantify the contribution of each of the 44 high-level concepts to ICI response predictions. Using the Kernel SHAP implementation in the `shap` package (v0.46.0), we analyzed the partially fine-tuned (Compass-PFT) model after training on all available cohorts. The analysis was performed at both global (pan-cancer) and cancer-specific levels (BLCA, KIRC, SKCM, LUAD, STAD, GBM, and LUSC), as shown in **Supplementary Fig. S18**.

Because our primary objective was to determine the relative importance of 44 high-level concepts for response prediction, the SHAP analysis focused on the classifier module of the Compass model. A key step in the SHAP workflow was the selection of a background dataset: to capture the underlying data distribution while ensuring computational feasibility, we applied K-means clustering to generate 100 centroid points from the input data. In cases where fewer than 100 samples were available, the entire dataset was used as the background. SHAP values were computed separately for responder (R) and non-responder (NR) classes, and the final ranking of the 44 concepts was derived from the mean absolute SHAP values across all patients within each dataset.

## 8 Patient survival analysis

We examined Compass prediction of long-term clinical outcomes using its learned concept representations and predicted response probabilities. Specifically, the Compass model functions in two capacities: (1) as a feature extractor generating gene (*S*_Gene_), granular concept (*S*_Geneset_), and high-level concept (*S*_Concept_) features for downstream risk modeling, and (2) as a direct predictor of individual response probabilities. These approaches were evaluated on the IMvigor210 cohort, comparing Compass-derived features, Compass-predicted response probabilities, and established biomarkers (TMB, PD-L1 (IC) score, IHC immune phenotype). All survival analyses used overall survival (OS) as the endpoint, with censoring applied strictly according to the original study criteria^23^. For statistical analysis, we used the lifelines package (v0.27.8) to generate Kaplan-Meier survival curves, calculate log-rank test p-values, and compute hazard ratios (HRs) with 95% confidence intervals.

### Survival analysis using Compass-derived features

For survival analysis based on Compass-derived features, we used the Compass-PFT model trained on all cohorts except the IMvigor210 cohort (using leave-one-cohort-out approach). From this model, we extracted 132 granular concept and 44 high-level concept features as inputs for ridge-regularized Cox proportional hazards models (RCOX).

The RCOX models were trained on a combined dataset of 562 patients with available survival data, excluding the IMvigor210 cohort. Feature values were standardized using z-score normalization. Implementation used the scikit-survival package (v0.20.0), with the regularization parameter (alpha) optimized through five-fold cross-validation to maximize concordance index (C-index). Final risk scores were normalized to a 0-1 range using min-max scaling based on the training data.

For testing on the IMvigor210 cohort, feature standardization and risk score scaling were applied using the scalers fitted on the training data. Patients in the test set were stratified into high-risk and low-risk groups based on the top 10% risk score cutoff values derived from the training set. Using this procedure, the granular concept-based RCOX model classified 261 patients into the high-risk group and 37 patients into the low-risk group, while the high-level concept-based RCOX model classified 264 patients into the high-risk group and 34 into the low-risk group. Kaplan-Meier (KM) plots were generated based on this stratification (**Supplementary Fig. S20a**).

### Survival analysis using Compass-predicted response probabilities

The Compass-PFT model (trained excluding IMvigor210 as described above) generated response probabilities (*P_R_*), stratifying patients into responders (*P_R_* ≥ 0.5, n=42) and non-responders (*P_R_ <* 0.5, n=256). Kaplan-Meier analysis assessed survival differences between these groups (**Supplementary Fig. S20b**).

### Comparative analysis with established biomarkers

We evaluated Compass’s performance relative to three clinical biomarkers in the IMvigor210 cohort (limited to patients with TMB data, n=234). Patients were stratified using standard cutoffs for each biomarker^23^:

- TMB level: High (≥ 10 mut/Mb, n=97) vs low (*<* 10 mut/Mb, n=137).
- PD-L1 (IC) score: IC2+ (n=90) vs IC0/1 (n=144)
- IHC immune phenotype: Inflamed (n=56) vs non-inflamed (combined desert and excluded, n=149)

## 9 Analysis of Compass concepts in the IMvigor210 cohort

To explore the contribution and biological relevance of Compass concepts to response prediction in the IMvigor210 cohort, we analyzed concept scores generated by the Compass-PFT model that was trained on all ICI cohorts except IMvigor210 (leave-one-cohort-out approach). We computed the Pearson correlation coefficient between each concept score *S*_Concept_ and the predicted probability of response *P*_(_*_R_*_|_*_NR_*_)_ in the held-out IMvigor210 cohort. Concept scores were ranked based on the strength of their correlation with predicted responders (*R*) and non-responders (*NR*). The top 16 most strongly correlated scores were selected for downstream analysis. These included eight concepts positively associated with responder prediction (*P_R_*): Macrophage, IFN-*γ* Pathway, Genome Integrity, Cell Proliferation, Cytotoxic T Cell, T Cell General, pDC, and Immune Checkpoint; and eight positively associated with non-responder prediction (*P_NR_*): NK Cell, Exhausted T Cell, B Cell General, Plasma Cell, Innate Lymphoid Cell, CD4 T Cell, TGF-*β* Pathway, and Endothelial (**Supplementary Fig. S21**).

### Functional categorization and gene expression correlations of Compass concepts

To assess biological relevance of these TIME concepts, we analyzed associations between concept scores and expression levels of their constituent genes. Specifically, Pearson correlation coefficients were computed between each concept score and the TPM expression levels of its corresponding genes. Genes were then ranked from most negatively to most positively correlated, and the proportions of positive and negative correlations were summarized in **Supplementary Fig. S22**. Based on their immunological roles and gene correlation patterns, the 16 concepts were grouped into four functional categories: pro-inflammatory (Macrophage, IFN*γ* Pathway, Cytotoxic T Cell, T Cell General, pDC, Immune Checkpoint), TMB-associated (Genome Integrity, Cell Proliferation, see **Extended Data Fig. 4b**), immune-exclusion (TGF-*β* Pathway, Endothelial), and immune-deficiency (NK Cell, Exhausted T Cell, B Cell General, Plasma Cell, Innate Lymphoid Cell, CD4 T Cell).

### Analysis of personalized patient Compass profiles

To examine patient heterogeneity, we stratified patients into four subgroups based on their Compass-predicted response probabilities and immune phenotypes: inflamed responders (*P_R_* ≥ 0.5, *n* = 10), non-inflamed responders (*P_R_* ≥ 0.5, *n* = 13), inflamed non-responders (*P_R_ <* 0.5, *n* = 33), and non-inflamed non-responders with strong non-response predictions (*P_R_ <* 0.0001, *n* = 37). For each group, the Z-score normalized Compass concept scores (top 16) were visualized in a heatmap. Patients were clustered using hierarchical clustering with cosine distance and complete linkage to reveal intra-group patterns (Inflamed R: cluster A_1_, A_2_, and A_3_; Non-inflamed R: cluster B_1_ and B_2_; Inflamed NR: cluster C_1_, C_2_, C_3_, C_4_ and C_5_; Non-inflamed NR: cluster D_1_, D_2_, D_3_, and D_4_). The average concept score across patient clusters is shown in **Fig. 5f**. Each patient was annotated with their predicted *P_R_*, immune phenotype (inflamed, excluded, desert), tumor mutational burden (TMB-high vs. TMB-low, using a threshold of 10 mutations/Mb), and patient ID (**Extended Data Fig. 5**).

## 10 Generation of personalized response maps

Personalized response maps provide a hierarchical visualization of information propagation within the Compass model, enabling interpretation of how molecular features contribute to response prediction. By tracing the flow of information from input gene expression (*X*_GeneTPM_) through successive layers, gene scores (*S*_Gene_), granular concept scores (*S*_Geneset_), and high-level concept scores (*S*_Concept_), to the final predicted probability *P*_(_*_R_*_|_*_NR_*_)_, these maps reveal the biological reasoning underlying the model’s output for individual patients (**Fig. 6**, **Supplementary Figs. S24-S27**).

### Gene expression input layer

The first layer of personalized response maps represents the input gene expression matrix *X*_GeneTPM_ ∈ R*^B^*^×^*^L^*, where *B* is the number of patients and *L* is the number of genes. All input expression values are z-score normalized within the cohort, such that the value for gene *g* in a given patient reflects its relative expression compared to other patients:

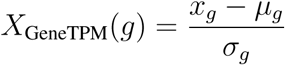

where *x_g_*is the log_2_(TPM + 1) value of gene *g*, and *µ_g_*, *σ_g_* are the cohort-level mean and standard deviation.

### Compass gene score layer

Gene scores *S*_Gene_ are computed by treating each gene as a singleton granular concept and applying the same projection process used for curated gene sets. Specifically, the transformer-based Gene Language Model (GLM) produces contextualized gene embeddings **X**_GeneGLM_ ∈ R*^B^*^×^*^L^*^×^*^d^*, where each gene embedding incorporates both the gene’s expression and its interactions with other genes through self-attention. Each gene embedding is then mapped to a scalar gene score using a shared linear projection layer:

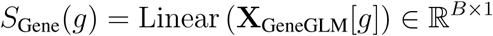

This formulation ensures that gene scores are directly comparable to granular concept scores, as they are derived using the same scoring mechanism. The resulting *S*_Gene_ reflects both individual gene activity and its contextual importance.

### Compass granular concept, high-level concept, and prediction layers

Gene scores *S*_Gene_ are aggregated into granular concept scores *S*_Geneset_ using attention-based weighting mechanisms that learn each gene’s contribution to its associated gene set. Each granular concept is projected into a scalar score using a shared linear layer applied to the weighted combination of gene embeddings.

The granular concept scores are further aggregated into high-level concept scores *S*_Concept_ ∈ R*^B^*^×4^^3^, where each score represents a broader functional module (e.g., immune cell types or signaling pathways). A second-level attention step is used to learn the relevance of each granular concept to its parent high-level concept. Finally, these scores are used to compute the overall response probability *P*_(_*_R_*_|_*_NR_*_)_, representing the likelihood that the patient will benefit from immunotherapy.

### Interactive clinical exploration tool

To generate personalized response maps for the IMvigor210 cohort, we employed the Compass-PFT model trained on all ICI cohorts except IMvigor210 (leave-one-cohort-out approach). These maps display z-score normalized features across hierarchical layers, highlighting inter-patient variation rather than absolute magnitude. Edge weights between layers represent the importance of each connection and are estimated by computing Pearson correlations between the *z*-scores of source and target nodes across the cohort.

An online Compass viewer enables interactive exploration, highlighting the top 16 high-level concepts (8 associated with response and 8 with non-response, as shown in **Supplementary Fig. S21**) and features exceeding user-specified thresholds (e.g., genes with |*z*| *>* 1 in the input layer or concepts with |*z*| *>* 0.5 in projection layers). Example maps in **Extended Data Fig. 6** illustrate how the interactive tool enables users to zoom in on particular concepts of interest (e.g., cytotoxic T cell activation) that drive predictions in a given patient.

---

**Extended Data Fig. 1:**
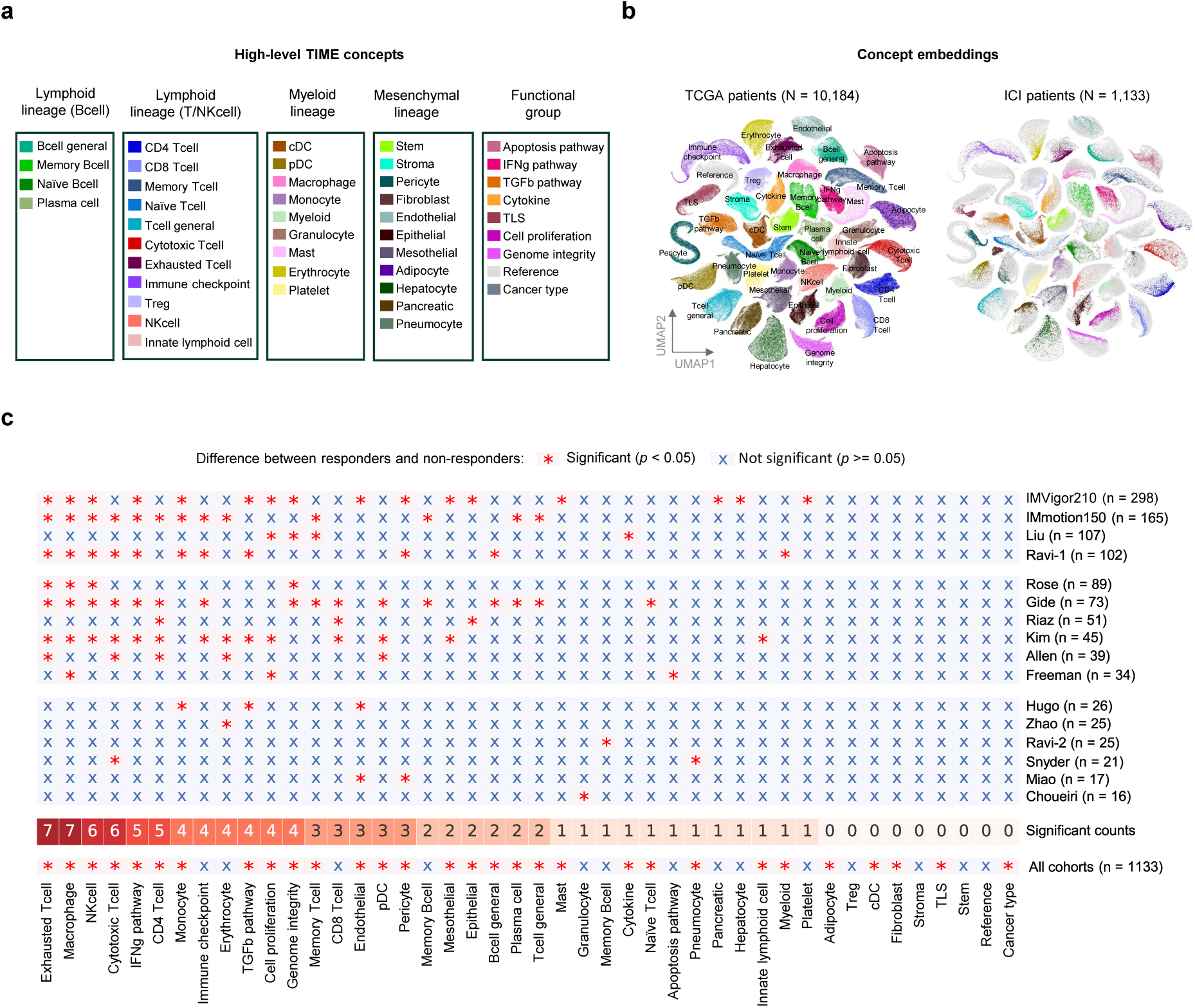
Analysis of Compass model concepts across cohorts and their association with treatment response. To investigate the biological relevance of the learned concept features, we analyze the 44 Compass-derived concepts and their associations with immunotherapy response. **(a) Overview of high-level TIME concepts in Compass.** The 44 concepts are organized into broader biological categories: 4 B cell–related concepts, 11 T/NK cell concepts, 9 myeloid lineage concepts, 11 mesenchymal lineage concepts, and 9 pathway/function-related concepts. Complete gene sets underlying these concepts are provided in **Supplementary Data S1**. Concept similarity based on gene overlap (Jaccard index) is shown in **Supplementary Fig. S14**, and variance across cancer types is analyzed in **Supplementary Fig. S15**. **(b) UMAP embedding of Compass concept representations using TCGA and ICI patients.** Each dot represents a patient’s 32-dimensional representation for one of the 43 learned concepts from the pre-trained model Compass-PT. The UMAP visualizes the 2D embedding of all such patient-level concept vectors, with clusters corresponding to distinct concepts. The left panel shows embeddings from TCGA patients, while the right panel overlays ICI patient embeddings (colored) onto the TCGA-derived UMAP (gray), demonstrating cross-population consistency. This alignment highlights the robustness and domain generalizability of the learned concept features. Additional UMAP and PCA visualizations for both granular and high-level concept embeddings are shown in **Supplementary Fig. S16-S17**. **(c) Predictive significance of Compass concepts.** The heatmap displays the statistical significance of TIME concepts for distinguishing responders from non-responders across 16 cohorts. Using leave-one-cohort-out validation, Compass-PFT models were iteratively trained while holding out each cohort for testing. The bottom row (”All cohorts”) shows significance when fine-tuned on the complete dataset. Significant associations (*p <* 0.05) are marked with red asterisks; non-significant results with blue crosses.

**Extended Data Fig. 2:**
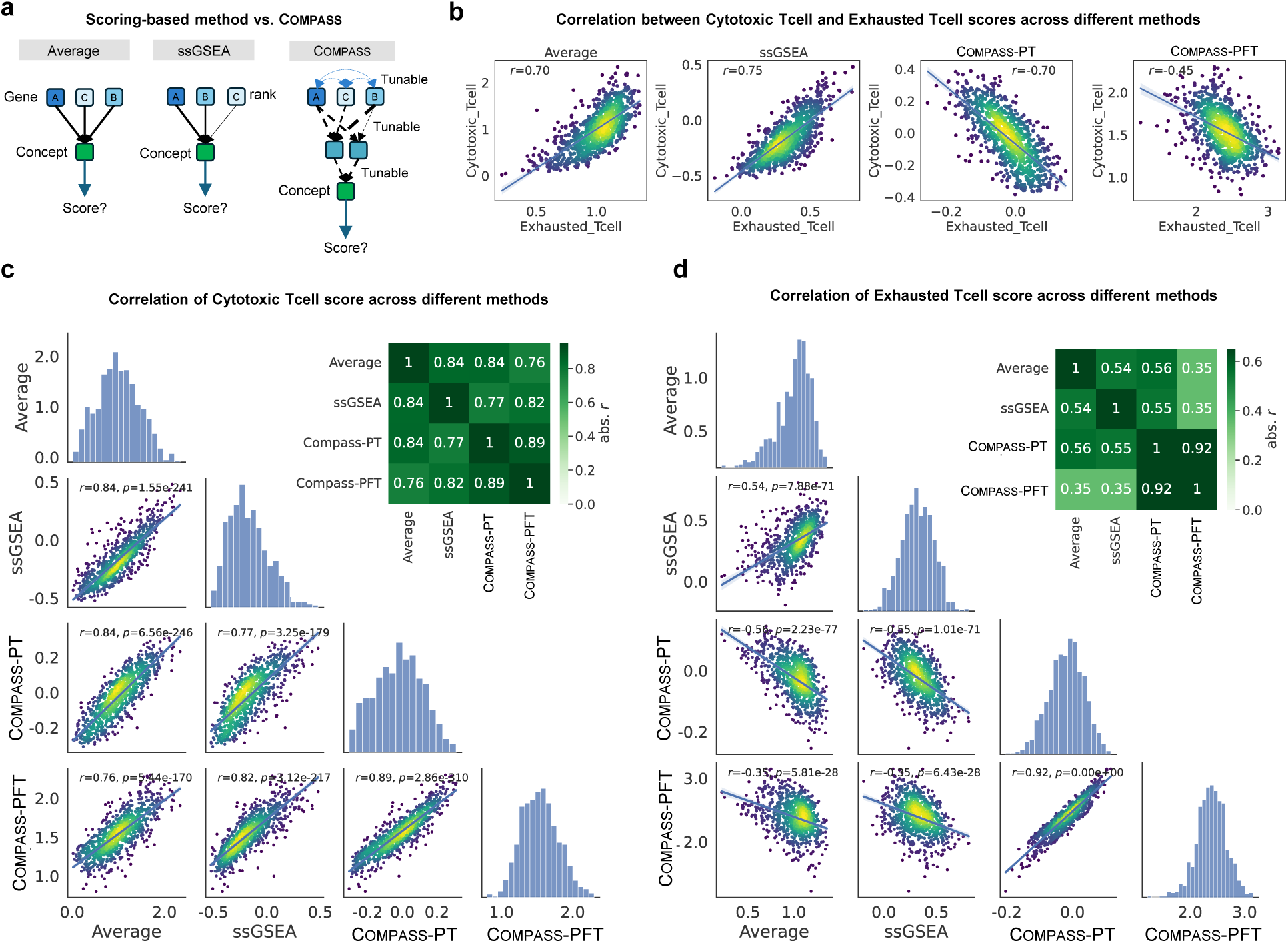
Comparison of Compass with conventional gene set scoring methods. To benchmark Compass against widely used scoring methods such as geometric averaging and ssGSEA, we assessed differences in concept score distributions and their predictive utility. **(a)** Conceptual comparison of Compass, geometric average, and ssGSEA. Geometric averaging assigns equal weights to genes, while ssGSEA is a rank-based, non-parametric method that emphasizes highly expressed genes. Both conventional methods treat genes as independent entities without modeling contextual interactions. In contrast, Compass encodes context-aware gene representations via a pre-trained gene-gene attention mechanism, and computes concept scores through a task-adaptable, fine-tunable hierarchical projection module, enabling transferability to new datasets. **(b)** Correlation of Cytotoxic T Cell and Exhausted T Cell scores across methods. High correlations are observed in ssGSEA (*r* = 0.70), geometric average (*r* = 0.75), and pretrained model Compass-PT (*r* = 0.70), while fine-tuned model Compass-PFT exhibits a notably reduced correlation (*r* = 0.45), reflecting its adaptation to ICI response prediction during fine-tuning. **(c)** Distribution and pairwise correlations of Cytotoxic T Cell scores across all methods. Despite different methodological assumptions, the resulting scores show strong agreement across approaches. **(d)** Distribution and pairwise correlations of Exhausted T Cell scores across all methods, demonstrating similar patterns of consistency.

**Extended Data Fig. 3:**
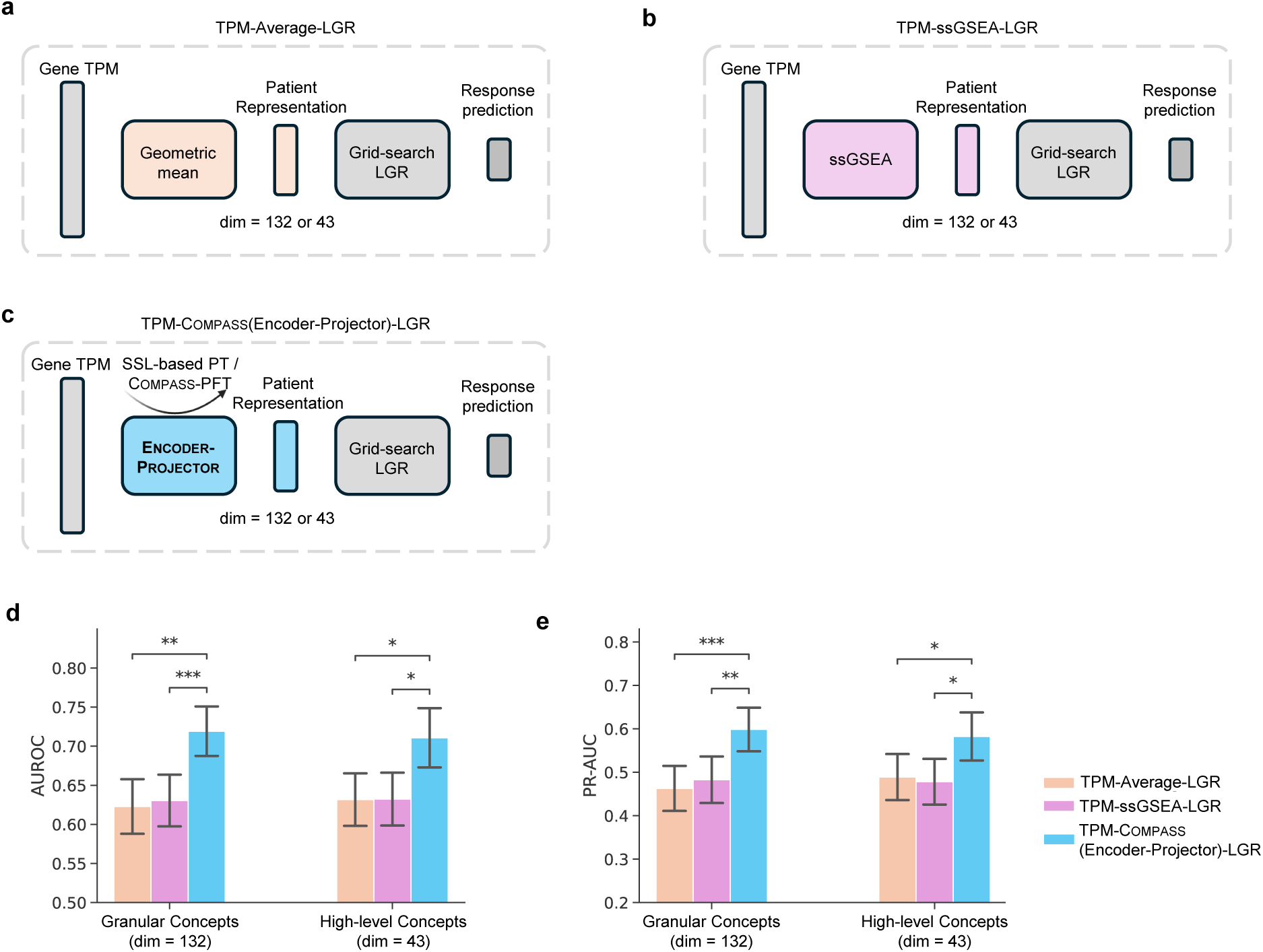
Benchmarking Compass against baseline patient representation strategies. Details are provided in **Supplementary Methods S2**. **(a)** Logistic regression with patient representation derived from the geometric mean of gene TPMs. **(b)** Logistic regression with patient representation derived from ssGSEA enrichment scores. **(c)** Logistic regression with patient representation derived from the SSL-based Compass-PT model or Compass-PFT model. **(d)** AUROC and **(e)** PR-AUC across 16 ICI-treated cohorts under the LOCO evaluation setting. Results are shown for both granular (132 dimensions) and high-level (43 dimensions) concept representations. Error bars represent the standard error across cohorts. Statistical significance was assessed using paired Wilcoxon signed-rank test: * *p <* 0.05, ** *p <* 0.01, *** *p <* 0.001.

**Extended Data Fig. 4:**
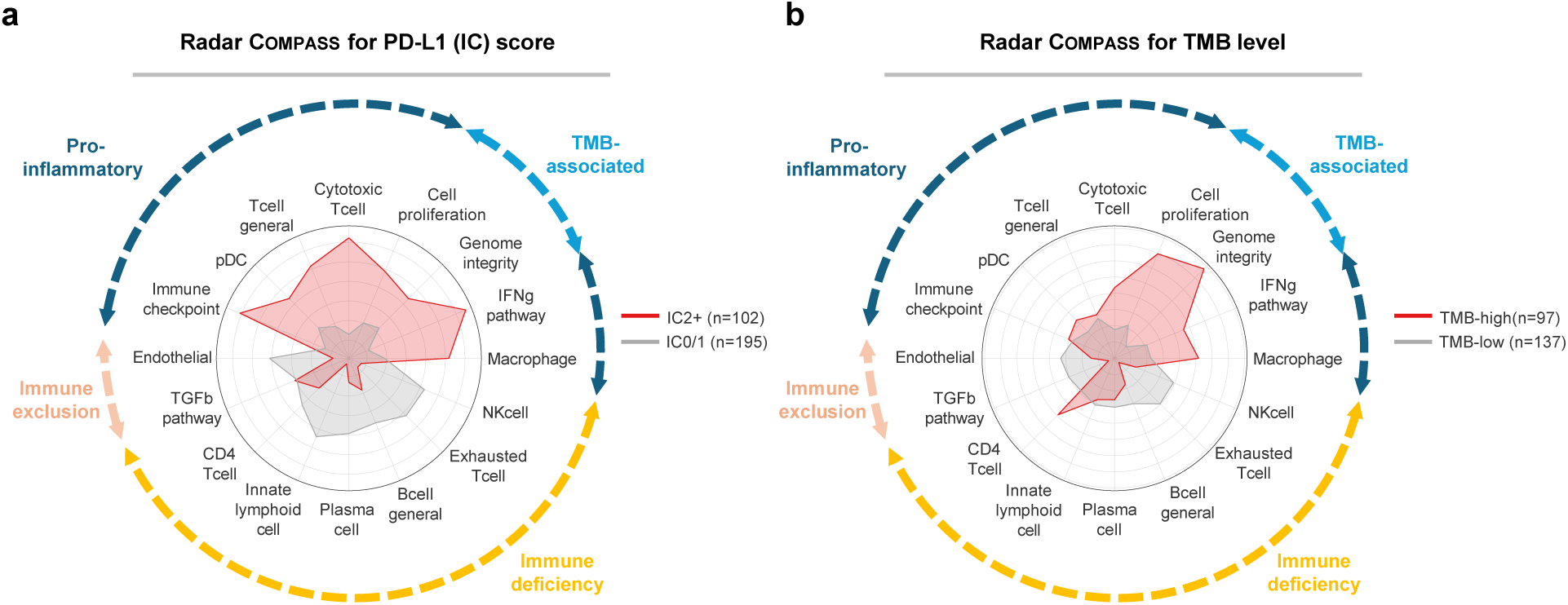
Radar Compass profiles for PD-L1 expression and TMB level in the IMvigor210 cohort. Radar plots compare immune and molecular features captured by Compass across patient subgroups stratified by **(a) PD-L1 immune cell (IC) score** and **(b) tumor mutation burden (TMB)**. Red-shaded regions represent patients with high PD-L1 expression (IC2+) or high TMB (≥ 10 mut/Mb), while gray-shaded regions represent those with low PD-L1 expression (IC0/1) or low TMB (*<* 10 mut/Mb). The top 16 immune pathways and cell types are shown along the radial axes, highlighting distinct immune and genomic characteristics between groups. Higher PD-L1 expression is associated with increased pro-inflammatory immune features, including Immune Checkpoint, Cytotoxic T cell, and IFN-*γ* Pathway activation. Higher TMB patients exhibit stronger TMB-associated features, such as Cell Proliferation and Genome Integrity, whereas TMB-low patients show reduced immune engagement and greater immune deficiency, marked by increased NK Cell and Exhausted T Cell features.

**Extended Data Fig. 5:**
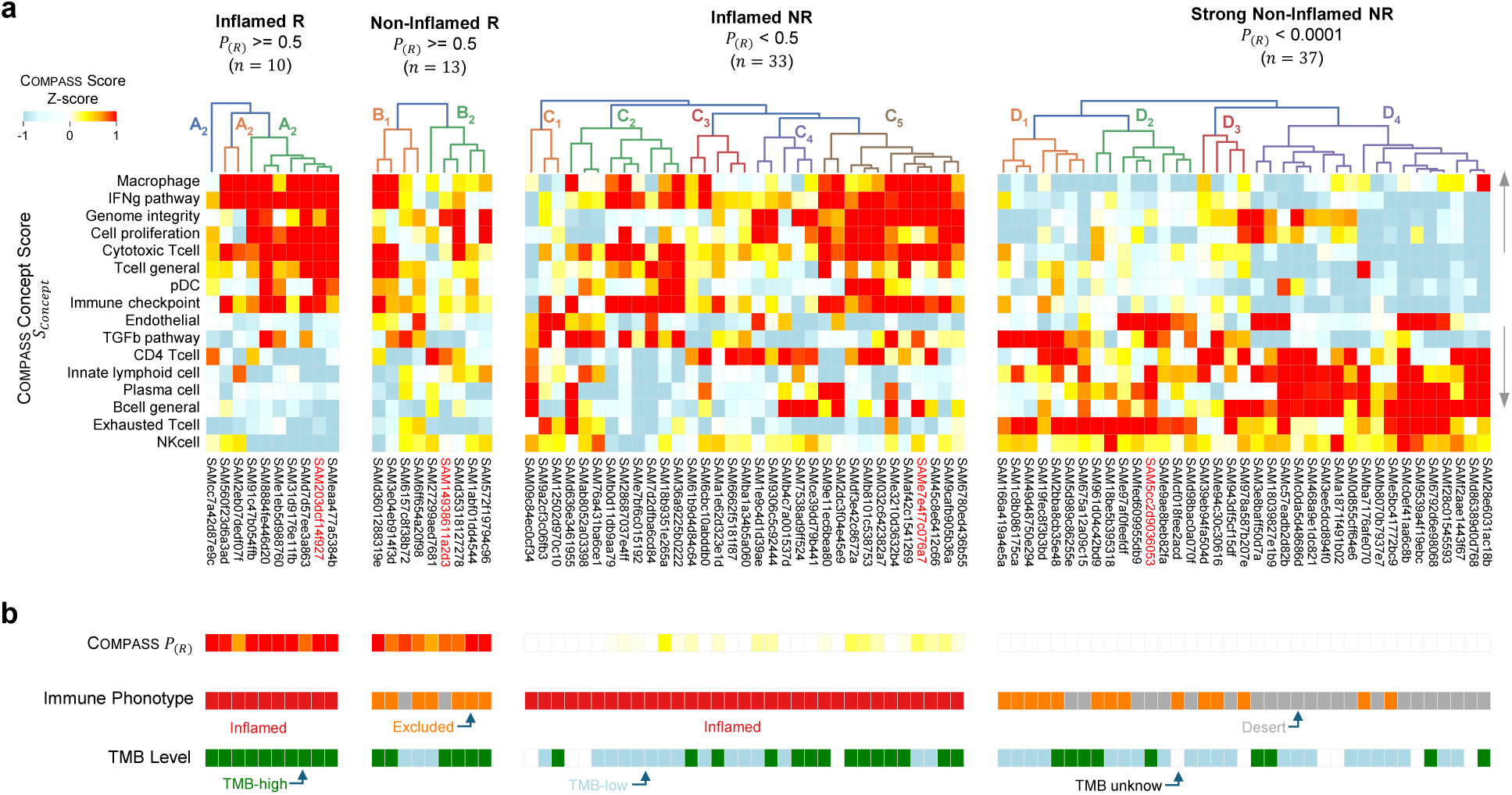
Heatmaps of Compass concept features, gene expressions, and clinical annotations across four patient subgroups in the IMvigor210 cohorts. Shown is an integrative analysis of Compass-based concept scores, gene scores, and gene expression across four patient subgroups stratified by immune phenotype and drug response. The patient order is consistent across all panels. The four subgroups include: (1) inflamed responders, (2) non-inflamed responders, (3) inflamed non-responders, and (4) strongly non-inflamed non-responders (**Methods 9**). **(a)** Heatmap of the top 16 Compass concept scores (*S*_Concept_), normalized as Z-scores, with hierarchical clustering applied within each patient subgroup. Rows represent different concepts, and columns represent patients. The four patients highlighted in red colors are further analysised in **Fig. 6**. **(b)** Patient-level annotations, including Compass-predicted response probability (*P_R_*), immune phenotype classification (inflamed, excluded, or desert), and TMB level (high, low, or unknown).

**Extended Data Fig. 6:**
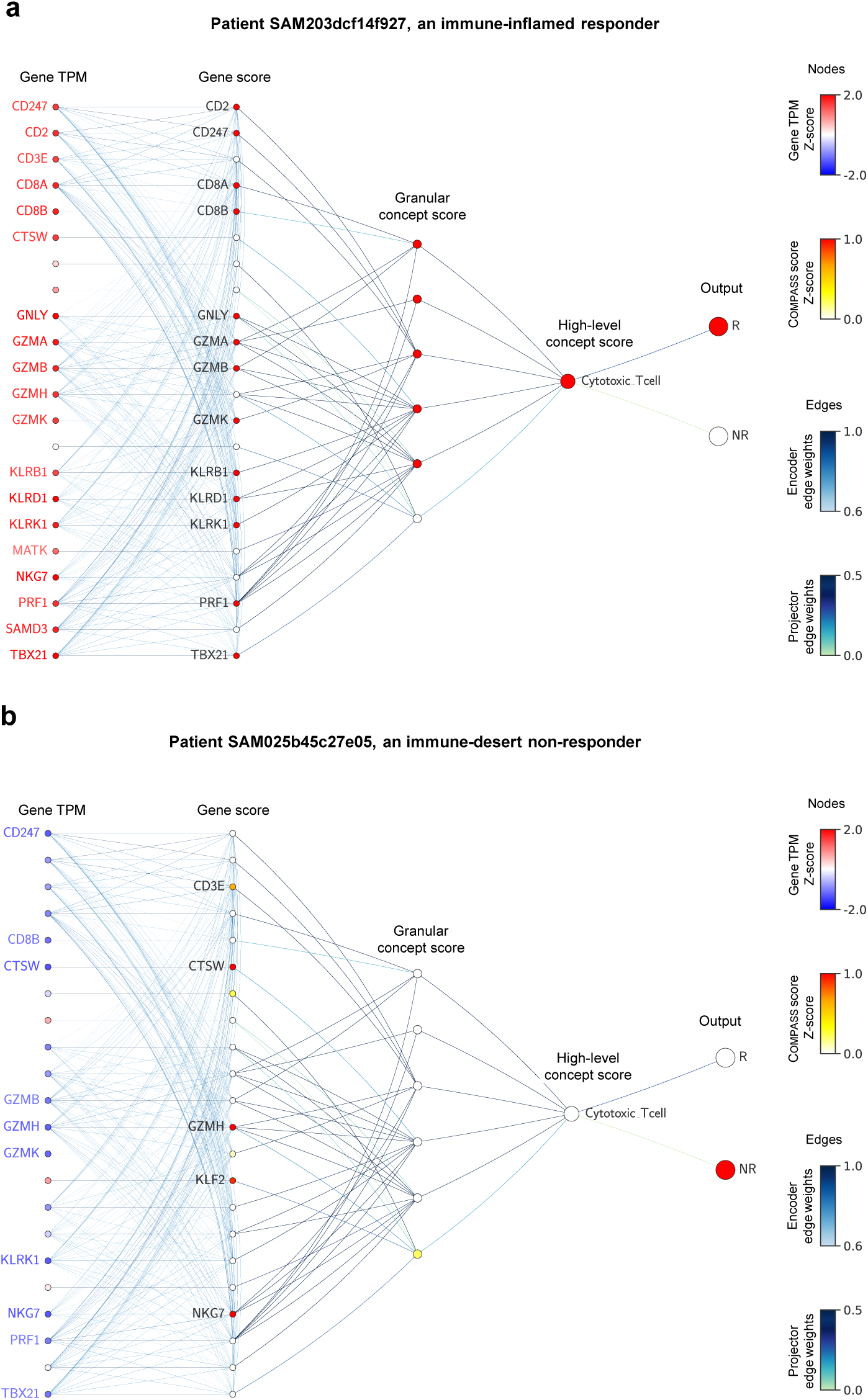
Zoomed-in views of personalized response maps for cytotoxic T cell activity. Our interactive visualization tool enables exploration of biological concepts and their associated gene signatures through adjustable parameters. Shown are representative examples focusing on the Cytotoxic T cell concept for two IMvigor210 patients with contrasting phenotypes. The interactive tool is available at www.immuno-Compass.com. **(a)** Immune-inflamed responder showing strong cytotoxic T cell activation. **(b)** Immune-desert non-responder demonstrating minimal cytotoxic T cell activity.

**Extended Data Table 1:** Overview of immune checkpoint inhibitor (ICI) clinical cohorts. Sixteen cohorts comprising a total of 1133 patients were used in our study, categorized based on size into large (greater than 100 patients), medium (30-100 patients), and small (less than 30 patients) groups. All RNA-seq transcript omics data listed in these cohorts are from pre-treatment samples. These cohorts span 7 cancer types: bladder cancer (BLCA), kidney cancer (KIRC), melanoma (SKCM), lung adenocarcinoma (LUAD), stomach adenocarcinoma (STAD), glioblastoma (GBM), and lung squamous cell carcinoma (LUSC). For each cohort, the number of patients, the number of responders (R) and non-responders (NR), accession IDs and references to the original studies are provided.

| Group | Cohort | Cancer Type | Patients (R / NR) | Reference | Data accession |
| --- | --- | --- | --- | --- | --- |
| Large cohort<br>(n = 672) | IMVigor210 | BLCA | 298 (68 / 230) | IMvigor210 Study Group. Lancet, 2017<sup>22</sup> | EGA:EGAS00001002556, IMvigor210CoreBiologies<sup>23</sup> |
|  | IMmotion150 | KIRC | 165 (48 / 117) | McDermott et al. Nat. Med., 2018<sup>35</sup> | EGA:EGAS00001002928 |
|  | Liu | SKCM | 107 (41 / 66) | Liu et al. Nat. Med., 2019<sup>43</sup> | dbGaP:phs000452.v3.p1 |
|  | Ravi-1 | LUAD | 102 (38 / 64) | Ravi et al. Nat. Genet., 2023<sup>40</sup> | dbGaP:phs002822.v1.p1 |
| Medium cohort<br>(n = 331) | Rose | BLCA | 89 (16 / 73) | Rose et al. Br. J. Cancer, 2021<sup>42</sup> | GEO:GSE176307 |
|  | Gide | SKCM | 73 (40 / 33) | Gide et al. Cancer Cell, 2019<sup>44</sup> | ENA:PRJEB23709 |
|  | Riaz | SKCM | 51 (10 / 41) | Riaz et al. Cell, 2017<sup>6</sup> | BioProject:PRJNA356761 |
|  | Kim | STAD | 45 (12 / 33) | Kim et al. Nat. Med., 2018<sup>39</sup> | ENA:PRJEB25780 |
|  | Van Allen | SKCM | 39 (13 / 26) | Van Allen et al. Science, 2015<sup>45</sup> | dbGaP:phs000452.v2.p1 |
|  | Freeman | SKCM | 34 (12 / 22) | Freeman et al. Cell Rep. Med, 2022<sup>47</sup> | dbGaP:phs002683.v1.p1 |
| Small cohort<br>(n = 130) | Hugo | SKCM | 26 (14 / 12) | Hugo et al. Cell, 2016<sup>46</sup> | GEO:GSE78220 |
|  | Zhao | GBM | 25 (11 / 14) | Zhao et al. Nat. Med., 2019<sup>37</sup> | SRA:PRJNA482620 |
|  | Ravi-2 | LUSC | 25 (8 / 17) | Ravi et al. Nat. Genet., 2023<sup>40</sup> | dbGaP:phs002822.v1.p1 |
|  | Snyder | BLCA | 21 (7 / 14) | Snyder et al. PLoS Med., 2017<sup>41</sup> | Zenodo: 10.5281/zenodo.546110 |
|  | Miao | KIRC | 17 (5 / 12) | Miao et al. Science, 2018<sup>38</sup> | dbGaP:phs001493.v1.p1 |
|  | Choueiri | KIRC | 16 (3 / 13) | Choueiri et al. Clin. Cancer Res., 2016<sup>36</sup> | CRI iAtlas data portal |
| **Total** | **16 cohorts** | **7 types** | **1133 patients** | **15 studies** |  |

## Notes

### Competing Interest Statement

D.M. and N.N are currently employed by F. Hoffmann-La Roche Ltd.The remaining authors declare no competing interests.

### Clinical Protocols

https://www.immuno-compass.com/

### Summary of Updates

This version of the manuscript has been revised to update the figures and extended data figures, revise the author contribution statement, and update the Supplementary Information.

