## Supplementary Information for "Generalizable AI predicts immunotherapy outcomes across cancers and treatments"

### Contents

**Supplementary Figures** **S2**

**Fig.** S1: LOPO performance of COMPASS-NFT across different feature levels . . . . . . . . . . S3
**Fig.** S2: LOCO performance of MCC and AUROC. . . . . . . . . . . . . . . . . . . . . . . . . . . S4
**Fig.** S3: LOPO evaluation and ACC performance in within-cohort LOPO across cohorts. . . . . S5
**Fig.** S4: Prediction MCC performance in within-cohort LOPO across cohorts. . . . . . . . . . . S6
**Fig.** S5: Prediction PR-AUC performance in within-cohort LOPO across cohorts. . . . . . . . . S7
**Fig.** S6: Prediction AUROC performance in within-cohort LOPO across cohorts. . . . . . . . . S8
**Fig.** S7: Success and failure in cohort-to-cohort transfer analysis. . . . . . . . . . . . . . . . . . S9
**Fig.** S8: Label distribution and reference accuracy across cohorts. . . . . . . . . . . . . . . . . . S10
**Fig.** S9: ROC curves for response prediction stratified by cancer type, drug, and target. . . . . . S11
**Fig.** S10: Ablation of the cancer-type token during fine-tuning. . . . . . . . . . . . . . . . . . . S12
**Fig.** S11: Prediction performance stratified by combinations of therapies. . . . . . . . . . . . . . S13
**Fig.** S12: Evaluation of model robustness across different sequencing platforms and biopsy sites. S14
**Fig.** S13: Comparison of single-stage and multi-stage fine-tuning strategies for LUAD-specific immunotherapy response prediction. . . . . . . . . . . . . . . . . . . . . . . . . . . . . . S15
**Fig.** S14: Jaccard similarity coefficient between 43 concepts based on overlapping genes. . . . . S16
**Fig.** S15: Variation of 44 concepts from COMPASS-PT model across different cancer types in TCGA and ICI patients. . . . . . . . . . . . . . . . . . . . . . . . . . . . . . . . . . . . S17
**Fig.** S16: PCA embedding of TCGA/ICI-patient concepts. . . . . . . . . . . . . . . . . . . . . . S18
**Fig.** S17: Granular concepts embeddings of TCGA patients and ICI-patients visualized by UMAP and PCA. . . . . . . . . . . . . . . . . . . . . . . . . . . . . . . . . . . . . . . . . . . S19
**Fig.** S18: Feature importance analysis in the COMPASS-PFT for predicting immunotherapy response. . . . . . . . . . . . . . . . . . . . . . . . . . . . . . . . . . . . . . . . . . . . . S20
**Fig.** S19: Comparison of concept scoring methods in distinguishing responders from non-responders.S21
**Fig.** S20: Kaplan-Meier survival curves stratified by risk groups or response predictions using the COMPASS model in the IMvigor210 cohort. . . . . . . . . . . . . . . . . . . . . . . . . S22
**Fig.** S21: Ranking of COMPASS high-level concepts by Pearson's correlation with response prediction in the IMvigor210 cohort. . . . . . . . . . . . . . . . . . . . . . . . . . . . . . S23
**Fig.** S22: Correlation of genes with COMPASS concept features in the IMvigor210 cohort. . . . . S24
**Fig.** S23: COMPASS concept features that positively-correlated to $P_{NR}$ in relation to immune cell infiltration and functional activity. . . . . . . . . . . . . . . . . . . . . . . . . . . . . S25
**Fig.** S24: Personalized response maps for two representative immune-inflamed responders. . . . S26
**Fig.** S25: Personalized response map for a representative immune non-inflamed responder. . . . S27
**Fig.** S26: Personalized response maps for four representative immune-inflamed non-responders. S28
**Fig.** S27: Personalized response maps for three representative non-inflamed non-responders. . . S29
**Fig.** S28: Benchmarking against transcriptomic ML models for response prediction. . . . . . . . S30
**Fig.** S29: Comparison of COMPASS and Clinical Transformer integration strategies. . . . . . . . S31
**Fig.** S30: Pretraining loss curves of Clinical Transformer models on TCGA data . . . . . . . . . S32
**Fig.** S31: Validation performance of Clinical Transformer survival models with the IMvigor210 cohort held out. . . . . . . . . . . . . . . . . . . . . . . . . . . . . . . . . . . . . . . . S33
**Fig.** S32: Comparison of Clinical Transformer survival models under baseline and transfer learning setups. . . . . . . . . . . . . . . . . . . . . . . . . . . . . . . . . . . . . . . . . . . S34

**Fig.** S33: Kaplan–Meier survival analysis of IMvigor210 patients stratified by different survival models. The IMvigor210 cohort served as the held-out test set for this evaluation. . . . . . S35
**Fig.** S34: Kaplan–Meier survival analysis of Gide cohort patients stratified by different survival models. The Gide cohort served as the held-out test set for this evaluation. . . . . . . . . . S36
**Fig.** S35: Calibration and decision-curve analysis evaluating the clinical utility of COMPASS . . S37
**Fig.** S36: Sensitivity analysis of `mask_p_prob` and `jitter_p_std` parameters during pretraining. S38
**Fig.** S37: Sensitivity analysis of the hard-negative sampling parameter $K$ during pretraining. . . S39

**Supplementary Tables** **S40**
**Table** S1: Sex distribution across immunotherapy cohorts analyzed in this study. . . . . . . . . . S40
**Table** S2: Overview of existing methods. . . . . . . . . . . . . . . . . . . . . . . . . . . . . . S41
**Table** S3: Performance comparison of methods in leave-one-cohort-out validation. . . . . . . . S42
**Table** S4: Accuracy and PR-AUC of different methods for predicting new indication cohorts. . . S43
**Table** S5: Accuracy and PR-AUC of different methods for predicting new ICI-treatment cohorts. S43
**Table** S6: Accuracy and PR-AUC of different methods for predicting new ICI-target cohorts. . . S43
**Table** S7: Summary of datasets for drug-specific model development . . . . . . . . . . . . . . S44
**Table** S8: Accuracy and PR-AUC of different drug-specific models. . . . . . . . . . . . . . . S44
**Table** S9: Benchmarking learned patient representations against fixed signature scoring (AUROC) S45
**Table** S10: Benchmarking learned patient representations against fixed signature scoring (AUPRC) S45
**Table** S11: Validation performance of Clinical Transformer survival models across feature representations and learning strategies. . . . . . . . . . . . . . . . . . . . . . . . . . . S46
**Table** S12: Test cohort evaluation of transformer-based survival models under the leave-one-cohort-out setting . . . . . . . . . . . . . . . . . . . . . . . . . . . . . . S46

**Supplementary Methods** **S47**
**Method** S1 Overview of signature scoring approaches: ssGSEA and geometric mean . . . . . . S47
**Method** S2 Benchmarking patient representations against signature scoring approaches . . . . . S47
**Method** S3 Benchmarking against genome-wide transcriptomic methods . . . . . . . . . . . . . S48
**Method** S4 Benchmarking against transformer-based methods for survival prediction . . . . . . S48
**Method** S5 Further ablations and sensitivity analyses of COMPASS . . . . . . . . . . . . . . . . S50

**Supplementary References** **S52**

### Supplementary Figures

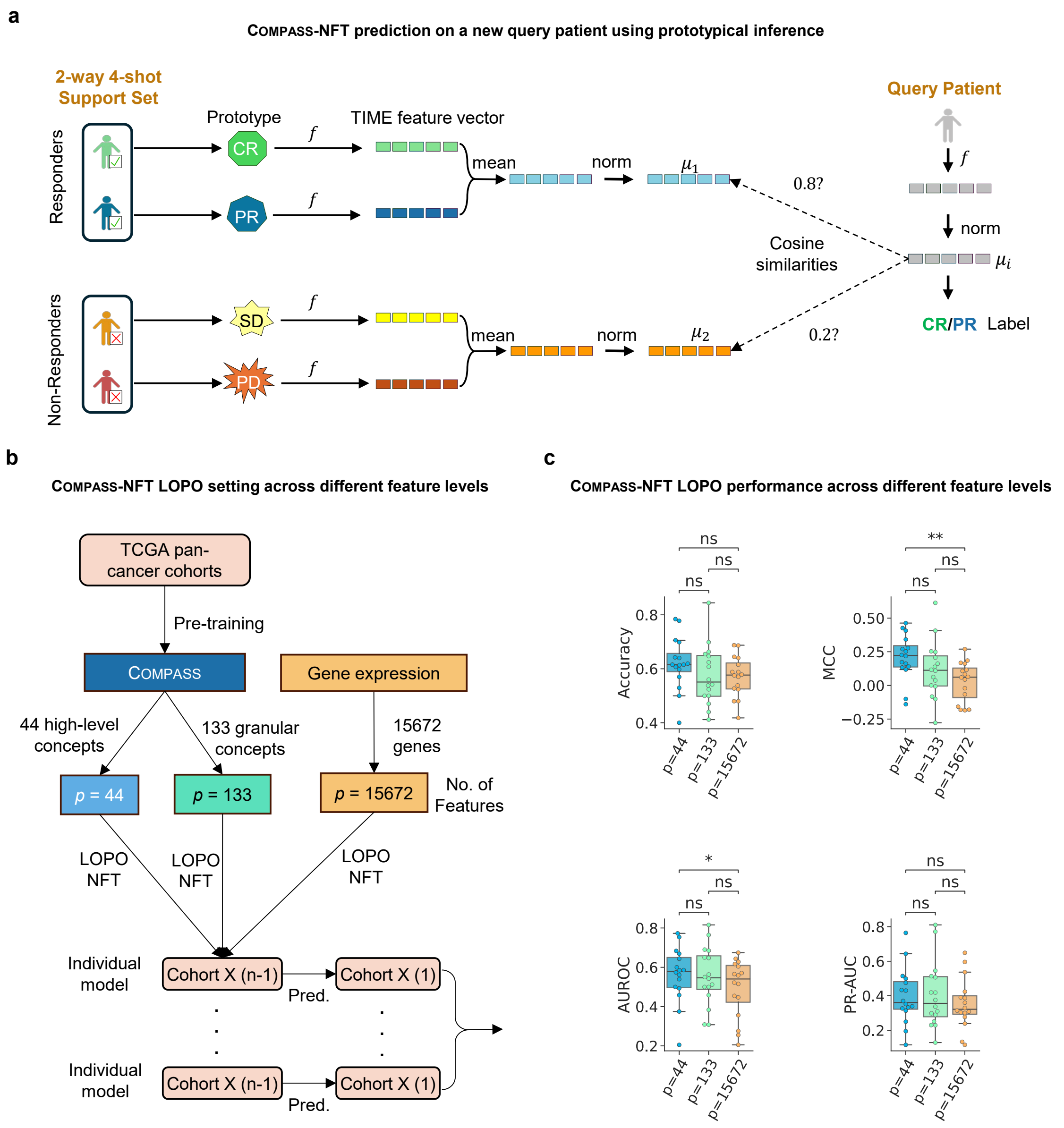


**Supplementary Fig. S1: Leave-one-patient-out (LOPO) performance of COMPASS-NFT across different feature levels.** To evaluate the predictive capacity of various feature representation levels for immunotherapy response, we tested three input types under the no fine-tuning (NFT) setting: raw gene expression, COMPASS-derived granular concepts, and high-level TIME concepts representing broader cell types or immune functions. Results indicate that feature representations at the concept level provide superior predictive performance compared to gene- or granular concepts-level inputs. **(a)** Prediction on a new patient using the COMPASS similarity-based no fine-tuning (NFT) mode. The query patient's feature vector is extracted using the pre-trained COMPASS model. Feature vectors of responders (CR/PR) and non-responders (SD/PD) from a support set are normalized, and mean vectors ($\mu_1$ and $\mu_2$) are computed. Cosine similarities between the query vector and each class mean are calculated, and the predicted label is assigned based on the higher similarity. **(b)** LOPO evaluation setup for COMPASS-NFT across different feature levels. The model is pre-trained on TCGA pan-cancer data and evaluated on individual cohorts using LOPO, based on one of three feature types: high-level concepts ($p$ = 44), granular concepts ($p$ = 133), or genes ($p$ = 15,672). **(c)** LOPO performance across different feature levels. Box plots summarize cohort-level performance (each dot represents one cohort; $n$=16) evaluated using Accuracy, Matthews correlation coefficient (MCC), AUROC, and PR-AUC. Statistical comparisons between feature levels were performed using two-sided paired $t$-tests with no adjustment for multiple comparisons. Exact $p$-values are as follows — Accuracy: high-level vs. granular $p = 0.093$, granular vs. genes $p = 0.866$, high-level vs. genes $p = 0.084$; MCC: high-level vs. granular $p = 0.083$, granular vs. genes $p = 0.100$, high-level vs. genes $p = 2.309 \times 10^{-3}$; AUROC: high-level vs. granular $p = 0.762$, granular vs. genes $p = 0.097$, high-level vs. genes $p = 0.048$; PR-AUC: high-level vs. granular $p = 0.831$, granular vs. genes $p = 0.062$, high-level vs. genes $p = 0.078$.

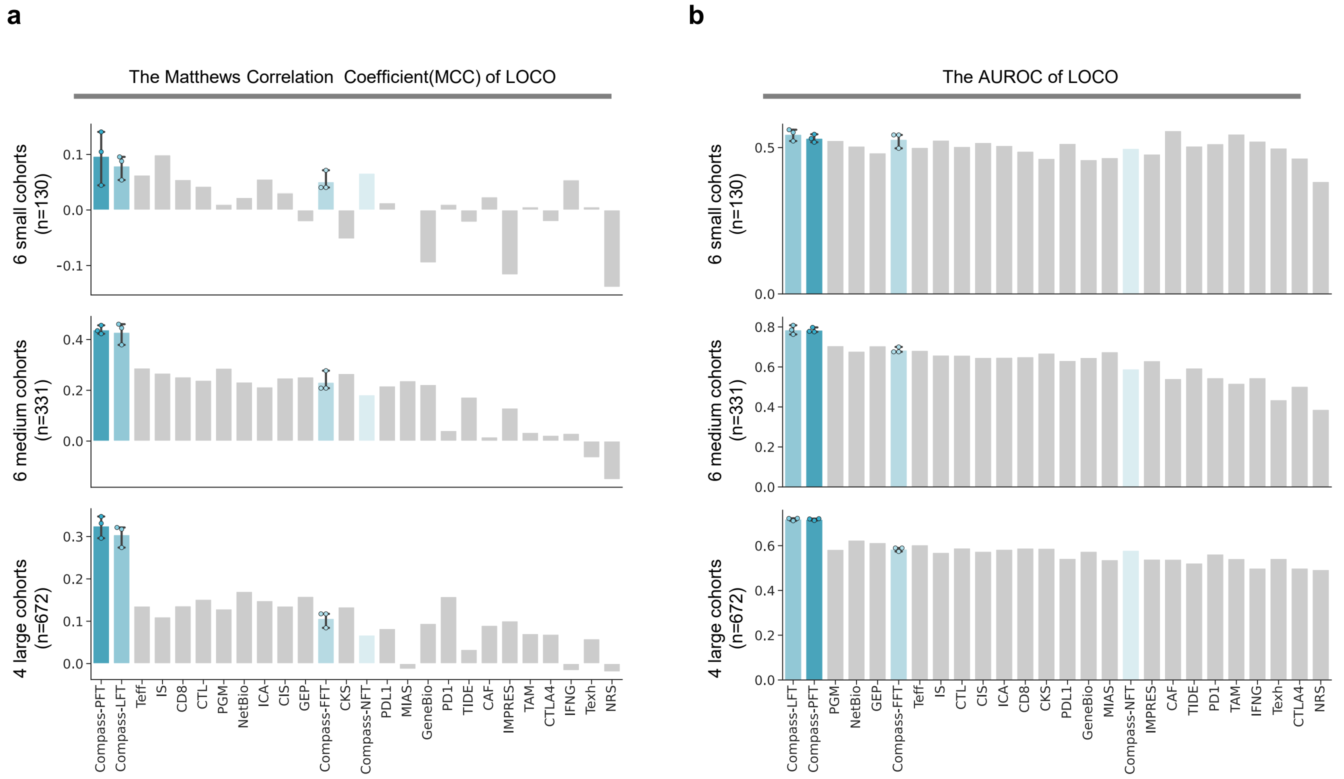


**Supplementary Fig. S2: LOCO performance of MCC and AUROC.** For the LOCO evaluation, the model was trained on 15 cohorts and tested on the remaining cohort in a round-robin manner. This process was repeated for each cohort, and a total of 22 baseline methods along with 4 fine-tuning (FT) modes of COMPASS were evaluated. Prediction accuracy was measured by grouping cohorts into small (6 cohorts, n = 130), medium (6 cohorts, n = 331), and large (4 cohorts, n = 672) categories to ensure meaningful statistical comparisons. For the 4 fine-tuning modes of COMPASS, the experiments were repeated 3 times using different random seeds. Bars show the mean performance, and error bars represent the 95% confidence interval across three repeats. **(a)** The Matthews correlation coefficient (MCC) performance across small, medium, and large cohort groups. **(b)** Receiver operating characteristic performance (AUROC) performance across small, medium, and large cohort groups. Accuracy and PR-AUC results are shown in **Fig. 2c–d**.

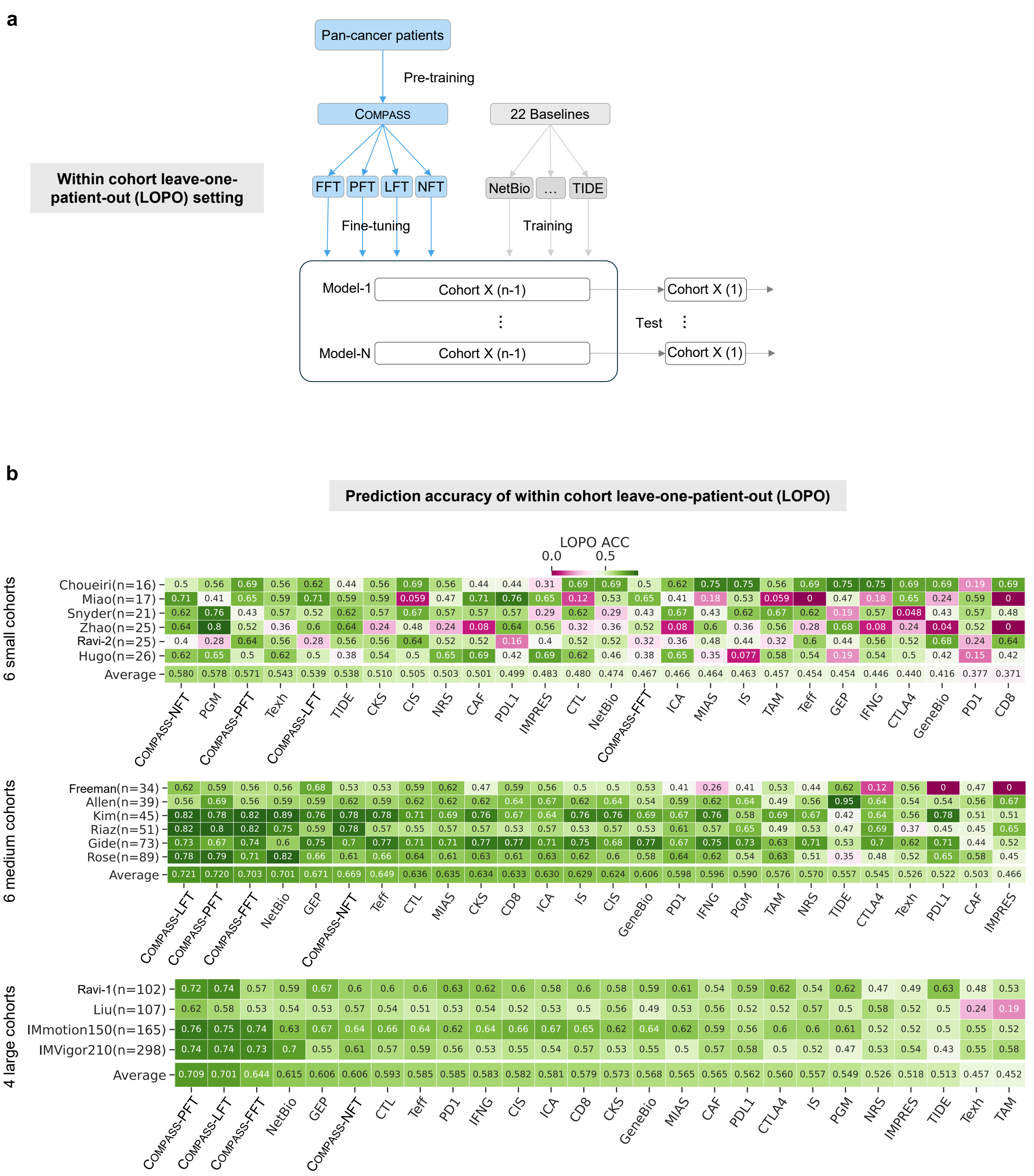


**Supplementary Fig. S3: Leave-one-patient-out (LOPO) evaluation and ACC performance in within-cohort LOPO across cohorts. (a)** Overview of the within-cohort LOPO experimental setup. The COMPASS model, pre-trained on TCGA pan-cancer cohorts, was fine-tuned using four different fine-tuning (FT) modes—Full Fine-Tuning (FFT), Partial Fine-Tuning (PFT), Linear-probing Fine-Tuning (LFT), and No Fine-Tuning (NFT)—and compared against 22 baseline methods including NetBio, GEP, TIDE, among others. For each cohort, a within-cohort LOPO setup was employed, where one patient was left out for validation during each iteration, and prediction accuracy was assessed. **(b)** Prediction accuracy of within-cohort LOPO in each cohort. The heatmap presents the LOPO accuracy (LOPO ACC) results for each cohort, grouped into 6 small cohorts, 6 medium cohorts, and 4 large cohorts. Results for each fine-tuning mode and baseline method are reported, with darker green indicating higher accuracy and pink highlighting the lowest-performing cases. Average accuracy across cohorts in each group is also provided for comparison.

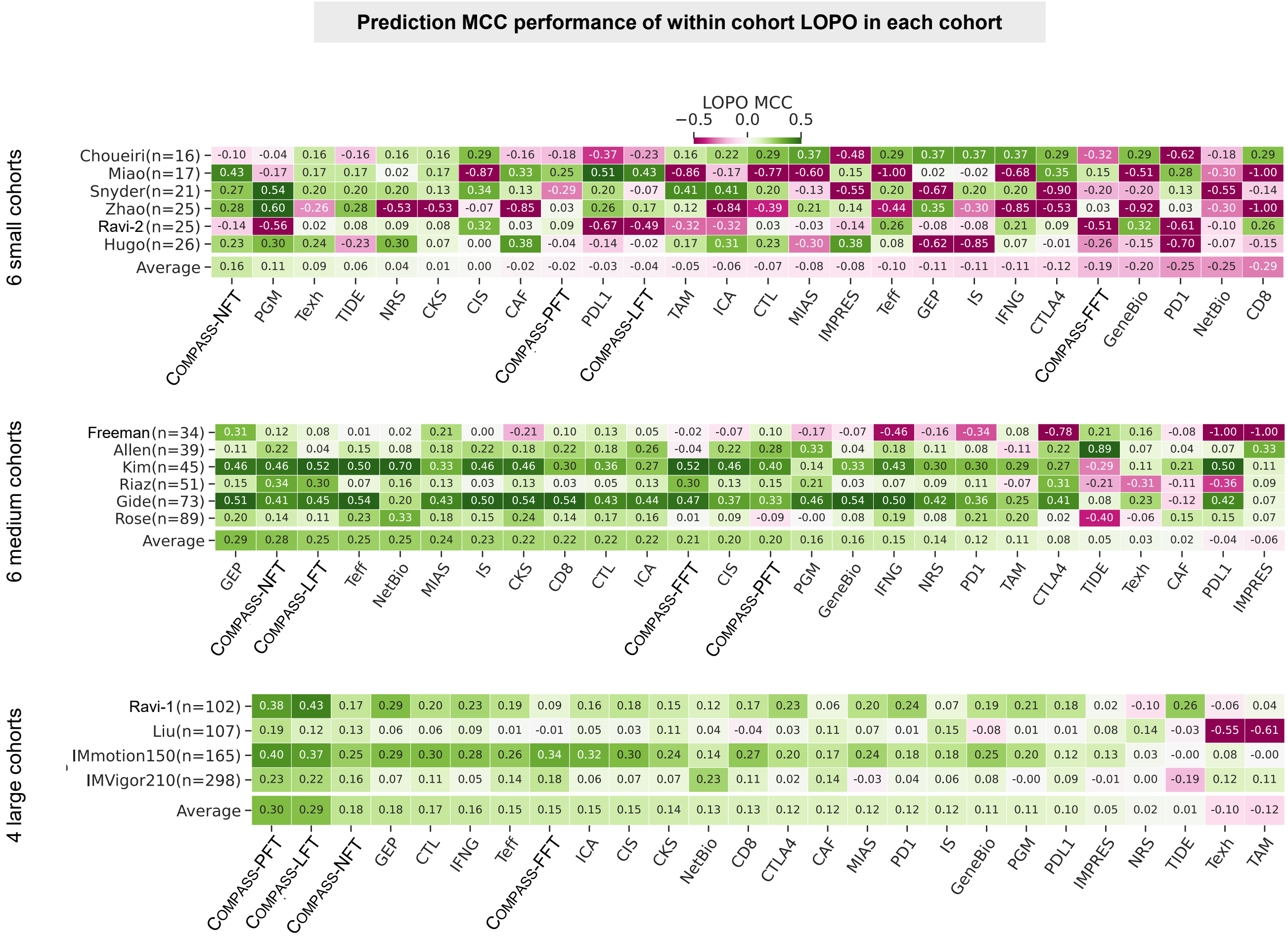


**Supplementary Fig. S4: Prediction MCC performance in within-cohort LOPO across cohorts.** The heatmap shows the Matthews correlation coefficient (MCC) performance for various methods across different cohorts using within-cohort leave-one-patient-out (LOPO) cross-validation. Darker green shades represent higher MCC values (better performance), while pink shades indicate lower MCC values (worse performance). On average, small cohorts show lower performance across all methods. Methods like TIDE and NRS perform relatively better in small cohorts but exhibit poorer results in medium and large-sized cohorts. Conversely, medium and large cohorts tend to achieve higher MCC values, with methods such as COMPASS-NFT and COMPASS-LFT performing more consistently well across these datasets.

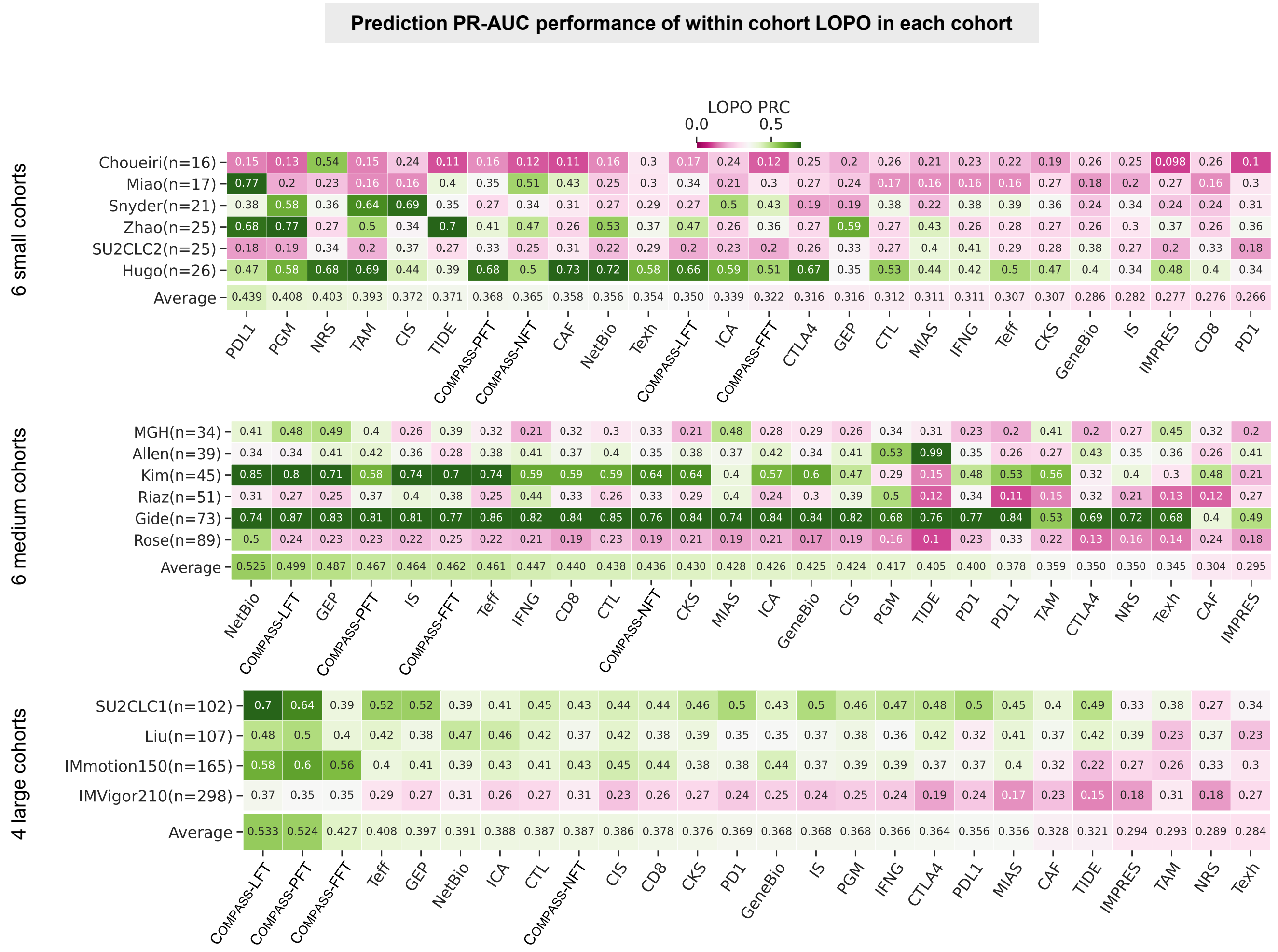


**Supplementary Fig. S5: Prediction PR-AUC performance in within-cohort LOPO across cohorts.** The heatmap shows the PR-AUC performance for various methods across different cohorts using within-cohort leave-one-patient-out (LOPO) cross-validation. Darker green shades represent higher PR-AUC values (better performance), while pink shades indicate lower PR-AUC values (worse performance). On average, small cohorts show lower performance across all methods.

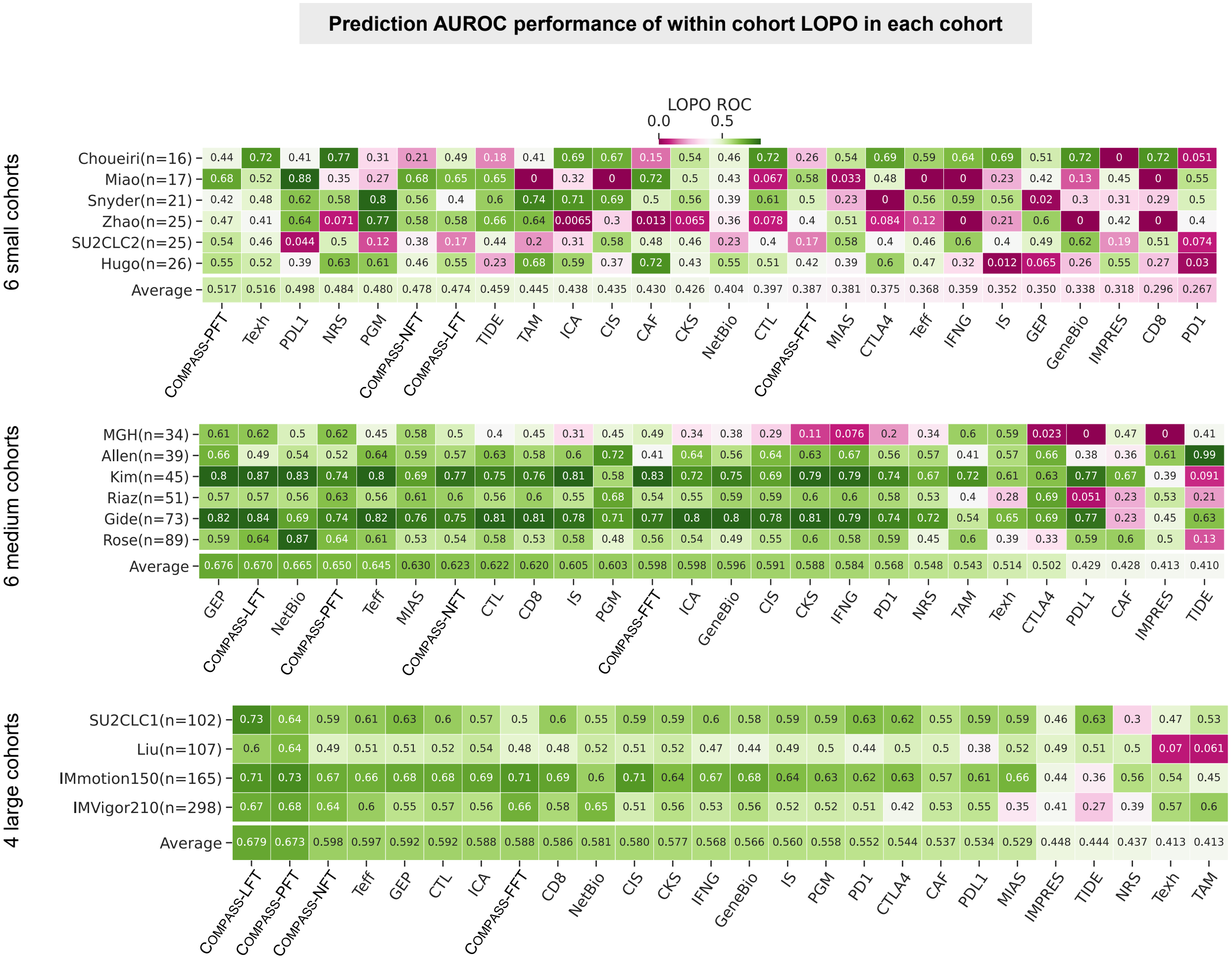


**Supplementary Fig. S6: Prediction AUROC performance in within-cohort LOPO across cohorts.** The heatmap shows the AUROC performance for various methods across different cohorts using within-cohort leave-one-patient-out (LOPO) cross-validation. Darker green shades represent higher AUROC values (better performance), while pink shades indicate lower AUROC values (worse performance). On average, small cohorts show lower performance across all methods.

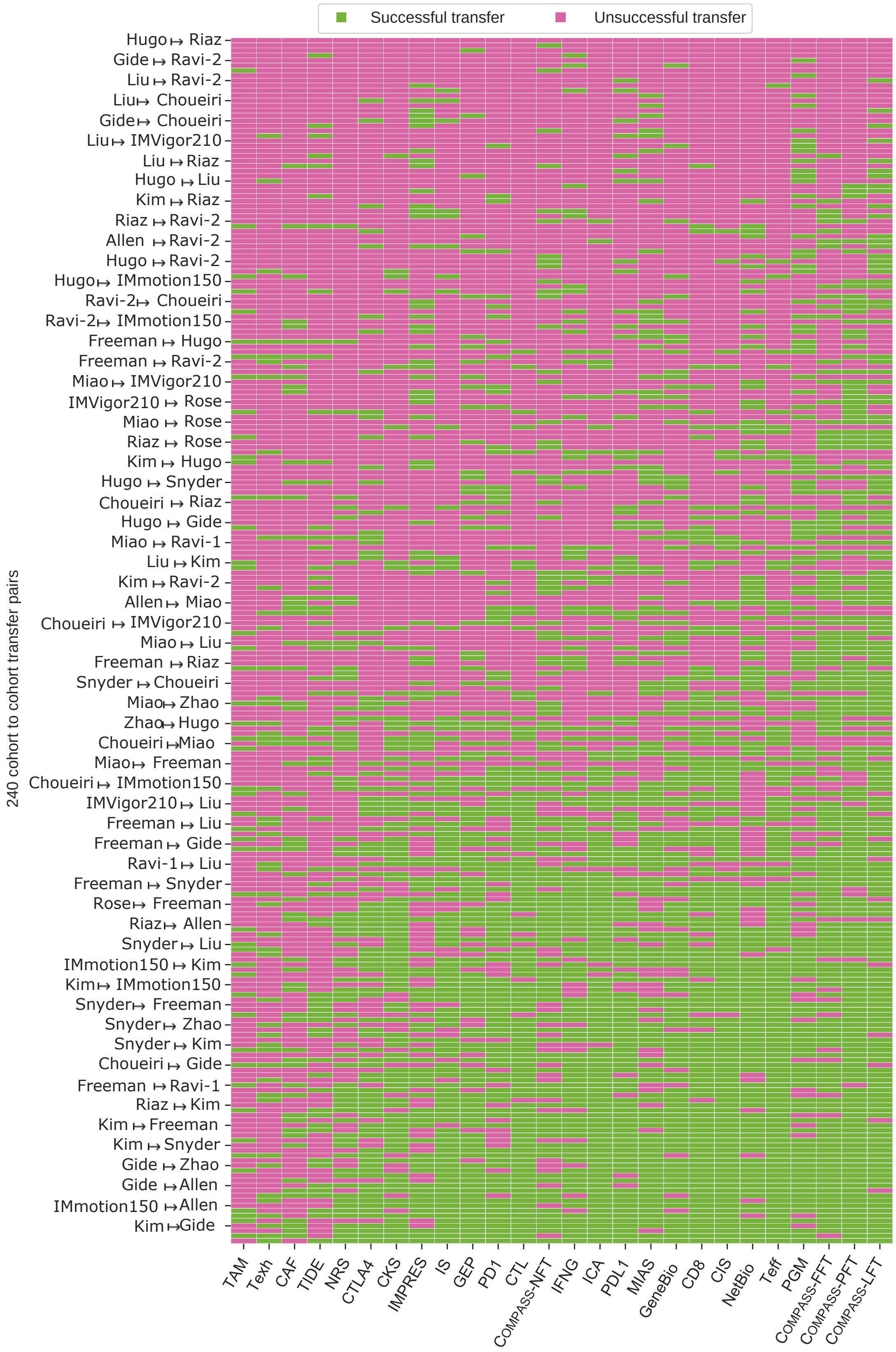


**Supplementary Fig. S7: Success and failure in cohort-to-cohort transfer analysis.** Heatmap showing transferability across 240 unique cohort pairs (16 × 15). Green bars denote successful transfers (model accuracy exceeds reference accuracy when trained on source cohort and tested on target cohort), while pink bars indicate unsuccessful transfers (accuracy below reference). Rows represent cohort pairs, and columns correspond to evaluated transfer methods.

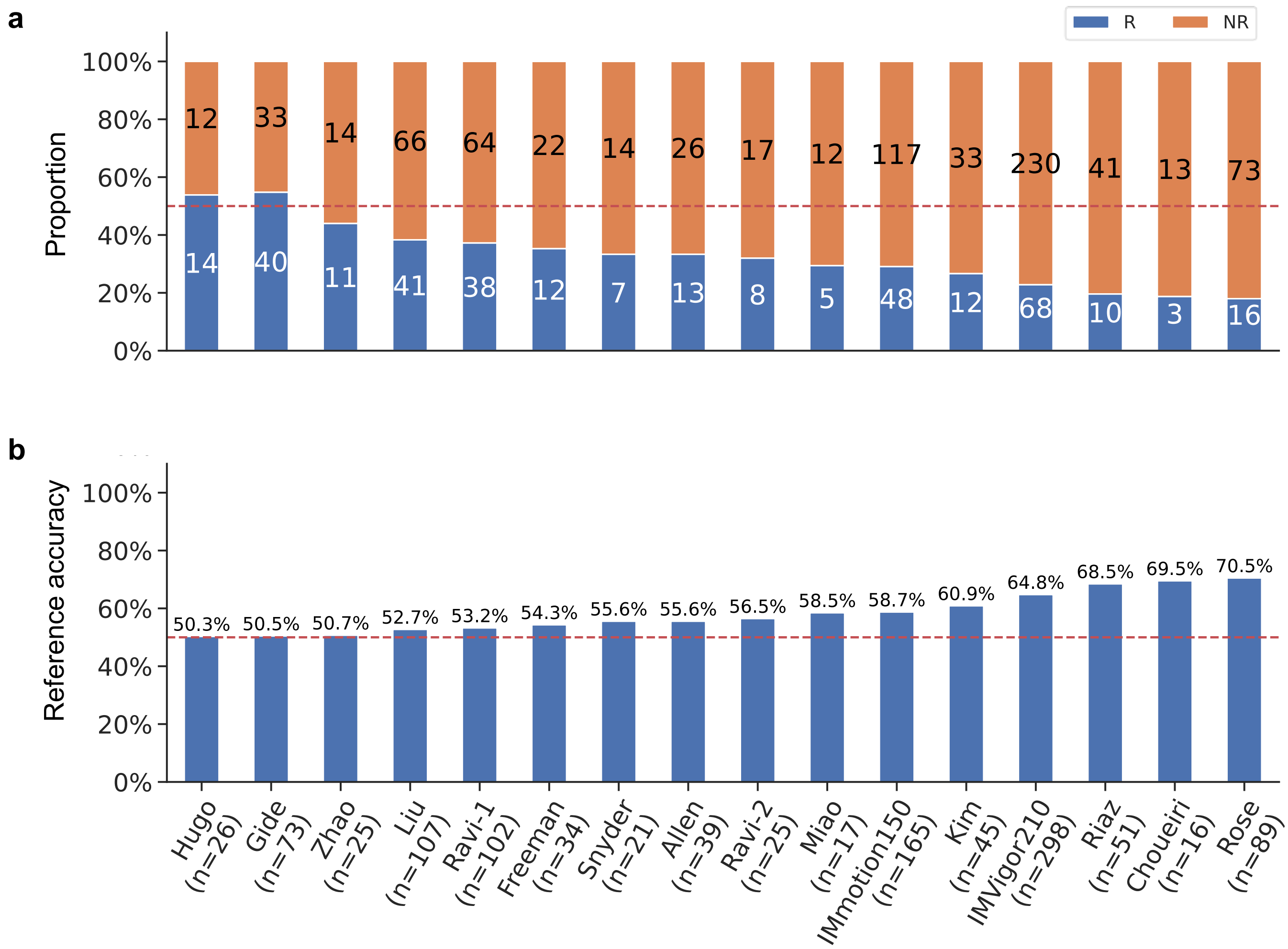


**Supplementary Fig. S8: Label distribution and reference accuracy across cohorts.** **(a)** Class distribution (Responders [R, blue] vs. Non-Responders [NR, orange]) for all cohorts. Sample sizes are annotated above bars; the dashed red line indicates balanced class proportions (50%). **(b)** Reference accuracy values were calculated based on each cohort's class distribution, representing expected random performance. Higher accuracy in imbalanced cohorts reflects bias toward the majority class.

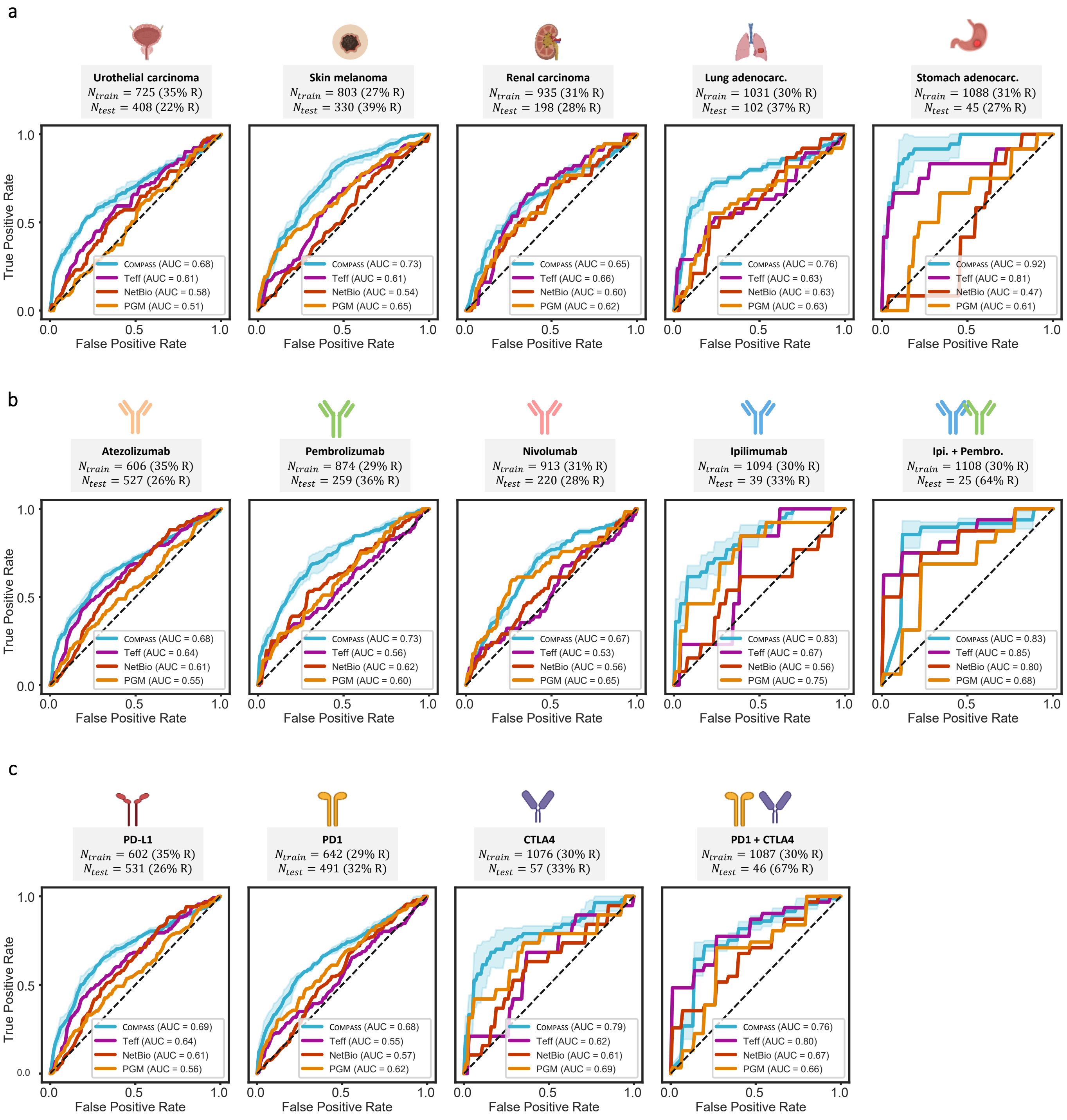


**Supplementary Fig. S9: ROC curves for response prediction stratified by cancer type, drug, and immune checkpoint target.** Receiver operating characteristic (ROC) curves comparing COMPASS-PFT, Teff, NetBio, and PGM models across multiple biological groupings. Icons sourced from BioRender.com. **(a)** cancer types including urothelial carcinoma, skin melanoma, renal carcinoma, lung adenocarcinoma, and stomach adenocarcinoma. **(b)** ICI drugs including atezolizumab, pembrolizumab, nivolumab, ipilimumab, and the combination of ipilimumab and pembrolizumab. **(c)** immune checkpoint targets including PD-L1, PD1, CTLA4, and PD1+CTLA4. Numbers of training ($N_{\text{train}}$) and test ($N_{\text{test}}$) patients, with response rates (R), are indicated in each panel. Lines show mean ROC curves with shaded bands representing ± 1 standard deviation across three repeats using different random seeds. COMPASS consistently achieves higher AUROC across diverse stratifications. Performance in other evaluation metrics is shown in **Fig. 3**.

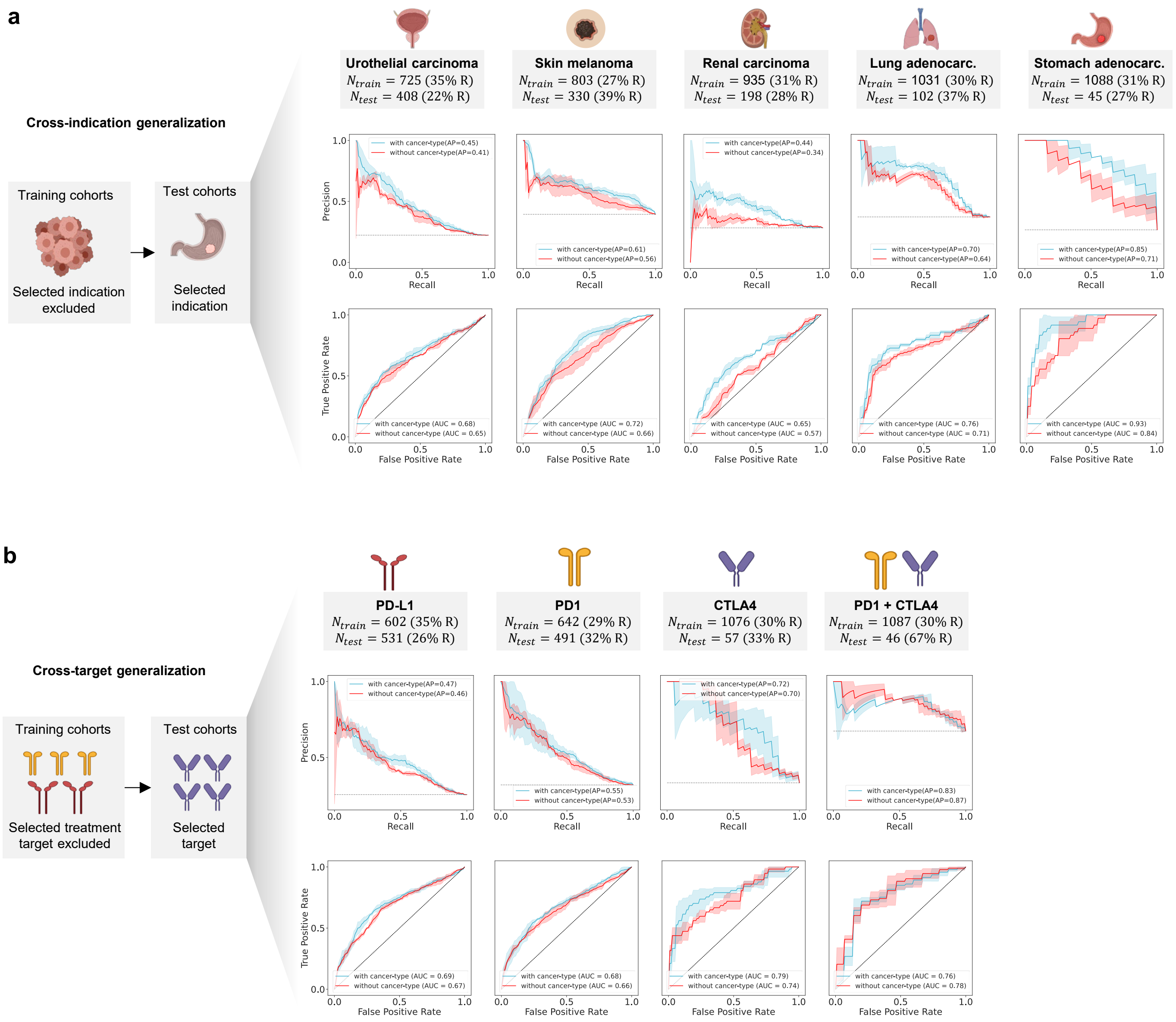


**Supplementary Fig. S10: Ablation of the cancer-type token during fine-tuning.** To determine whether predictive performance is driven by cancer-type-specific response prevalence encoded in the cancer-type token, we fine-tuned COMPASS either with or without this token and evaluated generalization performance. Details are provided in **Supplementary Methods Method S5**. Icons sourced from BioRender.com. **(a)** Cross-indication generalization: models were trained after excluding all cohorts corresponding to the indicated cancer type and evaluated on the held-out cohorts of that cancer type (leave-one-indication-out). Precision-recall curves (top) and ROC curves (bottom) are shown for urothelial carcinoma, melanoma, renal carcinoma, lung adenocarcinoma, and stomach adenocarcinoma. Blue curves denote models including the cancer-type token; red curves denote models without it. **(b)** Cross-target generalization: models were trained after excluding all cohorts corresponding to the indicated treatment target and evaluated on the held-out cohorts for that target (leave-one-target-out). Precision-recall (top) and ROC (bottom) curves are shown for PD-L1, PD-1, CTLA-4, and PD-1+CTLA-4. Blue curves denote models including the cancer-type token; red curves denote models without it. Lines show mean values with shaded bands representing ± 1 standard deviation across three repeats using different random seeds.

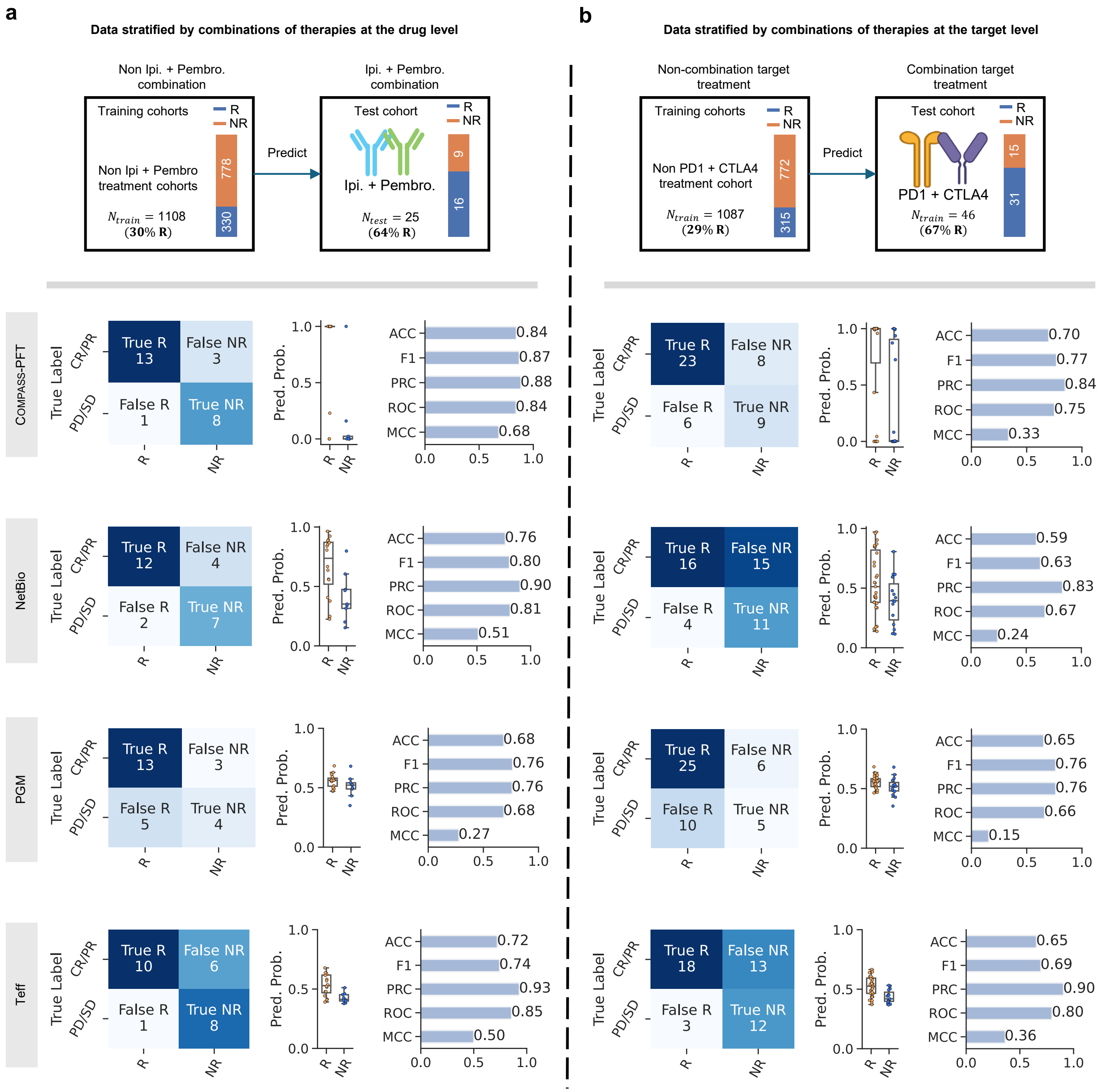


**Supplementary Fig. S11: Prediction performance stratified by combinations of therapies at both drug and target levels.** Icons sourced from BioRender.com. **(a)** The left panel presents data stratified by drug combinations (Ipi and Pembro), with the test cohort ($N_{test} = 25$) receiving Ipi and Pembro treatment and the training cohort without combination therapy ($N_{train} = 1108$). Four methods are compared: PFT, NetBio, PGM, and Teff. Each method's performance is evaluated using ACC, F1, PRC, ROC, and MCC metrics. Confusion matrices show true responders (R) and non-responders (NR), with predicted probabilities plotted on the right. **(b)** The right panel displays data stratified by combination therapies targeting PD1 and CTLA4, with the test cohort($N_{test} = 46$) receiving PD1 and CTLA4 combination therapy and the training cohort treated with single-target agents ($N_{train} = 1087$). Performance metrics (ACC, F1, PRC, ROC, MCC) for the same four methods are shown. True and false positives/negatives are presented in confusion matrices, with predicted probabilities on the right. Despite the imbalance in the proportion of responders dominating the test set, which differs from the distribution in the training set, all models demonstrate strong predictive capabilities for combination therapy response. This indicates that the models' performance is not affected by the imbalance in positive and negative samples in the training set.

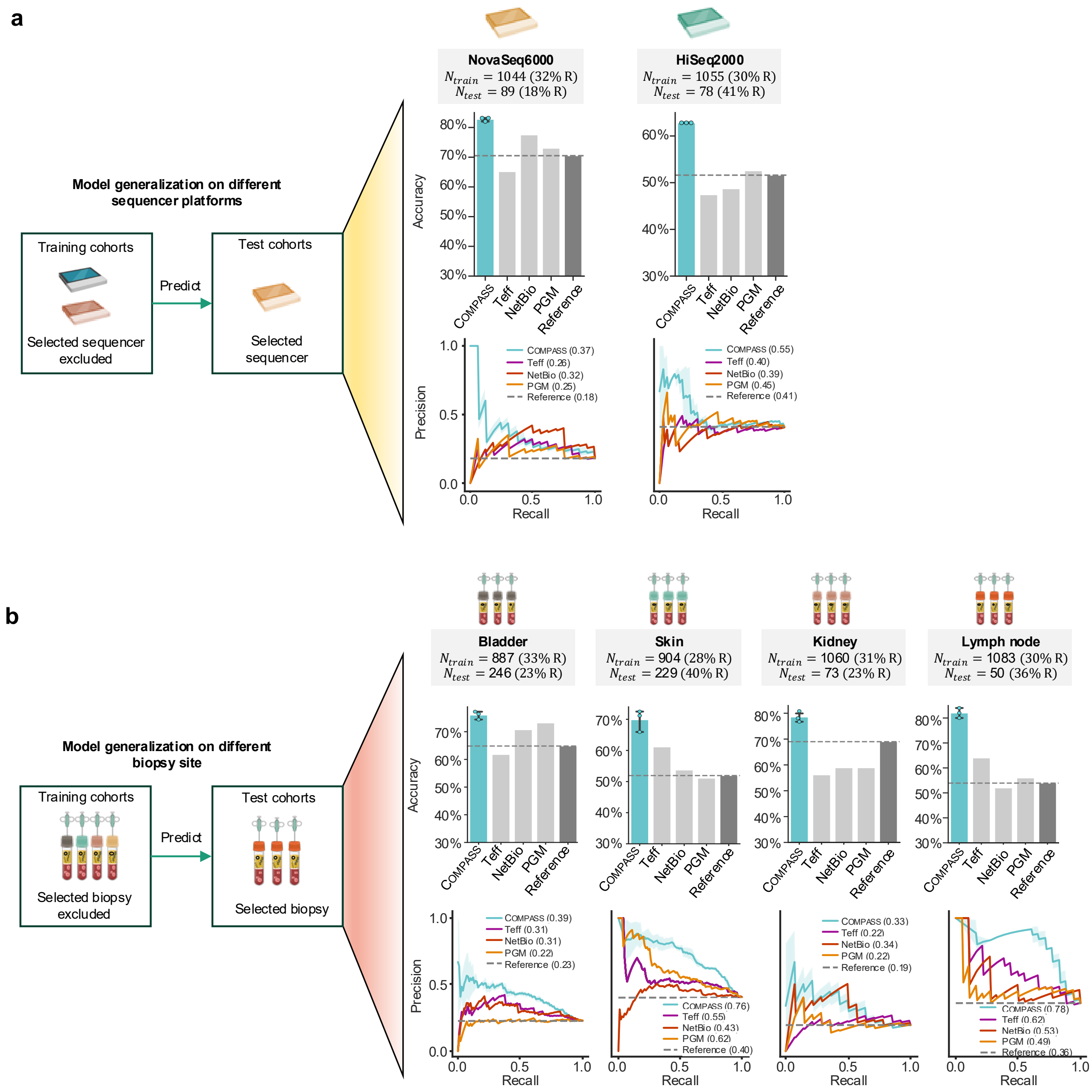


**Supplementary Fig. S12: Evaluation of model robustness across different sequencing platforms and biopsy sites.** To assess the robustness of the COMPASS-PFT model and compare it with better-performing baseline methods such as Teff, NetBio, PGM, and reference performance, we also evaluated the models across different sequencing platforms and biopsy sites. Each evaluation involved training the models on cohorts excluding a specific category and then testing on the excluded cohort. Training size ($N_{train}$) and test size ($N_{test}$) are indicated. The upper bar plots show mean accuracy (%) with error bars representing 95% confidence intervals. The lower line plots show mean precision–recall curves with shaded bands representing ± 1 standard deviation across three repeats using different random seeds. Icons sourced from BioRender.com. **(a)** Model robustness on different sequencing platforms. Shown is the evaluation of robustness across different sequencing technologies. Models were trained on cohorts excluding those processed by a specific sequencing platform and then tested on the excluded platform cohort. For example, the HiSeq2000 cohort is excluded during training and used for testing. The results display prediction accuracy and precision-recall curves for NovaSeq6000 and HiSeq2000 platforms. **(b)** Model robustness on different biopsy sites. Shown is the evaluation of robustness across different biopsy sites. Models were trained on cohorts excluding those from a specific biopsy site and then tested on the excluded biopsy site cohort. For example, the lymph node biopsy cohort is excluded during training and used for testing. The results display prediction accuracy and precision-recall curves for different biopsy sites: bladder, skin, kidney, lymph node.

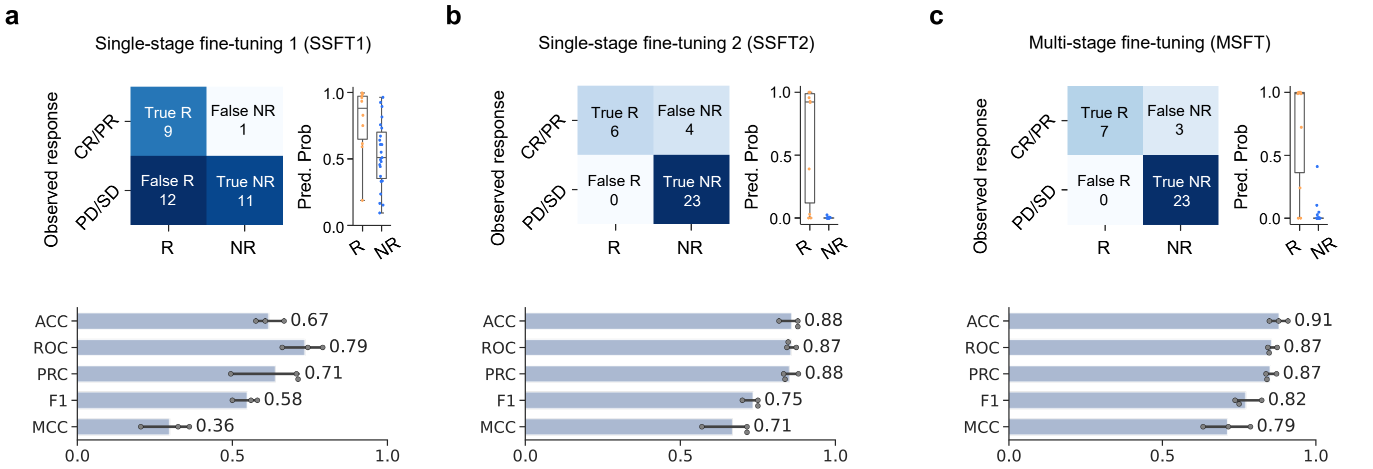


**Supplementary Fig. S13: Comparison of single-stage and multi-stage fine-tuning strategies for LUAD-specific immunotherapy response prediction.** Shown are evaluation results for three fine-tuning strategies applied to a pembrolizumab-treated LUAD cohort ($n$ = 33), using models pre-trained on TCGA pan-cancer data ($n$ = 10,184). **(a)** Performance of SSFT1, where the model was fine-tuned on a small LUAD cohort without pembrolizumab treatment ($n$ = 69) using linear probing. Displayed are the predicted response probability distribution, confusion matrix, and performance metrics including accuracy (ACC), ROC-AUC, PR-AUC, F1-score, and MCC. **(b)** Performance of SSFT2, where the model was fine-tuned on a pan-cancer cohort excluding LUAD ($n$ = 1,031) using partial fine-tuning. Shown are the same evaluation components as in (a). **(c)** Performance of MSFT, involving sequential fine-tuning first on the pan-cancer cohort ($n$ = 1,031) and subsequently on the no-pembrolizumab LUAD cohort ($n$ = 69). This approach demonstrates progressive model adaptation to the distinct characteristics of the LUAD cohorts, emphasizing tailored fine-tuning for these specialized groups. Bars show mean values with error bars representing 95% confidence intervals across three repeats using different random seeds. MSFT achieves superior separation between responders and non-responders, lower false-positive rates, and improved metrics compared to single-stage strategies.

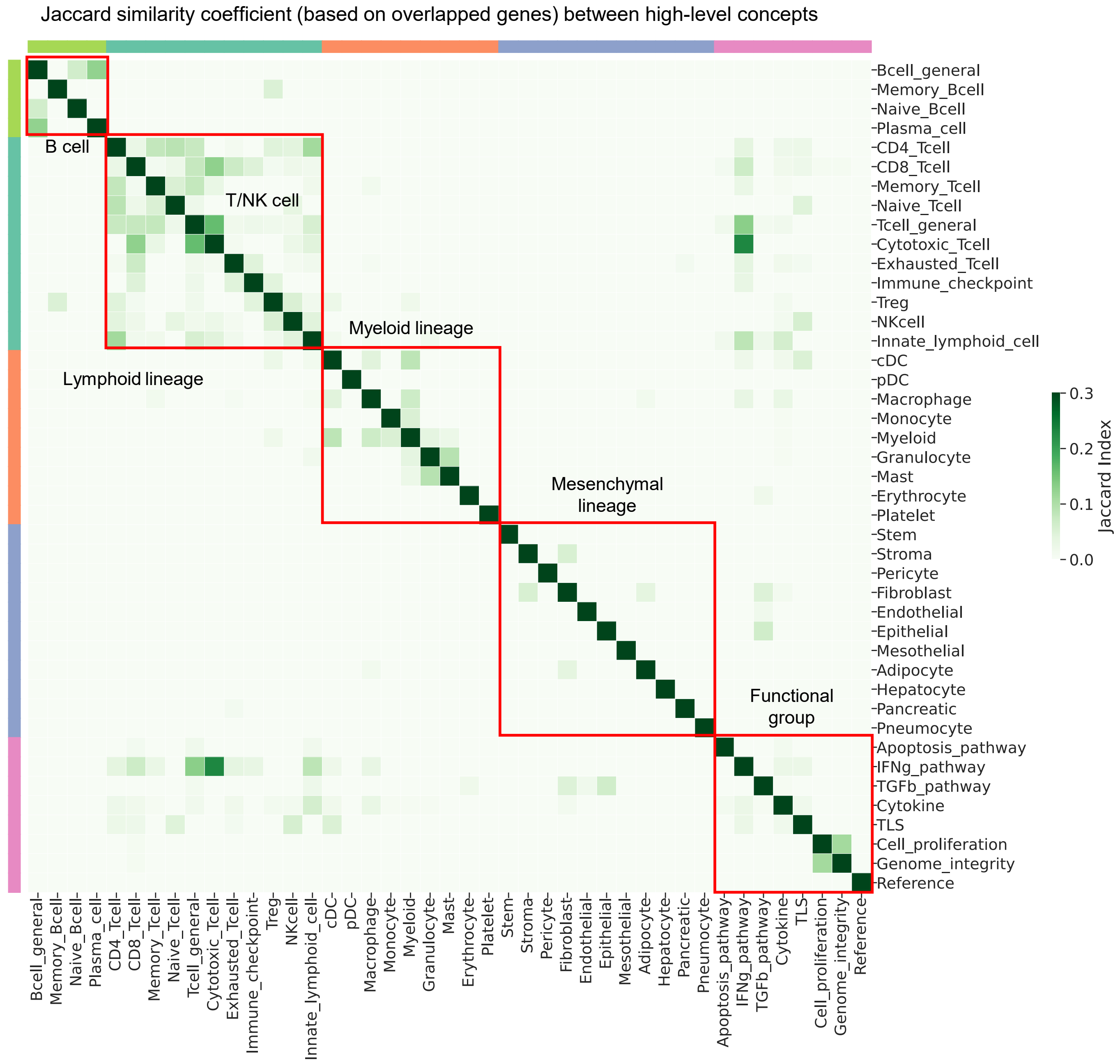


**Supplementary Fig. S14: Jaccard similarity coefficient between 43 concepts based on overlapping genes.** The Jaccard index between two concepts is calculated as the ratio of the overlapping genes to the union of genes associated with each concept. This heatmap visualizes the Jaccard similarity index among 43 high-level concepts utilized in COMPASS, based on their overlapping genes. The matrix is color-coded, where darker green shades represent higher similarity between concepts, and lighter shades indicate lower similarity. The concepts are categorized into broad groups, such as B-cell lineage, T/NK cell lineage, Myeloid lineage, Mesenchymal lineage, and Functional pathways, which are outlined with red boxes.

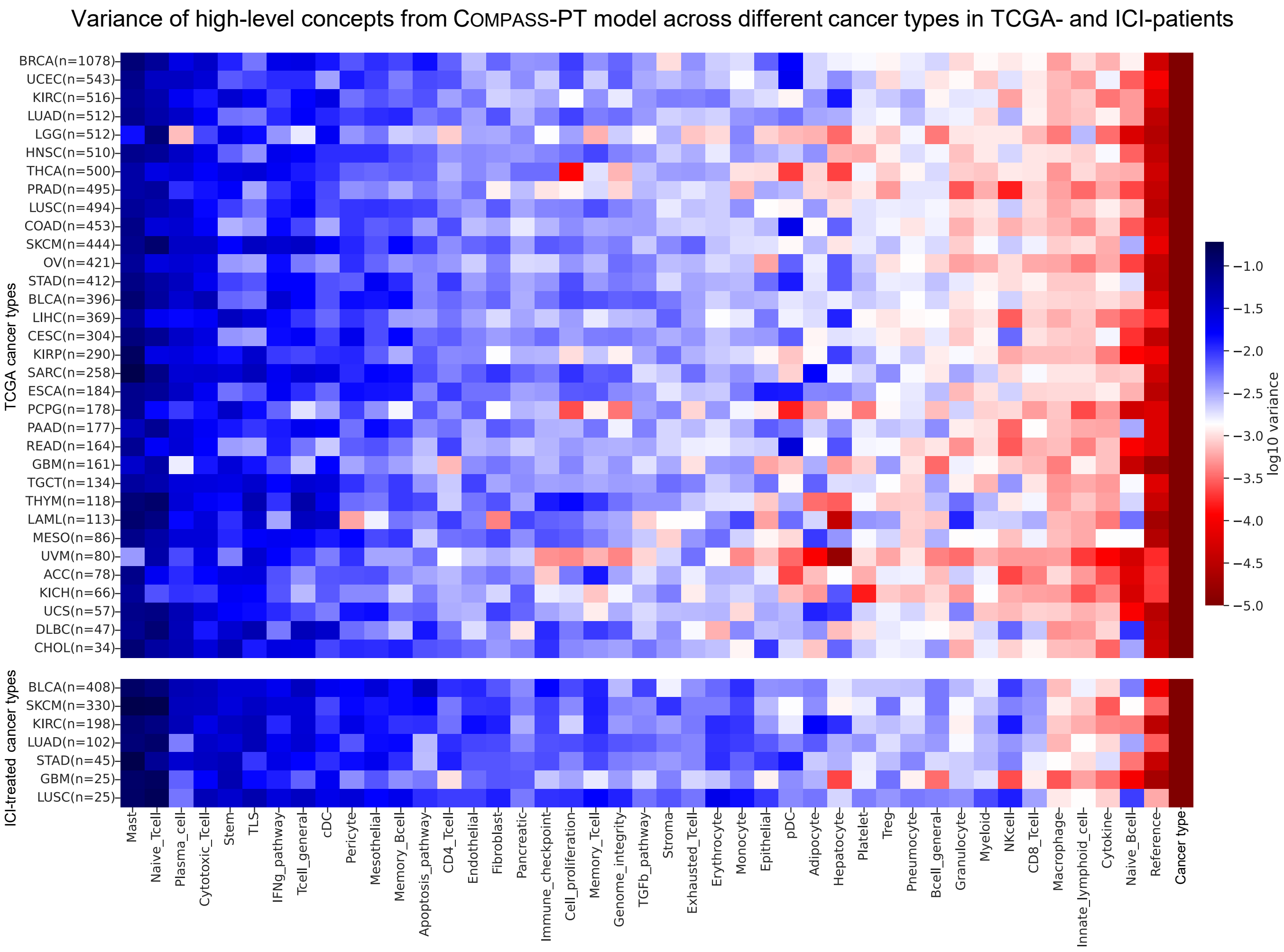


**Supplementary Fig. S15: Variation of 44 concepts from COMPASS-PT model across different cancer types in TCGA and ICI patients.** This heatmap displays the $\log_{10}$-transformed variance of 44 high-level concepts derived from the pre-trained model COMPASS-PT across 33 TCGA cancer types and 7 ICI-treated cancer cohorts. Blue shades indicate higher variance, while red shades represent lower variance across cancer types. Concepts such as `Reference` and `Cancer Type` consistently show the lowest variance within each cancer type, indicating that these concepts are highly stable and less variable across patients within the same cancer type. This may be due to the nature of the `Reference` concept serving as a baseline or control, and `Cancer Type` representing common cancer-specific traits that do not vary as much within a particular cancer type. In contrast, other immune and stromal concepts show higher variance, reflecting the diversity of immune responses and tumor microenvironments across different patients and cancer types.

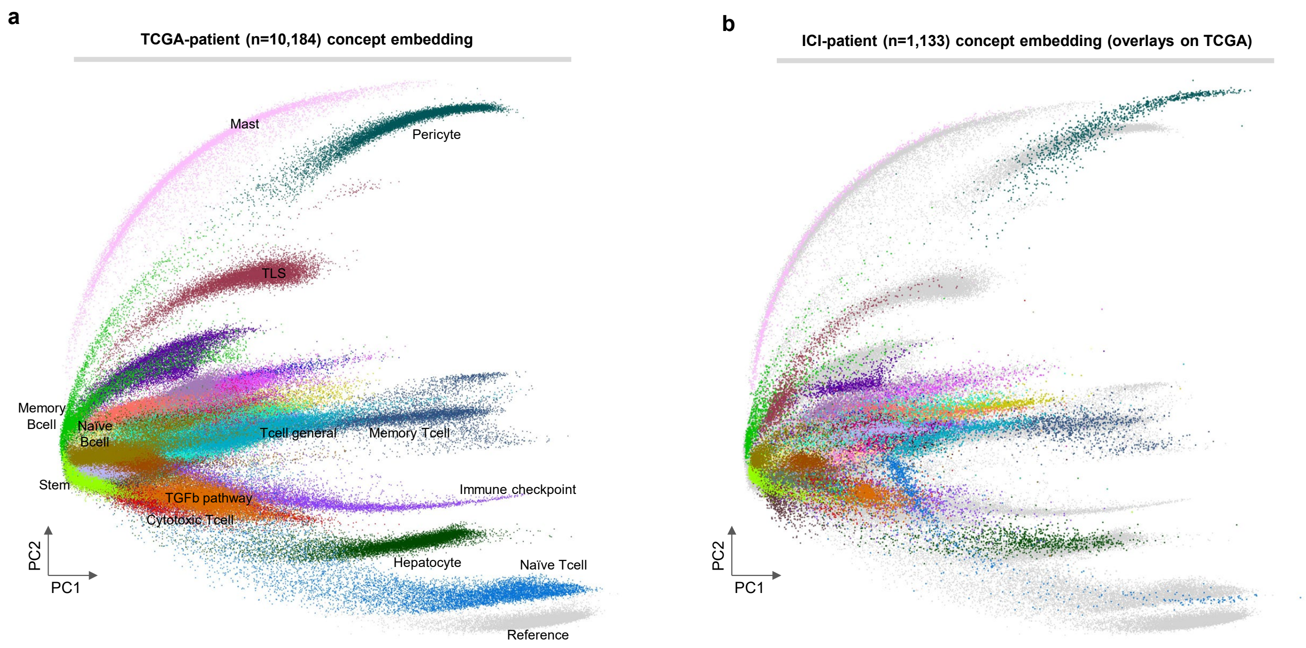


**Supplementary Fig. S16: PCA embedding of TCGA/ICI-patient concepts.** Shown are the principal component analysis (PCA) 2D embeddings of the 43 concepts (each concept represented by a 32-dimensional vector). The panel **(a)** shows the 2D embedding based on TCGA patients ($n$ = 10,184), and the right panel**(b)** overlays the concept embeddings of ICI-treated patients ($n$ = 1,133) onto the TCGA embeddings. This visualization demonstrates the extent to which ICI-treated patients align with or diverge from the broader TCGA patient cohort in terms of immune and stromal concept profiles.

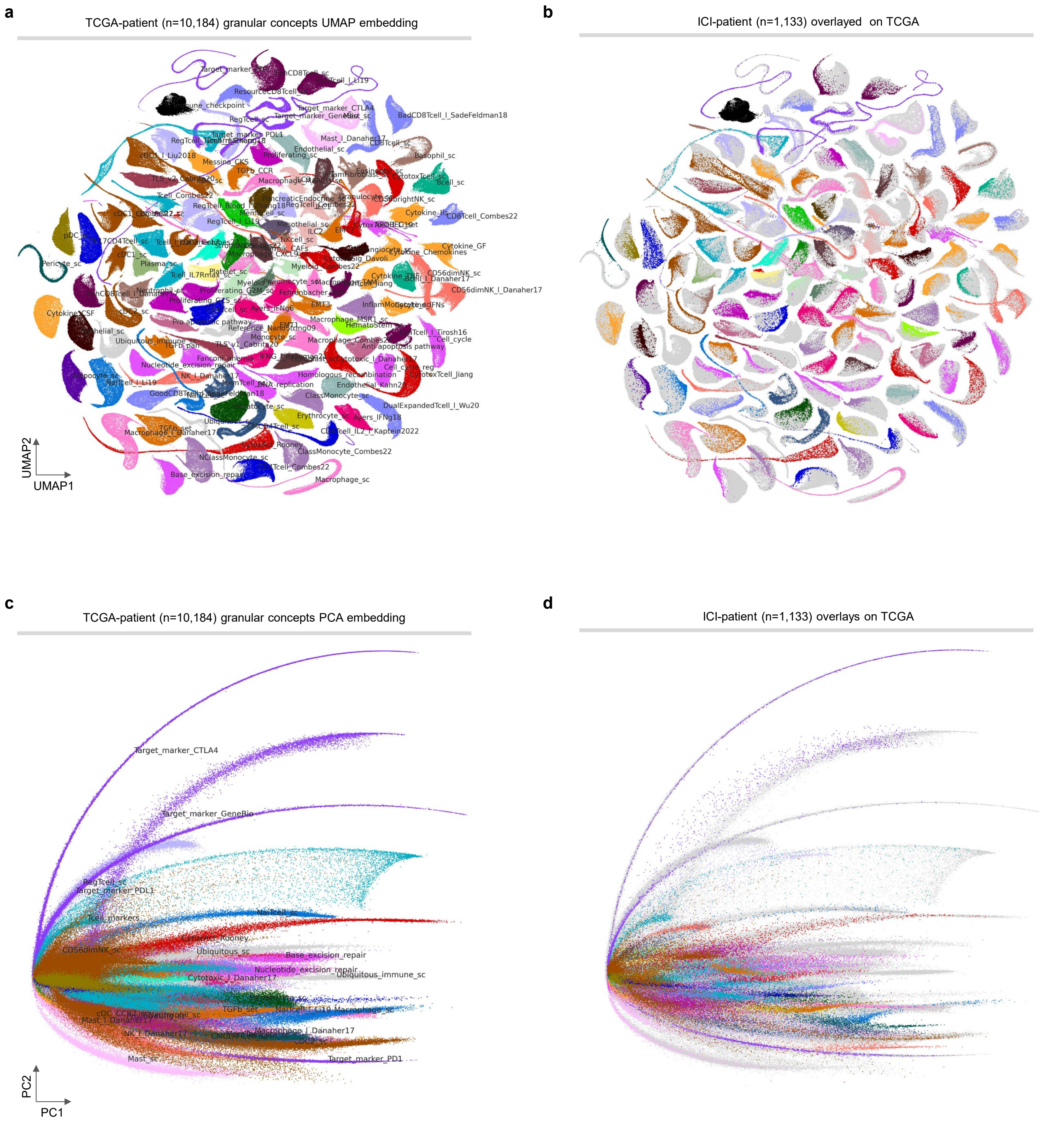


**Supplementary Fig. S17: Granular concepts embeddings of TCGA patients and ICI-patients visualized by UMAP and PCA.** Shown are the 2D embeddings of the 132 granular concepts (each granular gene set concept represented by a 32-dimensional vector). Each point in the figure is a single patient. Different colors correspond to different concepts learned by COMPASS. **(a)** UMAP embedding of granular concepts from TCGA patients ($n$ = 10,184), showing how COMPASS organizes the learned granular concepts in a reduced-dimensionality space. **(b)** UMAP embedding of granular concepts from ICI patients ($n$ = 1,133), overlayed on top of the TCGA embedding, highlighting the similarity or divergence of granular concepts between the ICI patients and broader TCGA dataset. **(c)** PCA embedding of granular concepts from TCGA patients, illustrating their spread across principal component space. **(d)** PCA embedding of granular concepts from ICI patients, superimposed on the TCGA granular concepts PCA space. This shows how granular gene set representations in ICI patients overlap or deviate from the broader TCGA embedding.

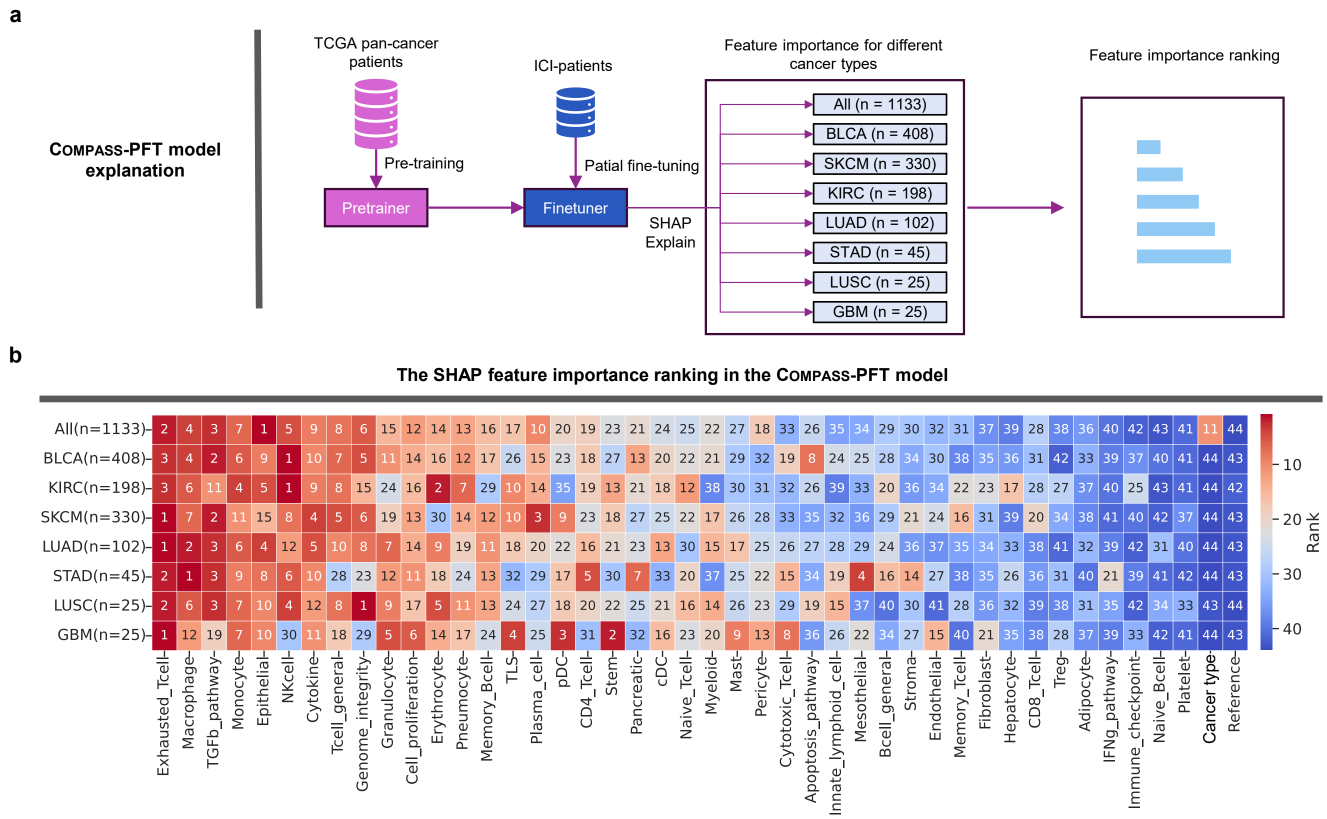


**Supplementary Fig. S18: Feature importance analysis in the COMPASS-PFT for predicting immunotherapy response. (a)** Feature importance calculation in the COMPASS-PFT model. The COMPASS-PFT model was first fine-tuned on the ICI cohorts (n = 1133), then the KernalSHAP method was employed to quantify the importance of each of the 44 concepts, comparing their influence across all patients and specific cancer cohorts (BLCA, KIRC, SKCM, LUAD, STAD, LUSC, GBM). **(b)** Concept feature rankings in COMPASS-PFT model. These heatmaps display the rankings of the 44 concepts, ordered by their mean importance scores from left to right. Higher-ranked concepts play a more critical role in the model's ability to predict the response to immunotherapy.

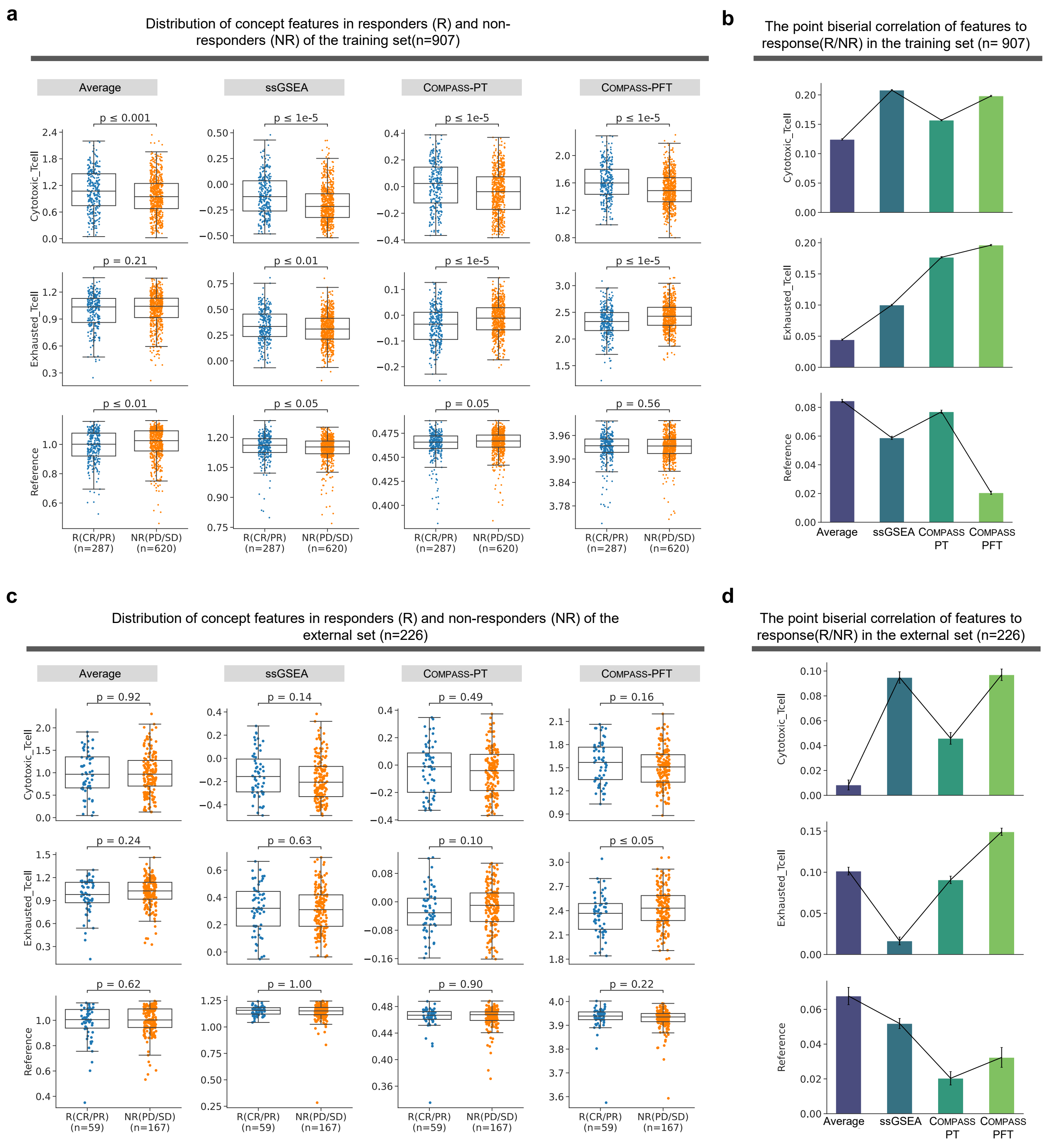


**Supplementary Fig. S19: Comparison of concept scoring methods in distinguishing responders from non-responders. (a)** Distributions and statistical significance of concept scores between responders (R) and non-responders (NR) across different scoring methods (geometric average, ssGSEA, pre-trained model COMPASS-PT, and fine-tuned model COMPASS-PFT) in the training set ($n$ = 907). Shown are three representative concepts—`Cytotoxic T Cell`, `Exhausted T Cell`, and `Reference`. Significance was assessed using the Mann–Whitney U test, with corresponding $p$-values indicated. **(b)** Point biserial correlations between concept scores and binary response labels in the training set. The strength of association is shown for the same three concepts across scoring methods, reflecting their predictive utility. **(c)** Distribution of concept scores for responders and non-responders in the external validation set ($n$ = 226), assessing the robustness of observed patterns. **(d)** Point biserial correlations in the external validation set, evaluating how well each scoring method generalizes beyond the training cohort.

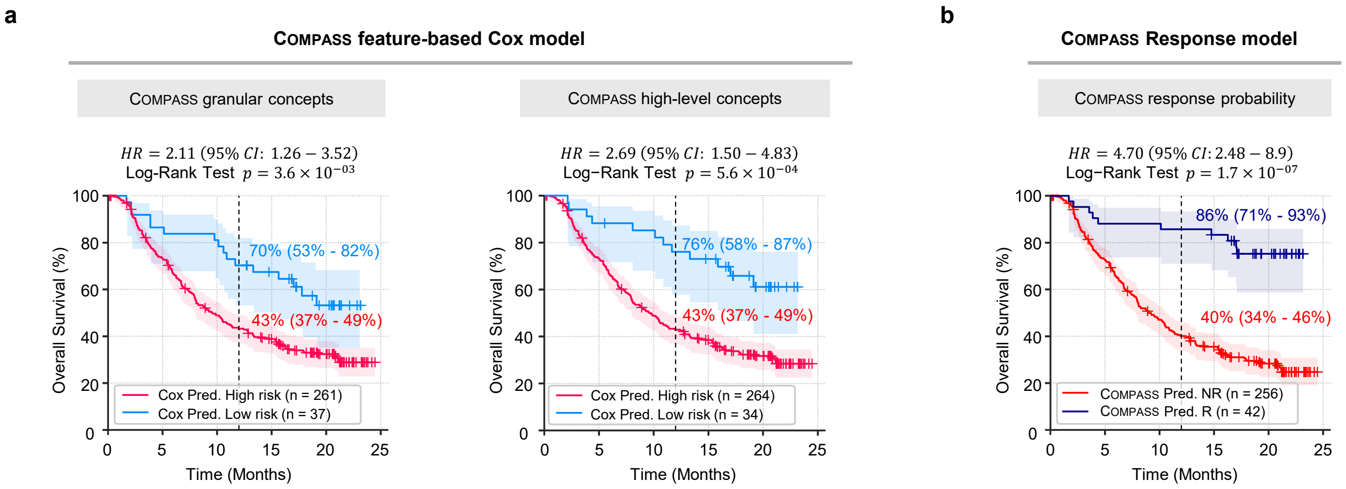


**Supplementary Fig. S20: Kaplan-Meier survival curves stratified by risk groups or response predictions using the COMPASS model in the IMvigor210 cohort.** Log-rank test p-values, and Cox proportional hazard ratios (HRs) for both feature-based and response-based analyses were calculated. Shaded bands represent 95% confidence intervals of the Kaplan–Meier survival estimates. **(a)** COMPASS feature-based Cox models: Ridge Cox regression models were trained on COMPASS-PFT derived features of 132 granular concepts and 44 high-level concepts to predict patient risk scores. Patients were stratified into high-risk (red) and low-risk (blue) groups based on the top 10% risk score cutoff values from the training set. Significant differences (log-rank test $p$)in overall survival were observed between high-risk and low-risk groups are shown. **(b)** COMPASS response model: Patients were stratified into responders ($P_R \geq 0.5$, blue) and non-responders ($P_R < 0.5$, red) based on response probabilities predicted by the COMPASS model. Responders exhibited significantly longer overall survival than non-responders (log-rank test $p = 1.7 \times 10^{-7}$). These results demonstrate the predictive power of COMPASS-derived features and response probabilities in stratifying patients by survival.

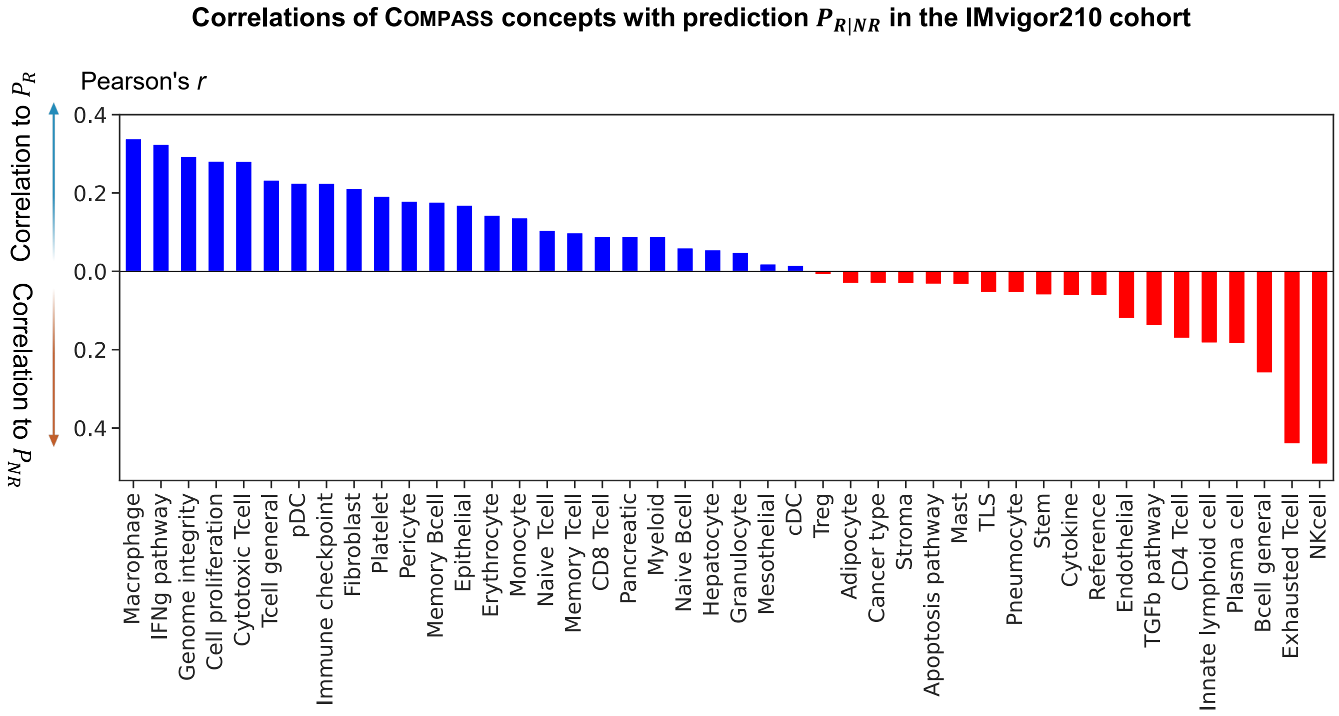


**Supplementary Fig. S21: Ranking of COMPASS high-level concepts by Pearson's correlation with response prediction in the IMvigor210 cohort.** This figure presents the correlation coefficients between each of the 44 high-level COMPASS concept scores and predicted response probabilities ($P_{(R|NR)}$), calculated using the COMPASS-PFT model trained on all ICI cohorts except IMvigor210 (leave-one-cohort-out approach). Concepts positively correlated with response ($P_R$) are depicted in blue, indicating their beneficial association with responders (R). In contrast, concepts positively correlated with non-response ($P_{NR}$) are displayed in red, underscoring their link with non-responders (NR). This ranking highlights the differential impact of each concept on predicting patient outcomes in immunotherapy.

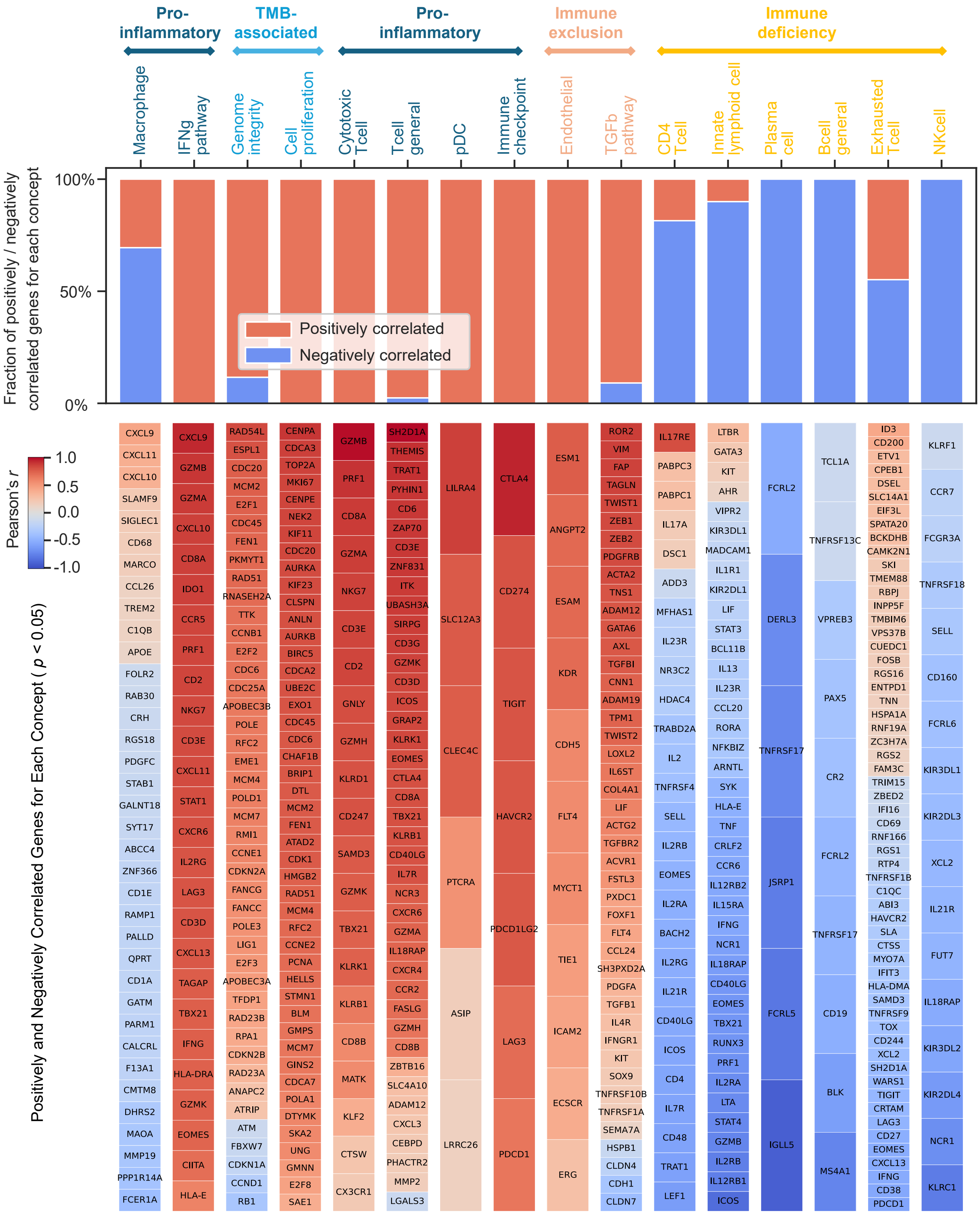


**Supplementary Fig. S22: Correlation of genes with COMPASS concept features in the IMvigor210 cohort.** The top panel illustrates the fraction of significantly correlated genes ($p < 0.05$) for each COMPASS concept, categorized as either positively (red) or negatively (blue) correlated. The bottom panel ranks these significant genes based on their Pearson correlation coefficients ($r$) with the corresponding concept feature. The correlations were computed by comparing COMPASS concept feature scores with gene expression levels, highlighting key genes that are strongly associated with each concept. These top-ranked 16 concepts can be grouped into Pro-inflammatory, TMB-associated, Immune exclusion, and Immune deficiency categories based on their correlated genes.

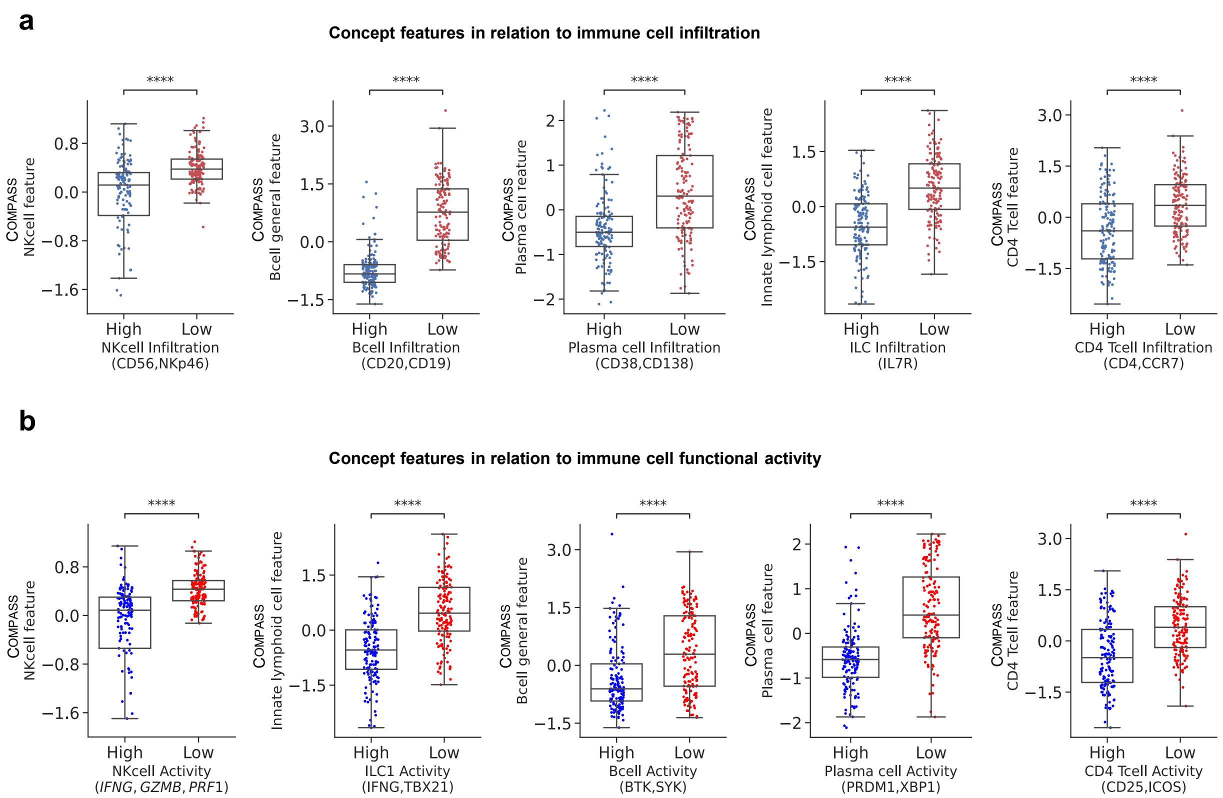


**Supplementary Fig. S23: COMPASS concept features that positively-correlated to $P_{NR}$ in relation to immune cell infiltration and functional activity.** Shown are the non-responder-associated concept features of `NK Cell`, `B Cell General`, `Plasma Cell`, `Innate Lymphoid Cell`, and `CD4 T Cell` in relation to **(a)** immune cell infiltration and **(b)** immune cell functional activity. The results indicate that for these concept features, COMPASS captures a distinct inverse relationship with immune cell infiltration and functional activity. The y-axis represents the COMPASS concept feature z-score, while the x-axis categorizes samples into high ($n = 149$) and low ($n = 149$) infiltration or activity groups. These groups are defined based on the ssGSEA signature scores of selected marker genes, with high-infiltration or high-activity corresponding to scores above the median and low-infiltration or low-activity corresponding to scores below the median. Statistical comparisons were performed using two-sided Mann-Whitney U tests with no adjustment for multiple comparisons. Exact $p$-values for high vs. low group comparisons are as follows — **(a)** infiltration: NK cell $p = 2.247 \times 10^{-11}$, B cell $p = 8.992 \times 10^{-42}$, Plasma cell $p = 1.878 \times 10^{-13}$, Innate lymphoid cell $p = 9.633 \times 10^{-20}$, CD4 T cell $p = 6.600 \times 10^{-10}$; **(b)** functional activity: NK cell $p = 2.639 \times 10^{-18}$, Innate lymphoid cell $p = 2.911 \times 10^{-19}$, B cell $p = 1.457 \times 10^{-9}$, Plasma cell $p = 2.553 \times 10^{-25}$, CD4 T cell $p = 1.384 \times 10^{-12}$.

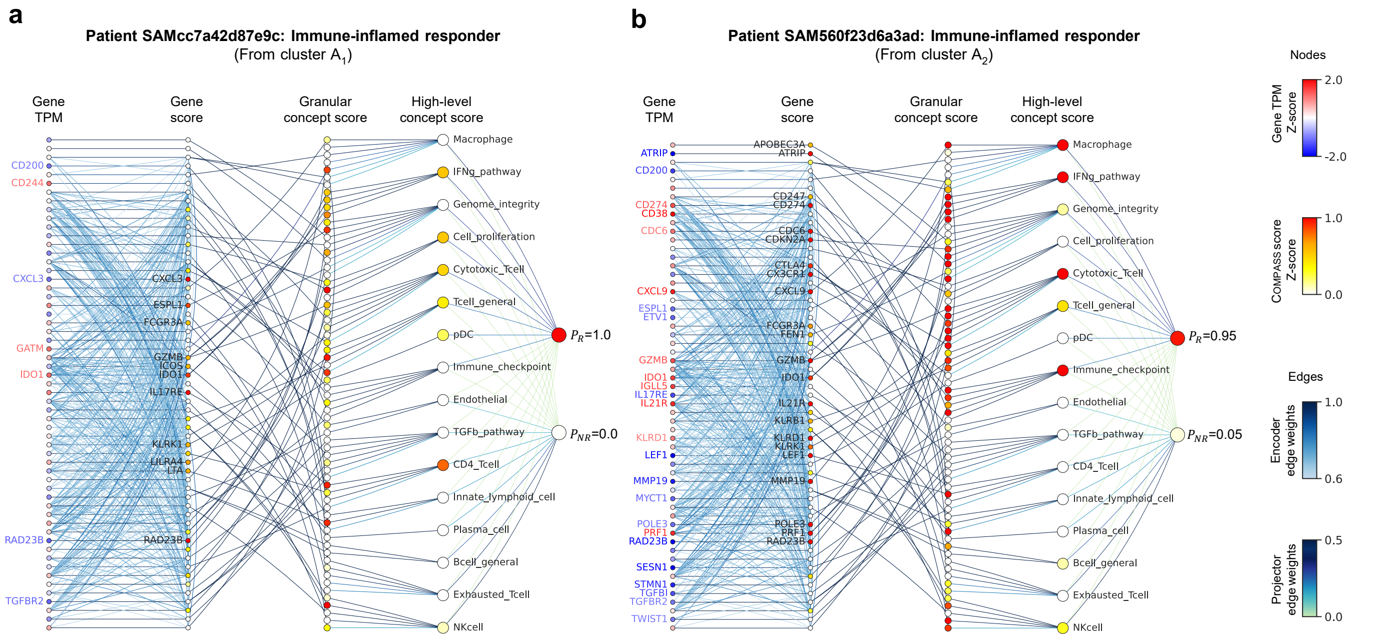


**Supplementary Fig. S24: Personalized response maps for two representative immune-inflamed responders.** Shown are two representative patients from clusters $A_1$ and $A_2$ within the immune-inflamed responder group, as identified in the **Extended Data Fig. 5**. The representative patient from cluster $A_3$ is shown in **Fig. 6a**. **(a)** Response map for patient SAMcc7a42d87e9c (Cluster $A_1$). **(b)** Response map for patient SAM560f23d6a3ad (Cluster $A_2$).

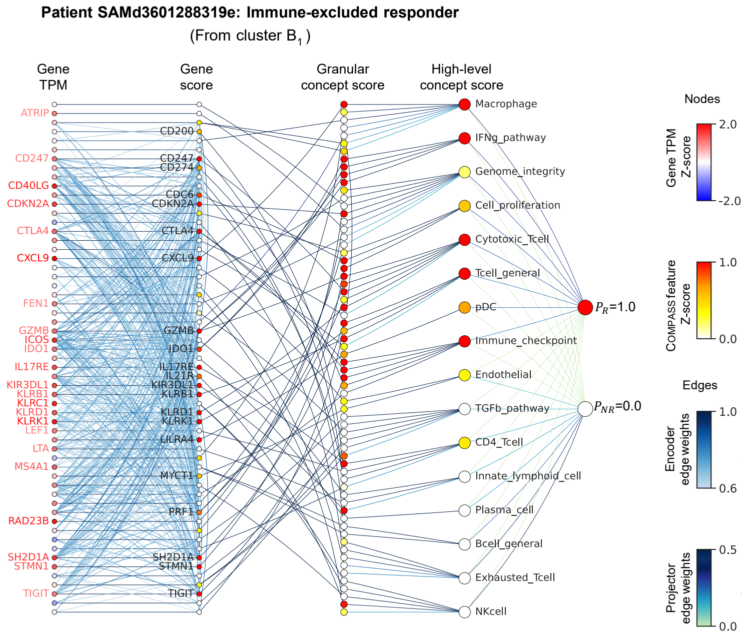


**Supplementary Fig. S25: Personalized response map for a representative immune non-inflamed responder.** Shown is patient SAMd3601288319 from cluster $B_1$ of the immune non-inflamed responder group. A representative patient from cluster $B_2$ is shown in **Fig. 6b**.

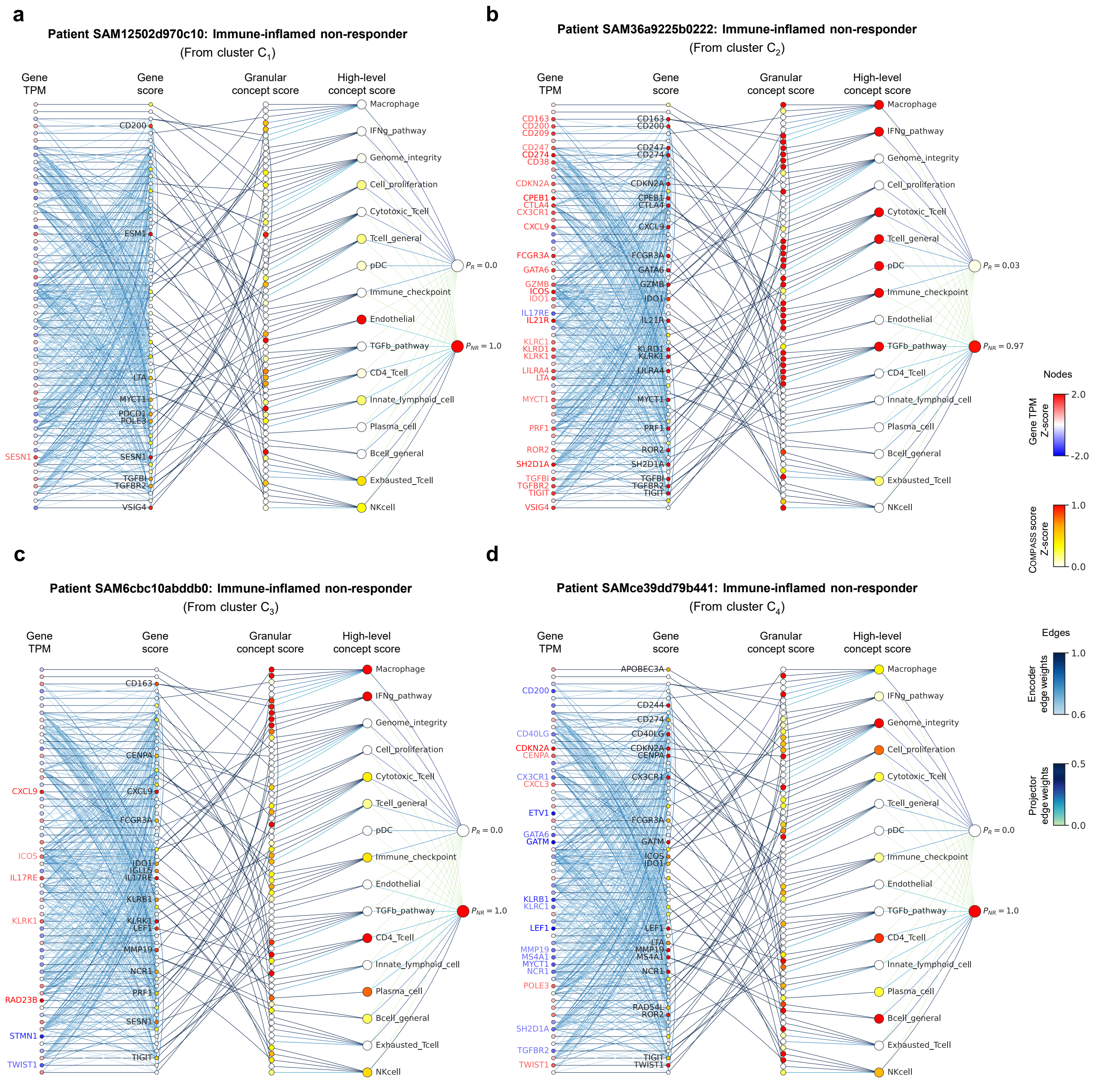


**Supplementary Fig. S26: Personalized response maps for four representative immune-inflamed non-responders.** Shown are four representative patients from clusters $C_1$–$C_4$ within the immune-inflamed non-responder group. The representative patient from cluster $C_5$ is presented in **Fig. 6c**. **(a)** Response map for patient SAM12502d970c10 (Cluster $C_1$). **(b)** Response map for patient SAM36a9225b0222 (Cluster $C_2$). **(c)** Response map for patient SAM6cbc10abddb0 (Cluster $C_3$). **(d)** Response map for patient SAMce39dd79b441 (Cluster $C_4$).

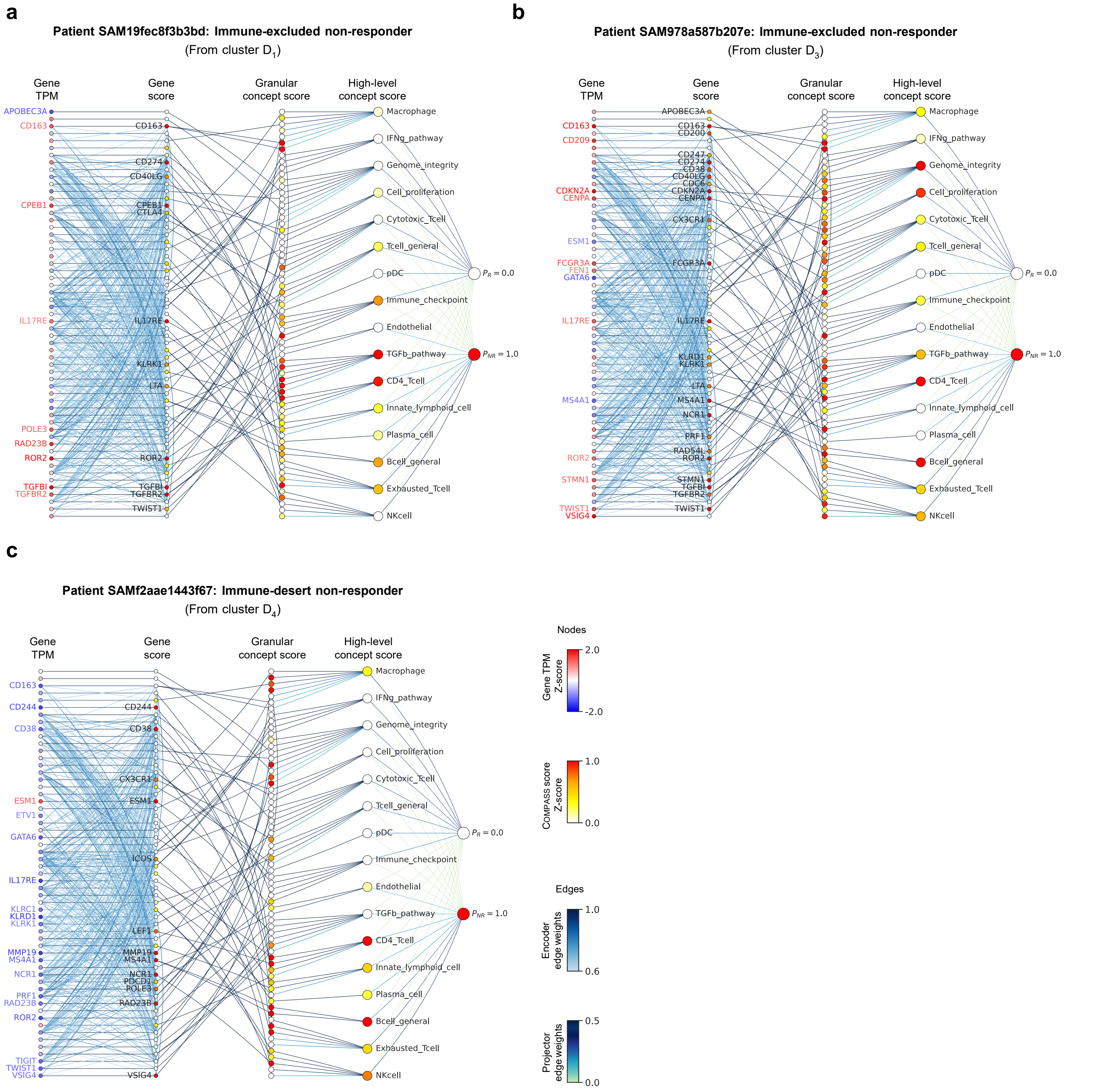


**Supplementary Fig. S27: Personalized response maps for three representative non-inflamed non-responders.** Shown are three representative patients from clusters $D_1$, $D_3$, and $D_4$ within the non-inflamed non-responder group. The representative patient from cluster $D_2$ is presented in **Fig. 6d**. **(a)** Response map for patient SAM19fec8f3b3bd (Cluster $D_1$). **(b)** Response map for patient SAM978a587b207e (Cluster $D_3$). **(c)** Response map for patient SAMf2aae1443f67 (Cluster $D_4$).

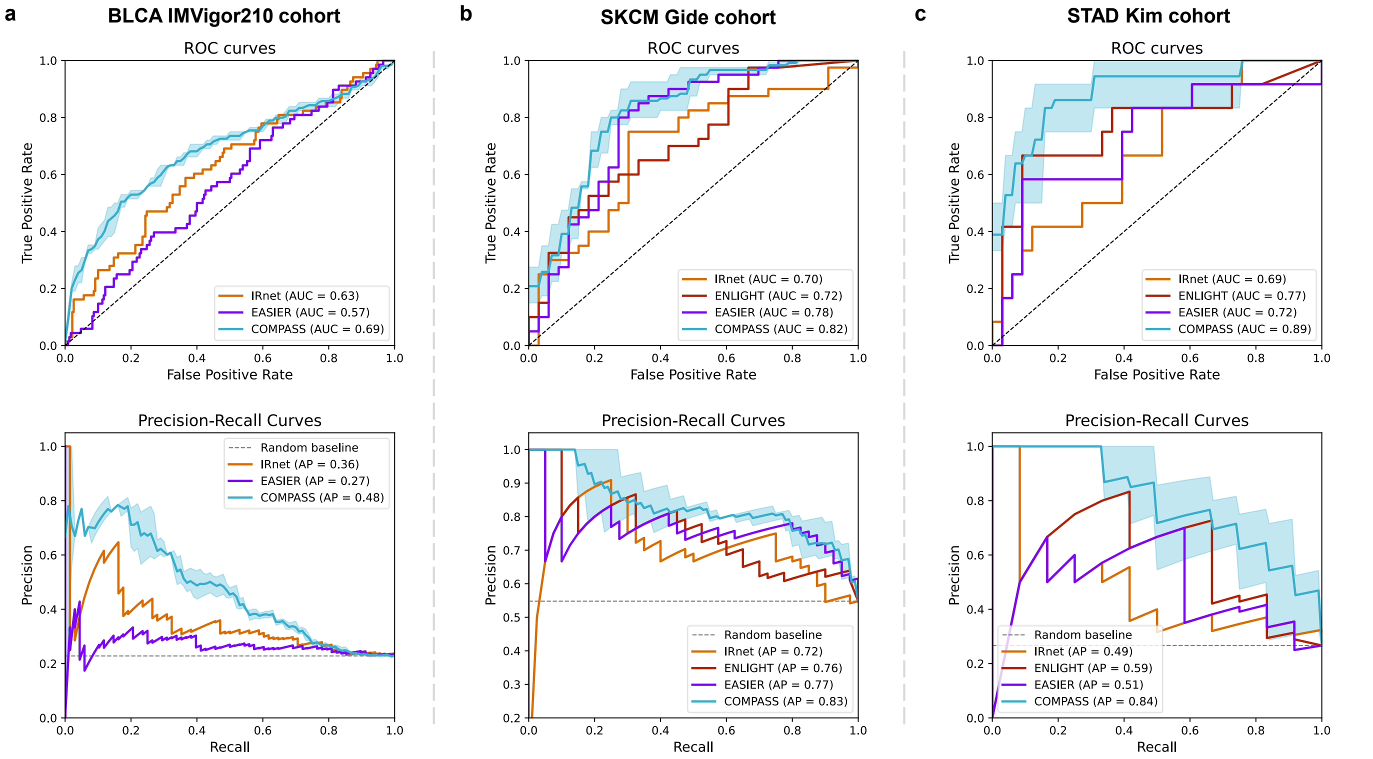


**Supplementary Fig. S28: Benchmarking against transcriptomic ML models for response prediction.** We compared the performance of COMPASS to three transcriptomic-based ML methods across three independent ICI-treated cohorts: **(a)** IMvigor210 (BLCA; anti–PD-L1), **(b)** Gide (SKCM; anti–PD-1), and **(c)** Kim (STAD; anti–PD-1). Shown are ROC curves (top) and precision–recall (PR) curves (bottom), along with the corresponding AUROC and AUPRC scores. Compared methods include IRnet (GNN-based architecture), EaSIeR (multi-view transcriptomic ensemble), and ENLIGHT (genetic interaction–based scoring). For fair comparison, retrainable models (e.g., EaSIeR) were evaluated using a leave-one-cohort-out protocol. ENLIGHT scores were obtained through the COMPASS web server and treated as continuous predictors. Lines show mean values with shaded bands representing ± 1 standard deviation across three repeats using different random seeds. IRnet predictions were generated using the authors' official implementation. Details are provided in **Supplementary Methods Method S3**. COMPASS consistently achieves superior performance across metrics and cohorts.

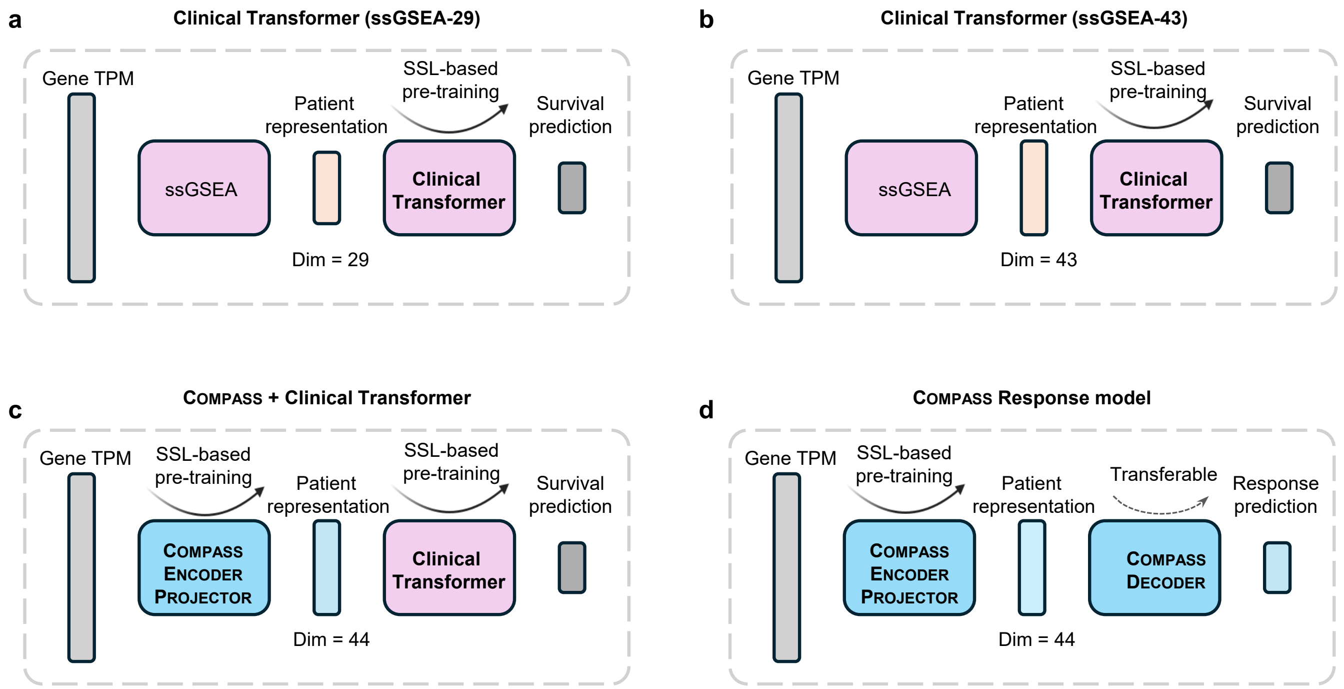


**Supplementary Fig. S29: Comparison of COMPASS and Clinical Transformer integration strategies.** **(a)** Clinical Transformer (ssGSEA-29) trained on patient representations derived from 29 immune-related signatures using ssGSEA. **(b)** Clinical Transformer (ssGSEA-43) trained on ssGSEA-derived 43 concept-level features. **(c)** COMPASS encoder-projector embedded gene expression into 44 concept-level features, which were then used as input to Clinical Transformer. **(d)** End-to-end COMPASS response model, in which SSL-pretrained encoder-projector representations are directly fed into a transferable COMPASS decoder for response prediction. Gene TPM values were first processed through different representation learning strategies, followed by SSL-based pre-training and downstream survival or response prediction. Dimensionality of patient representation is indicated below each panel.

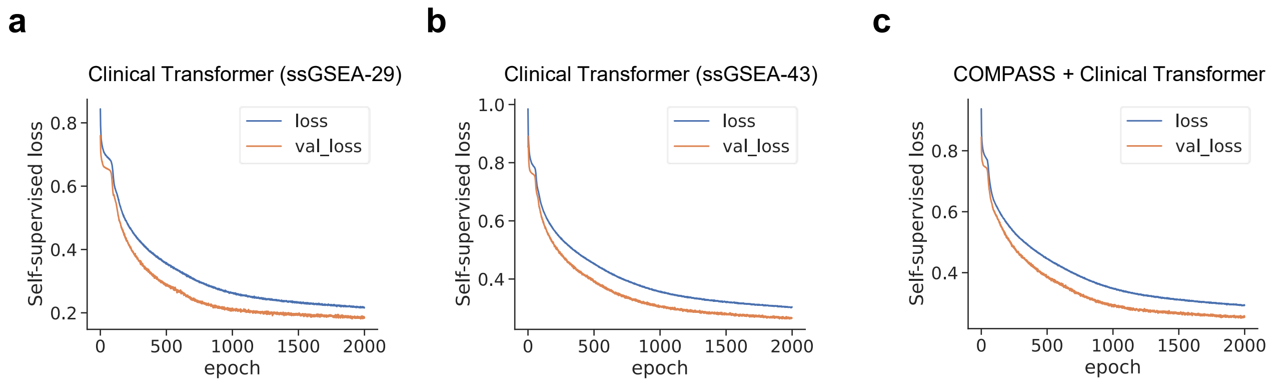


**Supplementary Fig. S30: Pretraining loss curves of Clinical Transformer models on TCGA data** Self-supervised training and validation loss trajectories over 2000 epochs are shown for models pretrained on TCGA pan-cancer ($n = 10{,}184$ patients): **(a)** Clinical Transformer (ssGSEA-29), **(b)** Clinical Transformer (ssGSEA-43), and **(c)** COMPASS + Clinical Transformer using 44 learned COMPASS concepts. Details of the pretraining are in **Supplementary Methods Method S4**. The 44 COMPASS concepts were obtained from pretraining on the same TCGA dataset. The observed differences in convergence behavior and final loss values primarily reflect the inherent dimensionality of the input features rather than methodological differences.

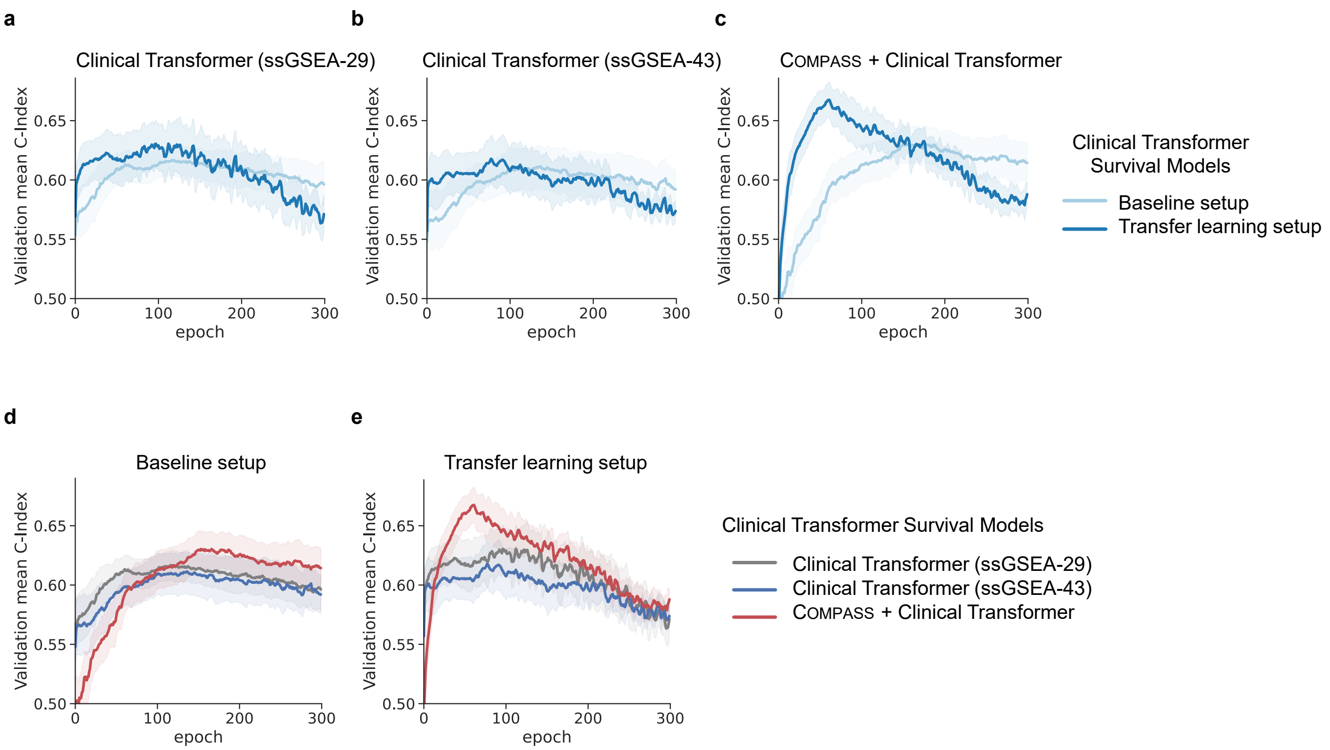


**Supplementary Fig. S31: Validation performance of Clinical Transformer survival models with the IMvigor210 cohort held out.** **(a–c)** Validation mean C-index over training epochs for **(a)** Clinical Transformer (ssGSEA-29), **(b)** Clinical Transformer (ssGSEA-43), and **(c)** COMPASS + Clinical Transformer, under baseline setup (light blue) and transfer learning setup (dark blue). Shaded regions represent standard error across 10 replicates. **(d–e)** Direct comparison of the three model variants under **(d)** baseline setup and **(e)** transfer learning setup. With transfer learning, COMPASS + Clinical Transformer consistently achieved a higher validation C-index than Clinical Transformer models using ssGSEA-derived inputs. The best validation C-index values across all training runs are summarized in **Supplementary Table S11**, and methodological details are provided in **Supplementary Methods Method S4**.

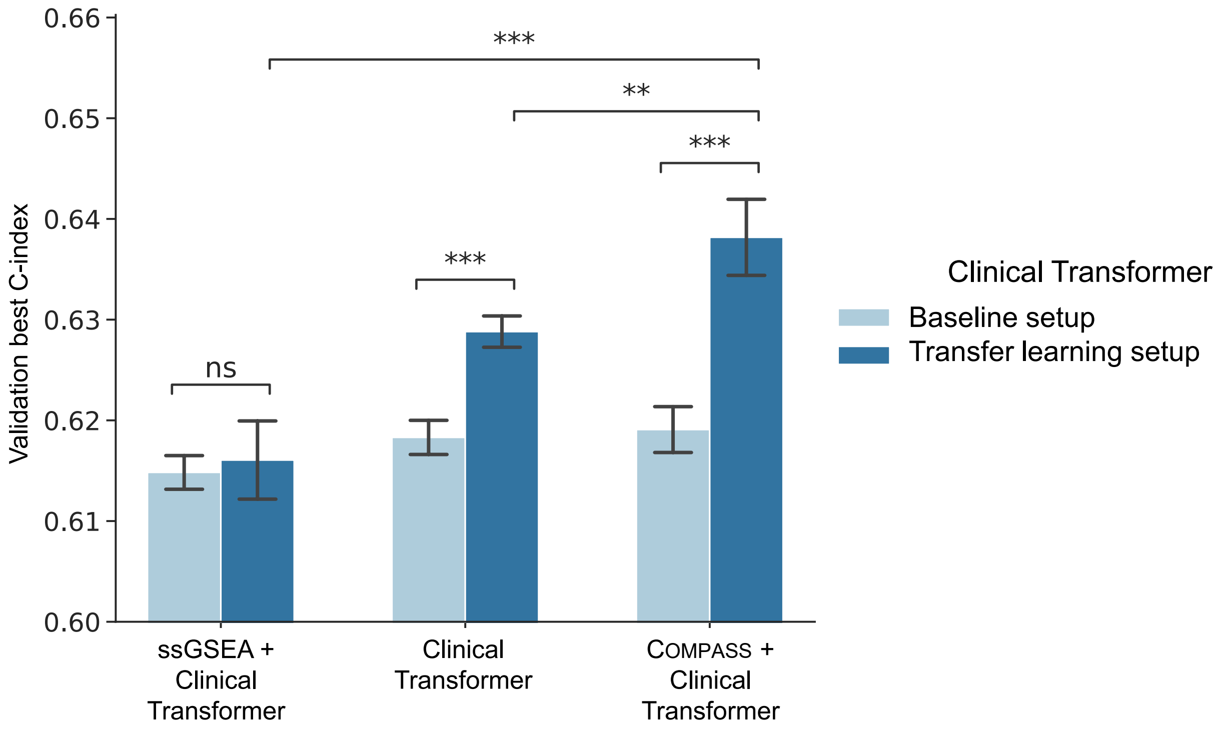


**Supplementary Fig. S32: Comparison of Clinical Transformer survival models under baseline and transfer learning setups.** Bar plots show the mean best validation C-index with error bars representing the standard error of the mean (SEM) across 11 leave-one-cohort-out experiments for Clinical Transformer (ssGSEA-29), Clinical Transformer (ssGSEA-43), and COMPASS + Clinical Transformer under the baseline (light blue) and transfer learning (dark blue) setups. Statistical comparisons were performed using two-sided paired Wilcoxon signed-rank tests with no adjustment for multiple comparisons. Exact $p$-values are as follows — baseline vs. transfer learning: Clinical Transformer $p = 9.766 \times 10^{-4}$, ssGSEA + Clinical Transformer $p = 0.638$ (ns), COMPASS + Clinical Transformer $p = 9.766 \times 10^{-4}$; among transfer learning models: Clinical Transformer vs. COMPASS + Clinical Transformer $p = 4.883 \times 10^{-3}$, ssGSEA + Clinical Transformer vs. COMPASS + Clinical Transformer $p = 9.766 \times 10^{-4}$. Validation C-index values across the 11 leave-one-cohort-out experiments are provided in **Supplementary Table S11**. Transfer learning markedly improved performance for both the Clinical Transformer (ssGSEA-43) and COMPASS + Clinical Transformer models ($p < 0.001$), whereas Clinical Transformer (ssGSEA-29) showed no significant change (ns). Among all models, COMPASS + Clinical Transformer with transfer learning achieved the highest validation C-index, demonstrating the benefit of integrating COMPASS-derived representations with Clinical Transformer architectures within a pretraining-based self-supervised learning framework. Details of the methods and experimental setup are provided in **Supplementary Figure S29** and **Supplementary Methods Method S4**.

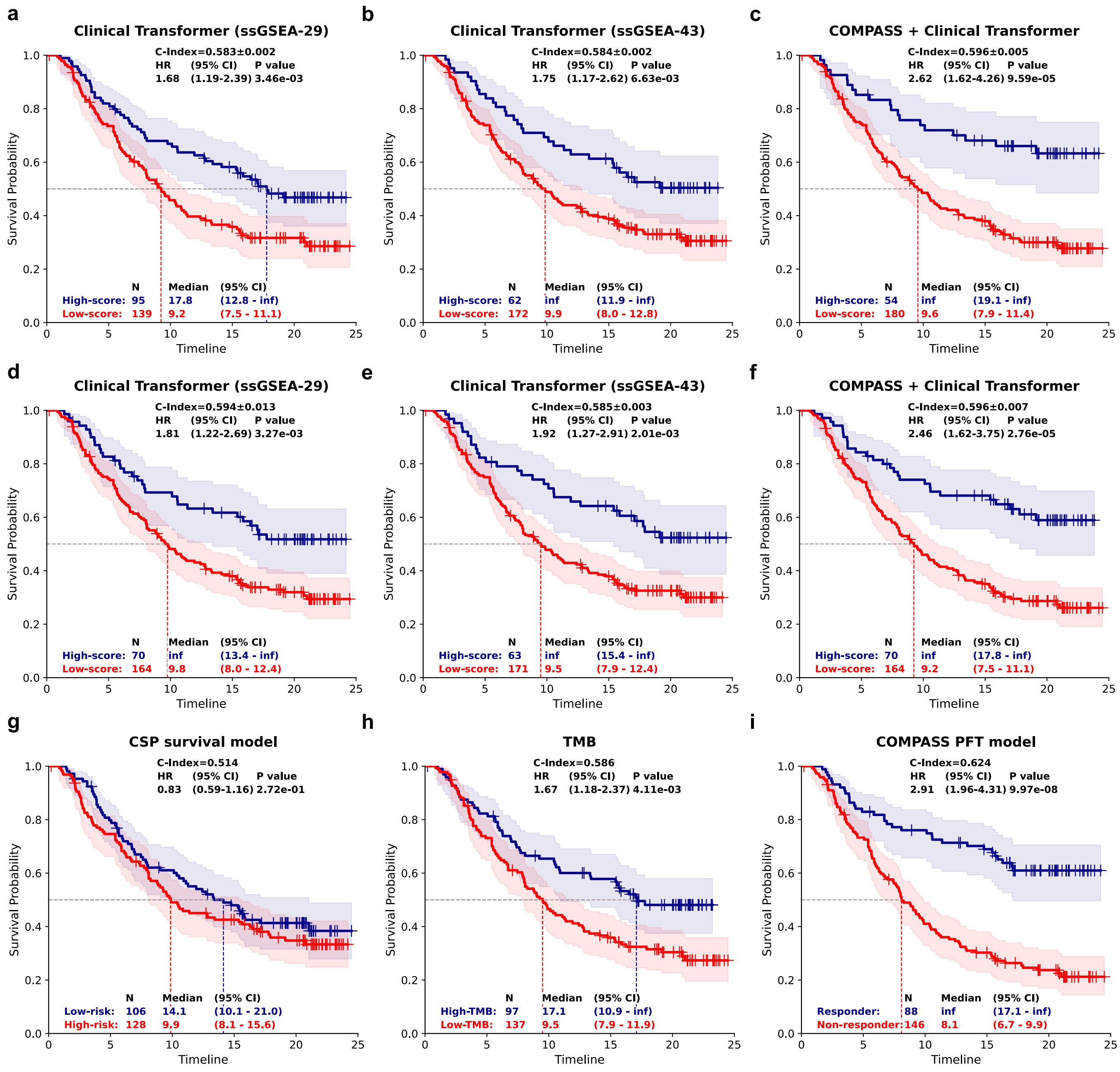


**Supplementary Fig. S33: Kaplan–Meier survival analysis of IMvigor210 patients stratified by different survival models. The IMvigor210 cohort served as the held-out test set for this evaluation. (a–c)** Kaplan–Meier curves for high-score versus low-score groups predicted under the baseline setup (training from scratch) for (a) Clinical Transformer (ssGSEA-29), (b) Clinical Transformer (ssGSEA-43), and (c) COMPASS + Clinical Transformer. **(d–f)** Corresponding Kaplan–Meier curves under the transfer learning setup (with pretraining) for (d) Clinical Transformer (ssGSEA-29), (e) Clinical Transformer (ssGSEA-43), and (f) COMPASS + Clinical Transformer. **(g–i)** Survival stratification based on (g) CSP survival model, (h) TMB, and (i) COMPASS-PFT end-to-end response prediction model. Reported metrics include C-index, hazard ratio (HR) with 95% confidence interval (CI), log-rank $p$-value, median survival time, and group sizes. Across both setups, COMPASS-based models achieved stronger survival discrimination compared to baseline models and conventional biomarkers, highlighting the benefit of pretraining and representation learning. Shaded areas represent 95% confidence intervals.

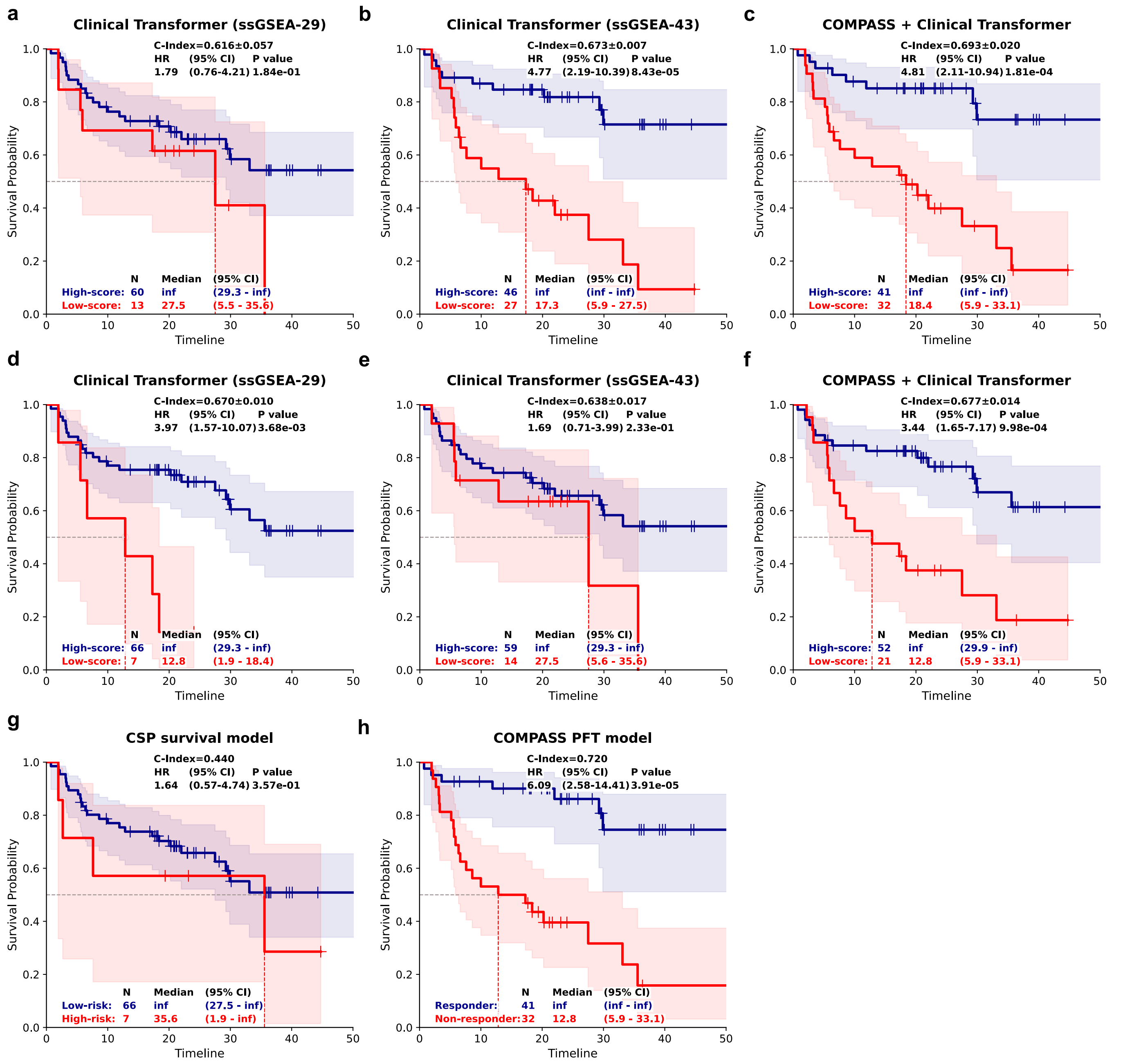


**Supplementary Fig. S34: Kaplan–Meier survival analysis of Gide cohort patients stratified by different survival models. The Gide cohort served as the held-out test set for this evaluation. (a–c)** Kaplan–Meier curves for high-score versus low-score groups predicted under the baseline setup (training from scratch) for (a) Clinical Transformer (ssGSEA-29), (b) Clinical Transformer (ssGSEA-43), and (c) COMPASS + Clinical Transformer. **(d–f)** Corresponding Kaplan–Meier curves under the transfer learning setup (with pretraining) for (d) Clinical Transformer (ssGSEA-29), (e) Clinical Transformer (ssGSEA-43), and (f) COMPASS + Clinical Transformer. **(g–h)** Survival stratification using (g) CSP survival model and (h) COMPASS-PFT end-to-end response prediction model. Reported metrics include C-index, hazard ratio (HR) with 95% confidence interval (CI), log-rank $p$-value, median survival time, and group sizes. In the Gide cohort, COMPASS-based models—particularly under transfer learning—achieved superior discrimination compared to baseline Clinical Transformer and ssGSEA approaches, and also outperformed conventional CSP survival model. Shaded areas represent 95% confidence intervals.

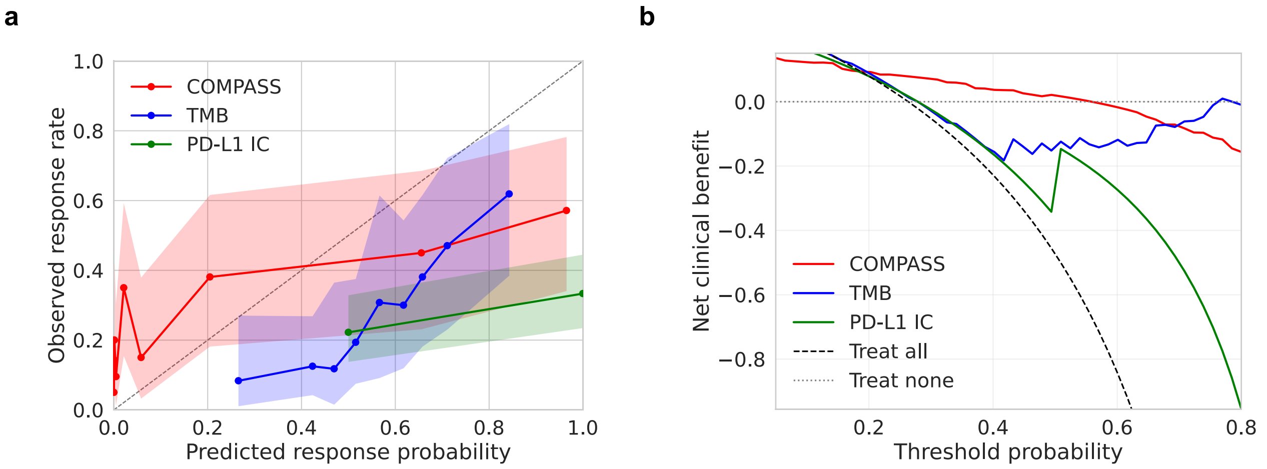


**Supplementary Fig. S35: Calibration and decision-curve analysis evaluating the clinical utility of COMPASS (a)** Calibration plots comparing predicted response probabilities with observed response rates for COMPASS, TMB, and PD-L1 IC in the IMvigor210 cohort. These plots assess how closely each model's predicted probabilities align with the empirical response frequencies across risk strata. The dashed diagonal line denotes perfect calibration, such that predicted probabilities exactly match observed response rates. Shaded areas indicate 95% confidence intervals estimated by bootstrap resampling. **(b)** Decision-curve analysis showing clinical net benefit across a range of treatment threshold probabilities. This analysis evaluates the potential clinical value of each predictor by quantifying the tradeoff between identifying responders and avoiding unnecessary treatment. COMPASS maintains positive net benefit across a broad range of thresholds and performs comparably to TMB, while exceeding PD-L1 IC as well as the treat-all and treat-none reference strategies.

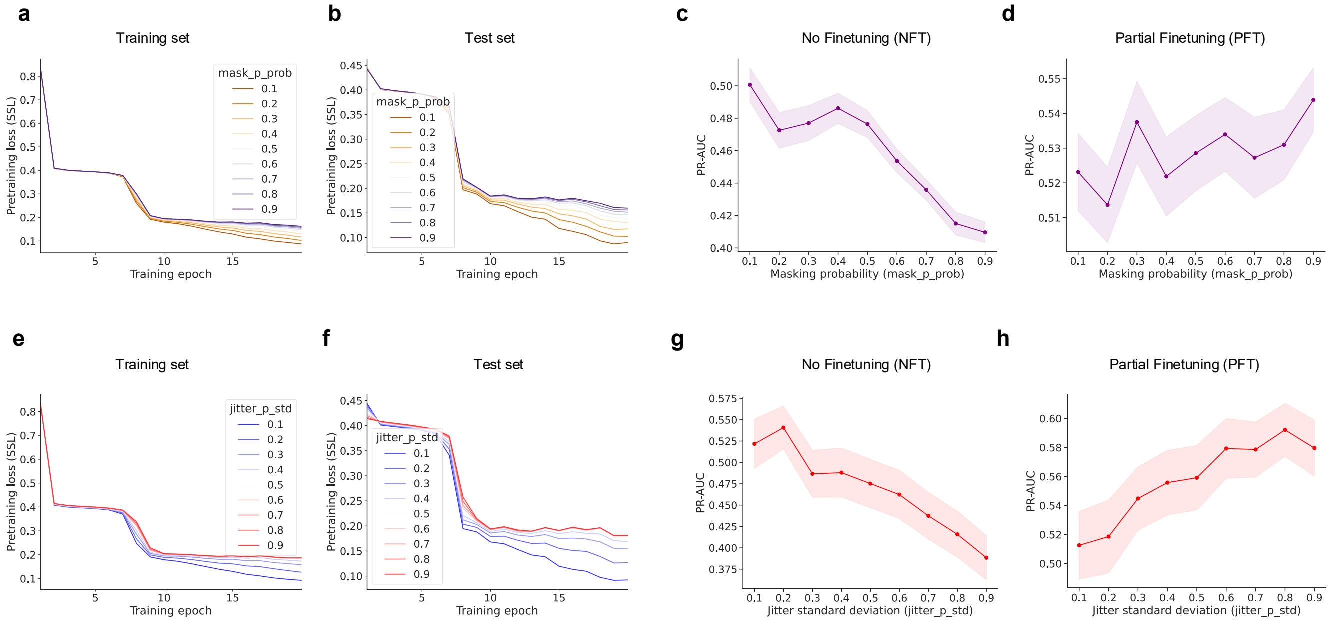


**Supplementary Fig. S36: Sensitivity analysis of `mask_p_prob` and `jitter_p_std` parameters during pretraining.** We performed a parameter sensitivity analysis to assess how increasing pretraining perturbation strength affects representation learning through mask probability (`mask_p_prob`) and feature jitter (`jitter_p_std`). Details are provided in **Supplementary Methods Method S5**. **(a–d)** Ablation of `mask_p_prob` while keeping all other parameters fixed and setting `jitter_p_std` = 0. The mask probability was varied from 0.1 to 0.9. **(a,b)** Pretraining SSL loss on the training and test sets, respectively. **(c,d)** Downstream PR-AUC under NFT (no fine-tuning) and PFT (post fine-tuning), evaluated using LOCO across 10 medium- and large-sized cohorts. These analyses quantify how stronger masking during pretraining influences both optimization behavior and transfer performance. **(e–h)** Ablation of `jitter_p_std` while keeping all other parameters fixed and setting `mask_p_prob` = 0. The jitter standard deviation was varied from 0.1 to 0.9. **(e,f)** Pretraining SSL loss on the training and test sets. **(g,h)** Downstream PR-AUC under NFT and PFT, evaluated using LOCO across the same 10 cohorts. Lines show mean PR-AUC across cohorts and shaded bands represent standard error of the mean (SEM) across 10 cohorts. These results assess the extent to which feature-level perturbations during pretraining affect the quality and transferability of the learned representations.

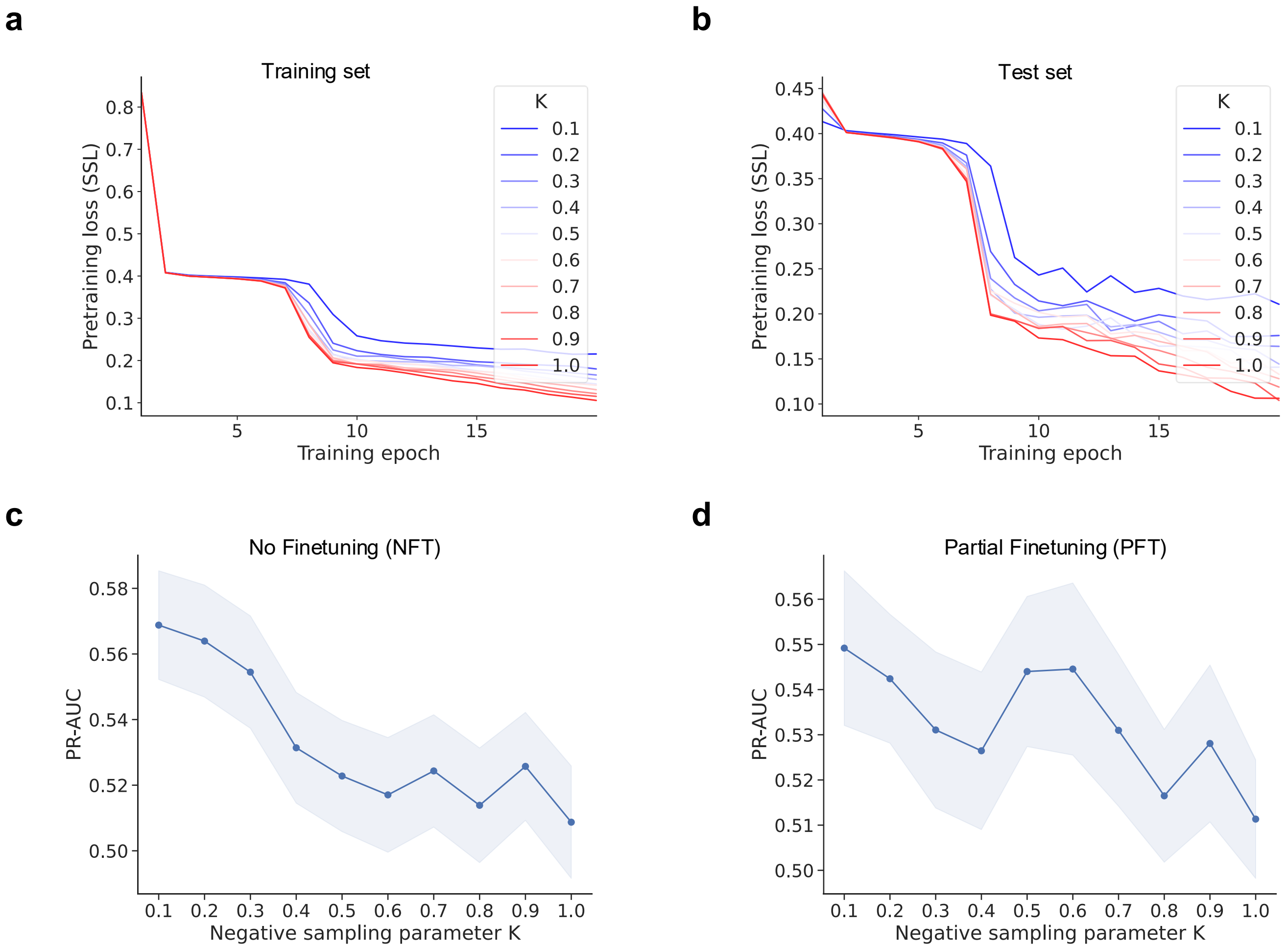


**Supplementary Fig. S37: Sensitivity analysis of the hard-negative sampling parameter $K$ during pretraining.** We evaluated how the hardness of negative sampling influences representation learning by varying the parameter $K$, which controls the size of the transcriptomic neighborhood used to draw negative samples. Details are provided in **Supplementary Methods Method S5**. **(a,b)** Pretraining self-supervised loss on the training and test sets across epochs for different values of $K$. Increasing $K$ yields lower and more stable SSL loss, consistent with easier contrastive discrimination when negatives are drawn from a broader transcriptomic neighborhood. **(c)** Downstream PR-AUC under NFT (no fine-tuning) decreases as $K$ increases, indicating that restricting negatives to more local neighborhoods improves the quality of frozen representations. **(d)** Downstream PR-AUC under PFT (post fine-tuning) shows a similar decline with increasing $K$, suggesting that local negative sampling produces more transferable representations. Lines show mean PR-AUC across cohorts and shaded bands represent standard error of the mean (SEM) across 10 cohorts. These results indicate that restricting negatives to local transcriptomic neighborhoods (smaller $K$) strengthens representation learning, whereas relaxing this locality constraint degrades both frozen and fine-tuned performance.

### Supplementary Tables

**Supplementary Table S1: Sex distribution across immunotherapy cohorts analyzed in this study.** Shown are the numbers of female and male patients in each cohort, along with the corresponding percentages. Percentages were calculated as the proportion of each sex within the total cohort size. Sex information was not available for IMmotion150, Riaz, and Snyder cohorts.

| Cohort | Female | Male | Total | Cancer type | Female (%) | Male (%) |
|---|---|---|---|---|---|---|
| IMvigor210 (n = 298) | 65 | 233 | 298 | BLCA | 21.8% | 78.2% |
| Liu (n = 107) | 47 | 60 | 107 | SKCM | 43.9% | 56.1% |
| Ravi-1 (n = 102) | 60 | 42 | 102 | LUAD | 58.8% | 41.2% |
| Rose (n = 89) | 33 | 56 | 89 | BLCA | 37.1% | 62.9% |
| Gide (n = 73) | 26 | 47 | 73 | SKCM | 35.6% | 64.4% |
| Kim (n = 45) | 12 | 33 | 45 | STAD | 26.7% | 73.3% |
| Allen (n = 39) | 14 | 25 | 39 | SKCM | 35.9% | 64.1% |
| MGH (n = 34) | 9 | 25 | 34 | SKCM | 26.5% | 73.5% |
| Hugo (n = 26) | 8 | 18 | 26 | SKCM | 30.8% | 69.2% |
| Ravi-2 (n = 25) | 10 | 15 | 25 | LUSC | 40.0% | 60.0% |
| Zhao (n = 25) | 13 | 12 | 25 | GBM | 52.0% | 48.0% |
| Miao (n = 17) | 6 | 11 | 17 | KIRC | 35.3% | 64.7% |
| Choueiri (n = 16) | 3 | 13 | 16 | KIRC | 18.8% | 81.3% |

**Supplementary Table S2: Overview of existing methods.** A total of 22 immunotherapy response prediction baseline methods are integrated into our study. Most are signature-scoring based (average, PCA, or ssGSEA) from gene expression to predict ICI response; for scoring-based methods, we trained a logistic regression (LGR) on the computed scores, and for gene-selection methods, we trained LGR on the selected genes. Two additional columns summarize each method's original design context (cancer type) and therapy/target context. "ICB (class)" denotes models developed for general immune checkpoint blockade (ICB) settings without specifying the target, while "ICB (class; biology correlate)" refers to immune or stromal biology signatures associated with ICB mechanism,s but not originally designed as predictive models. "Pan-solid tumors" refers to multi–solid-cancer analyses excluding hematologic malignancies.

| **Method** | **Description** | **Originally designed for (cancer type)** | **Therapy/target context** | **Reference** |
|---|---|---|---|---|
| GeneBio | Immunotherapy target markers (PD1, PDL1, and CTLA4) | Pan–solid tumors | PD-1 / PD-L1 / CTLA-4 (and combinations) | Kong et al., Nat Commun, 2022[1] |
| CTLA4 | Single target marker CTLA4 | General (marker-level analysis) | CTLA-4 | Kong et al., Nat Commun, 2022[1] |
| PD1 | Single target marker PDCD1 | General (marker-level analysis) | PD-1 | Kong et al., Nat Commun, 2022[1] |
| PDL1 | Single target marker CD274 | General (marker-level analysis) | PD-L1 | Kong et al., Nat Commun, 2022[1] |
| CD8 | CD8 T cell (average expression of *CD8A* and *CD8B*) | Melanoma → pan–solid tumors | ICB (class) | Chen et al., Cancer Discov., 2016[2]; Kong et al., 2022[1] |
| CIS | Davoli cytotoxic immune signature (average of cytotoxic genes) | Pan–solid tumors (TCGA) | None (biology, not therapy-specific) | Davoli et al., Science, 2017[3] |
| Teff | T-effector–IFN-$\gamma$ signature (average of Teff/IFN-$\gamma$ genes) | NSCLC | Anti-PD-L1 (atezolizumab) | Fehrenbacher et al., Lancet, 2016[4] |
| PGM | Gene pairs associated with response/survival; top model includes lymphocyte MAP4K1 and tumor TBX3 | Melanoma | ICB (class) | Freeman et al., Cell Rep Med, 2022[5] |
| NRS | Neoadjuvant response signature (average of NRS genes) | Melanoma (resectable, neoadjuvant) | Anti-PD-1 (neoadjuvant) | Huang et al., Nat Med, 2019[6] |
| IFNG | 18-gene IFN-$\gamma$–related signature (T-cell–inflamed) | Melanoma → pan–solid tumors | Anti-PD-1 (pembrolizumab) | Ayers et al., J Clin Invest, 2017[7] |
| IMPRES | Auslander IMPRES (sum of ratios of 15 checkpoint/immune gene pairs) | Melanoma | ICB (anti-CTLA-4 / anti-PD-1; combinations) | Auslander et al., Nat Med, 2018[8] |
| TIDE | Tumor Immune Dysfunction and Exclusion score | Melanoma / NSCLC (two variants) | ICB (class) | Jiang et al., Nat Med, 2018[9] |
| CTL | Cytotoxic T lymphocytes markers (average of CTL genes) | Melanoma / NSCLC (within TIDE context) | ICB (class) | Jiang et al., Nat Med, 2018[9] |
| TAM | Tumor-associated macrophages (average of TAM genes) | General (macrophage biology) | ICB (class; biology correlate) | Joyce et al., Science, 2015[10]; Jiang et al., 2018[9] |
| Texh | T-cell exhaustion signature (average of exhaustion genes) | General (exhaustion biology); validated in melanoma/NSCLC ICB | ICB (class; biology correlate) | Giordano et al., EMBO J, 2015[11]; Jiang et al., 2018[9] |
| CKS | Messina 12-chemokine signature (PCA; PC1 of 12 chemokines) | Melanoma | Immunotherapy (TLS-like biology) | Messina et al., Sci Rep, 2012[12] |
| CAF | Cancer-associated fibroblasts (average of CAF markers) | Pan–solid tumors (marker compendium) | None (not therapy-specific) | Nurmik et al., Int J Cancer, 2020[13] |
| IS | Roh immune score (average of immune genes) | Melanoma | Sequential anti-CTLA-4 → anti-PD-1 | Roh et al., Sci Transl Med, 2017[14] |
| ICA | Rooney immune cytolytic activity (GZMA and PRF1) | Pan–solid tumors (TCGA) | None (biology, not therapy-specific) | Rooney et al., Cell, 2015[15] |
| MIAS | MHC-I association immunoscore (100 genes; ssGSEA) | Melanoma (6 cohorts) | Anti-PD-1 | Wu et al., Nat Commun, 2022[16] |
| GEP | T cell–inflamed GEP score (ssGSEA NES of 18 genes) | Pan–solid tumors (22 types, KEYNOTE trials) | Anti-PD-1 (pembrolizumab) | Cristescu et al., Science, 2018[17]; Wu et al., 2022[16] |
| NetBio200 | Top 200 genes with highest influence scores as ICI target-proximal genes | Pan–solid tumors (network-based design) | Anti-CTLA-4 / anti-PD-1 / anti-PD-L1 / combinations | Kong et al., Nat Commun, 2022[1] |

**Supplementary Table S3: Performance comparison of methods in leave-one-cohort-out validation.** This table reports Accuracy (ACC, expressed as mean ± standard deviation in %), Matthews correlation coefficient (MCC), Precision-Recall area under the curve (PR-AUC), and ROC area under the curve (ROC-AUC) for each method. For the 4 fine-tuning modes of COMPASS, the experiments were repeated 3 times using different random seeds (24, 42, and 64), and the ± represent the standard deviation across these repetitions. Results are grouped by cohort size: 4 large cohorts ($n = 672$), 6 medium cohorts ($n = 331$), and 6 small cohorts ($n = 130$). For each metric within each cohort group, the best-performing result is highlighted in bold.

| Group | **Accuracy (%) ↑** | | | **MCC ↑** | | | **PR-AUC ↑** | | | **ROC-AUC ↑** | | |
|---|---|---|---|---|---|---|---|---|---|---|---|---|
| | Large (n = 672) | Medium (n = 331) | Small (n = 130) | Large (n = 672) | Medium (n = 331) | Small (n = 130) | Large (n = 672) | Medium (n = 331) | Small (n = 130) | Large (n = 672) | Medium (n = 331) | Small (n = 130) |
| COMPASS-PFT | **72.4±1.6** | 74.6±0.5 | **62.3±1.5** | **0.325±0.026** | **0.438±0.017** | 0.097±0.049 | 0.573±0.017 | 0.600±0.030 | 0.411±0.009 | **0.718±0.004** | 0.784±0.012 | 0.532±0.014 |
| COMPASS-LFT | 69.8±4.9 | **75.2±1.3** | 61.3±0.4 | 0.304±0.026 | 0.428±0.043 | 0.079±0.022 | **0.580±0.017** | **0.621±0.048** | **0.426±0.005** | 0.718±0.006 | **0.785±0.023** | 0.545±0.020 |
| COMPASS-FFT | 59.7±1.9 | 65.7±4.9 | 60.0±1.1 | 0.101±0.023 | 0.243±0.049 | 0.056±0.022 | 0.377±0.011 | 0.509±0.042 | 0.370±0.004 | 0.582±0.011 | 0.688±0.017 | 0.520±0.032 |
| NetBio | 63.8 | 61.9 | 53.1 | 0.170 | 0.232 | 0.022 | 0.369 | 0.421 | 0.395 | 0.625 | 0.678 | 0.505 |
| Teff | 57.9 | 65.9 | 54.6 | 0.136 | 0.288 | 0.063 | 0.382 | 0.473 | 0.402 | 0.604 | 0.682 | 0.500 |
| CD8 | 58.3 | 64.0 | 54.6 | 0.136 | 0.253 | 0.055 | 0.363 | 0.458 | 0.374 | 0.589 | 0.651 | 0.487 |
| CTL | 58.5 | 63.7 | 53.8 | 0.152 | 0.239 | 0.043 | 0.364 | 0.467 | 0.394 | 0.589 | 0.658 | 0.504 |
| CIS | 57.9 | 63.7 | 53.1 | 0.136 | 0.248 | 0.031 | 0.351 | 0.458 | 0.394 | 0.574 | 0.647 | 0.517 |
| IS | 55.1 | 63.4 | 55.4 | 0.110 | 0.267 | **0.099** | 0.356 | 0.477 | 0.379 | 0.570 | 0.659 | 0.525 |
| ICA | 58.0 | 62.2 | 53.1 | 0.148 | 0.213 | 0.055 | 0.363 | 0.474 | 0.390 | 0.583 | 0.648 | 0.507 |
| PDL1 | 52.7 | 63.1 | 54.6 | 0.083 | 0.217 | 0.013 | 0.314 | 0.451 | 0.366 | 0.543 | 0.632 | 0.514 |
| PGM | 58.2 | 63.4 | 48.5 | 0.129 | 0.287 | 0.010 | 0.377 | 0.530 | 0.380 | 0.583 | 0.706 | 0.524 |
| CKS | 56.2 | 64.7 | 47.7 | 0.134 | 0.266 | −0.052 | 0.377 | 0.432 | 0.359 | 0.588 | 0.669 | 0.463 |
| PD1 | 64.7 | 54.7 | 48.5 | 0.158 | 0.041 | 0.010 | 0.380 | 0.354 | 0.384 | 0.562 | 0.546 | 0.513 |
| TIDE | 53.1 | 61.0 | 50.8 | 0.033 | 0.173 | −0.022 | 0.312 | 0.417 | 0.459 | 0.522 | 0.594 | 0.505 |
| GeneBio | 53.0 | 62.2 | 47.7 | 0.095 | 0.222 | −0.095 | 0.373 | 0.460 | 0.335 | 0.574 | 0.647 | 0.459 |
| TAM | 55.8 | 53.8 | 53.1 | 0.071 | 0.034 | 0.006 | 0.335 | 0.349 | 0.372 | 0.542 | 0.518 | 0.546 |
| COMPASS-NFT | 57.0 | 54.7 | 49.2 | 0.067 | 0.182 | 0.066 | 0.335 | 0.338 | 0.356 | 0.579 | 0.590 | 0.497 |
| CTLA4 | 49.4 | 56.2 | 53.1 | 0.069 | 0.023 | −0.021 | 0.278 | 0.308 | 0.335 | 0.500 | 0.503 | 0.464 |
| IMPRES | 47.9 | 58.3 | 51.5 | 0.101 | 0.130 | −0.117 | 0.303 | 0.431 | 0.344 | 0.539 | 0.631 | 0.478 |
| GEP | 55.8 | 58.0 | 43.8 | 0.158 | 0.252 | −0.021 | 0.411 | 0.523 | 0.344 | 0.613 | 0.705 | 0.482 |
| CAF | 55.2 | 49.5 | 51.5 | 0.090 | 0.016 | 0.024 | 0.324 | 0.359 | 0.388 | 0.539 | 0.541 | **0.558** |
| Texh | 54.0 | 48.6 | 53.1 | 0.058 | −0.065 | 0.006 | 0.321 | 0.304 | 0.360 | 0.542 | 0.436 | 0.498 |
| IFNG | 41.8 | 49.8 | 48.5 | −0.016 | 0.030 | 0.054 | 0.296 | 0.351 | 0.395 | 0.499 | 0.546 | 0.522 |
| MIAS | 37.6 | 57.7 | 44.6 | −0.013 | 0.237 | 0.001 | 0.341 | 0.459 | 0.355 | 0.537 | 0.676 | 0.465 |
| NRS | 48.8 | 42.6 | 45.4 | −0.019 | −0.151 | −0.139 | 0.314 | 0.257 | 0.316 | 0.493 | 0.387 | 0.384 |

**Supplementary Table S4: Accuracy and PR-AUC of different methods for predicting new indication cohorts.** Accuracy (%) and PR-AUC are reported for five test cohorts under a new indication setting. For COMPASS-PFT, the results are shown as mean ± standard deviation across three independent runs. The best-performing method for each cohort and metric is highlighted in bold.

| New Indication | Training / Test Size | Accuracy (%) ↑ | | | | | PR-AUC ↑ | | | | |
|---|---|---|---|---|---|---|---|---|---|---|---|
| | | COMPASS-PFT | Teff | NetBio | PGM | Reference | COMPASS-PFT | Teff | NetBio | PGM | Reference |
| KIRC (Urothelial Carcinoma) | 725 / 408 | **71.9 ± 0.8** | 65.2 | 69.7 | 37.4 | 59.4 | **0.44 ± 0.03** | 0.37 | 0.35 | 0.36 | 0.22 |
| SKCM (Skin Melanoma) | 803 / 330 | **65.7 ± 1.4** | 60.0 | 50.6 | 59.7 | 52.2 | **0.62 ± 0.01** | 0.49 | 0.42 | 0.57 | 0.39 |
| BLCA (Renal Carcinoma) | 935 / 198 | **72.9 ± 2.4** | 60.3 | 71.8 | 72.3 | 65.3 | **0.46 ± 0.02** | 0.29 | 0.27 | 0.23 | 0.28 |
| LUAD (Lung Adenocarcinoma) | 1031 / 102 | **76.5 ± 1.7** | 58.8 | 58.8 | 51.0 | 53.2 | **0.70 ± 0.03** | 0.55 | 0.46 | 0.49 | 0.37 |
| STAD (Stomach Adenocarcinoma) | 1088 / 45 | **83.7 ± 3.4** | 75.6 | 46.7 | 64.4 | 60.9 | **0.85 ± 0.04** | 0.76 | 0.24 | 0.32 | 0.27 |

**Supplementary Table S5: Accuracy and PR-AUC of different methods for predicting new ICI-treatment cohorts.** Accuracy (%) and PR-AUC values are reported for five test cohorts representing new ICI drugs. For COMPASS-PFT, values are shown as mean ± standard deviation across three independent runs. The best-performing method for each cohort and metric is highlighted in bold.

| New Drug | Training / Test Size | Accuracy (%) ↑ | | | | | PR-AUC ↑ | | | | |
|---|---|---|---|---|---|---|---|---|---|---|---|
| | | COMPASS-PFT | Teff | NetBio | PGM | Reference | COMPASS-PFT | Teff | NetBio | PGM | Reference |
| Atezolizumab | 606 / 527 | **69.3 ± 1.4** | 59.2 | 64.9 | 64.7 | 61.9 | **0.46 ± 0.03** | 0.37 | 0.31 | 0.30 | 0.26 |
| Pembrolizumab | 874 / 259 | **71.0 ± 0.4** | 53.7 | 59.5 | 55.2 | 54.2 | **0.64 ± 0.03** | 0.44 | 0.48 | 0.47 | 0.36 |
| Nivolumab | 913 / 220 | **63.6 ± 2.1** | 51.4 | 45.9 | 50.9 | 59.5 | **0.47 ± 0.01** | 0.35 | 0.35 | 0.45 | 0.28 |
| Ipilimumab | 1094 / 39 | **76.1 ± 5.3** | 59.0 | 53.8 | 51.3 | 55.6 | **0.76 ± 0.07** | 0.44 | 0.43 | 0.61 | 0.33 |
| Ipi+Pembro | 1108 / 25 | **85.3 ± 2.3** | 72.0 | 76.0 | 68.0 | 53.9 | **0.88 ± 0.02** | **0.93** | 0.90 | 0.76 | 0.64 |

**Supplementary Table S6: Accuracy and PR-AUC of different methods for predicting new ICI-target cohorts.** Accuracy (%) and PR-AUC values are reported for four ICI target settings. For COMPASS-PFT, results are reported as mean ± standard deviation over three independent runs. The best result for each metric and target is shown in bold.

| New Target | Training / Test Size | Accuracy (%) ↑ | | | | | PR-AUC ↑ | | | | |
|---|---|---|---|---|---|---|---|---|---|---|---|
| | | COMPASS-PFT | Teff | NetBio | PGM | Reference | COMPASS-PFT | Teff | NetBio | PGM | Reference |
| PDL1 | 602 / 531 | **68.9 ± 3.1** | 59.1 | 64.6 | 64.8 | 61.9 | **0.48 ± 0.04** | 0.37 | 0.31 | 0.30 | 0.26 |
| PD1 | 642 / 491 | **67.5 ± 0.6** | 54.0 | 47.3 | 55.4 | 56.5 | **0.55 ± 0.06** | 0.39 | 0.36 | 0.45 | 0.32 |
| CTLA4 | 1076 / 57 | **70.8 ± 2.0** | 61.4 | 50.9 | 50.9 | 55.6 | **0.72 ± 0.09** | 0.43 | 0.45 | 0.59 | 0.33 |
| PD1+CTLA4 | 1087 / 46 | **72.5 ± 5.0** | 65.2 | 58.7 | 65.2 | 56.0 | **0.83 ± 0.03** | **0.90** | 0.83 | 0.76 | 0.67 |

**Supplementary Table S7: Summary of datasets for drug-specific model development**

| Drug Model | Dataset | Description | Total | R | NR |
|---|---|---|---|---|---|
| Atezolizumab model | Stage-1 set (General ICI) | Data excluding all PD-L1 treatments | 602 | 210 | 392 |
| | Stage-2 set (Drug-specific) | Bladder cancer (BLCA) cohort receiving Atezo treatment | 354 | 84 | 270 |
| | Test set (Drug-specific) | Kidney cancer (KIRC) cohort receiving Atezo treatment | 167 | 49 | 118 |
| Pembrolizumab model | Stage-1 set (General ICI) | Data excluding all PD-1 treatments | 596 | 158 | 438 |
| | Stage-2 set (Drug-specific) | Melanoma (SKCM) cohort receiving Pembro treatment | 120 | 53 | 67 |
| | Test set (Drug-specific) | Adenocarcinoma cohorts (STAD + LUAD) receiving Pembro treatment | 78 | 22 | 56 |
| Nivolumab model | Stage-1 set (General ICI) | Data excluding all PD-1 treatments | 596 | 158 | 438 |
| | Stage-2 set (Drug-specific) | Melanoma (SKCM) cohort receiving Nivo treatment | 105 | 31 | 74 |
| | Test set (Drug-specific) | Lung cancer cohorts (LUAD + LUSC) receiving Nivo treatment | 63 | 21 | 42 |

**Supplementary Table S8: Accuracy and PR-AUC of different drug-specific models.** Metrics are reported for three drug-specific prediction tasks. For models with repeated experiments, results are shown as mean $\pm$ standard deviation over three independent runs. The best-performing value in each row is highlighted in bold.

| **Accuracy (%) ↑** | | | | | |
|---|---|---|---|---|---|
| **Drug Model** | **Reference** | **PGM** | **COMPASS-SSFT1** | **COMPASS-SSFT2** | **COMPASS-MSFT** |
| Atezolizumab model | 58.5 | 67.1 | 70.3 $\pm$ 0.7 | 60.7 $\pm$ 2.8 | **73.7 $\pm$ 2.4** |
| Pembrolizumab model | 59.5 | 59.0 | 77.4 $\pm$ 3.7 | 78.6 $\pm$ 3.9 | **82.9 $\pm$ 2.7** |
| Nivolumab model | 55.6 | 60.3 | 67.2 $\pm$ 0.9 | 72.5 $\pm$ 0.9 | **73.0 $\pm$ 1.6** |
| **PR-AUC ↑** | | | | | |
| **Drug Model** | **Reference** | **PGM** | **COMPASS-SSFT1** | **COMPASS-SSFT2** | **COMPASS-MSFT** |
| Atezolizumab model | 0.29 | 0.39 | 0.39 $\pm$ 0.05 | **0.54 $\pm$ 0.12** | 0.50 $\pm$ 0.09 |
| Pembrolizumab model | 0.28 | 0.41 | 0.61 $\pm$ 0.12 | 0.67 $\pm$ 0.05 | **0.73 $\pm$ 0.03** |
| Nivolumab model | 0.33 | 0.53 | 0.44 $\pm$ 0.09 | 0.60 $\pm$ 0.03 | **0.61 $\pm$ 0.05** |

**Supplementary Table S9: Benchmarking learned patient representations against fixed signature scoring (AUROC).** Leave-one-cohort-out AUROC across ten cohorts for logistic regression (LGR) models using fixed gene set scoring methods (Average and ssGSEA) and learned COMPASS patient representations at the granular (132-dimensional) and high-level (43-dimensional) concept levels. The bottom row reports the mean ± SD AUROC across all ten folds. Bold values indicate the best-performing method within each cohort. Details of the experimental setup are provided in Supplementary Methods **Method S2**.

| Model | **Granular concepts** (dim = 132) | | | **High-level concepts** (dim = 43) | | |
|---|---|---|---|---|---|---|
| | TPM-Average-LGR | TPM-ssGSEA-LGR | TPM-COMPASS (Encoder–Projector)-LGR | TPM-Average-LGR | TPM-ssGSEA-LGR | TPM-COMPASS (Encoder–Projector)-LGR |
| IMVigor210 (n=298) | 0.658 | 0.656 | **0.712** | 0.642 | 0.637 | **0.696** |
| IMmotion150 (n=165) | **0.722** | 0.705 | 0.690 | 0.705 | 0.662 | **0.743** |
| Liu (n=107) | 0.540 | 0.543 | **0.712** | 0.590 | 0.613 | **0.674** |
| Ravi-1 (n=102) | 0.665 | 0.654 | **0.752** | 0.650 | 0.660 | **0.784** |
| Rose (n=89) | 0.743 | 0.729 | **0.782** | **0.767** | 0.744 | 0.717 |
| Gide (n=73) | 0.768 | 0.798 | **0.839** | 0.761 | 0.782 | **0.831** |
| Riaz (n=51) | 0.583 | 0.593 | **0.737** | 0.544 | 0.580 | **0.707** |
| Kim (n=45) | 0.912 | 0.904 | **0.939** | 0.927 | 0.939 | **0.960** |
| Allen (n=39) | 0.642 | 0.642 | **0.787** | 0.663 | 0.675 | **0.888** |
| MGH (n=34) | 0.716 | 0.697 | **0.742** | 0.693 | 0.655 | **0.773** |
| **Mean** | 0.695±0.104 | 0.692±0.103 | 0.769±0.074 | 0.694±0.107 | 0.695±0.104 | **0.777±0.092** |

**Supplementary Table S10: Benchmarking learned patient representations against fixed signature scoring (AUPRC).** Leave-one-cohort-out AUPRC across ten cohorts for logistic regression (LGR) models using fixed gene set scoring methods (Average and ssGSEA) and learned COMPASS patient representations at the granular (132-dimensional) and high-level (43-dimensional) concept levels. The bottom row reports the mean ± SD AUROC across all ten folds. Bold values indicate the best-performing method within each cohort. Details of the experimental setup are provided in Supplementary Methods **Method S2**.

| Model | **Granular concepts** (dim = 132) | | | **High-level concepts** (dim = 43) | | |
|---|---|---|---|---|---|---|
| | TPM-Average-LGR | TPM-ssGSEA-LGR | TPM-COMPASS (Encoder-Projector)-LGR | TPM-Average-LGR | TPM-ssGSEA-LGR | TPM-COMPASS (Encoder-Projector)-LGR |
| IMVigor210 (n=298) | 0.327 | 0.314 | 0.431 | 0.316 | 0.308 | **0.459** |
| IMmotion150 (n=165) | 0.452 | 0.444 | 0.594 | 0.451 | 0.437 | **0.615** |
| Liu (n=107) | 0.459 | 0.470 | **0.580** | 0.503 | 0.523 | 0.571 |
| Ravi-1 (n=102) | 0.529 | 0.519 | 0.715 | 0.483 | 0.501 | **0.737** |
| Rose (n=89) | 0.329 | 0.313 | **0.453** | 0.375 | 0.367 | 0.356 |
| Gide (n=73) | 0.817 | 0.838 | **0.874** | 0.823 | 0.831 | 0.848 |
| Riaz (n=51) | 0.243 | 0.248 | **0.480** | 0.241 | 0.243 | 0.383 |
| Kim (n=45) | 0.793 | 0.825 | 0.792 | 0.852 | 0.899 | **0.879** |
| Allen (n=39) | 0.526 | 0.524 | 0.741 | 0.531 | 0.528 | **0.862** |
| MGH (n=34) | 0.687 | 0.682 | 0.680 | 0.663 | 0.648 | **0.711** |
| **Mean** | 0.516±0.197 | 0.518±0.208 | 0.634±0.151 | 0.524±0.203 | 0.528±0.213 | **0.642±0.197** |

**Supplementary Table S11: Validation performance of Clinical Transformer survival models across feature representations and learning strategies.** For each cohort, values represent validation (not test) performance when that cohort was held out for hyperparameter tuning. Results are reported as mean concordance index (C-index) ± standard deviation across ten random 10% train/validation splits in the leave-one-cohort-out setting. Bold values indicate the best-performing model within each row. Baseline models were trained from scratch, while transfer learning models were initialized from TCGA-pretrained checkpoints obtained through self-supervised pretraining. Details of the methods and experimental setup are provided in **Supplementary Methods Method S4**.

| **Survival Model** | **Baseline Setup** | | | **Transfer Learning Setup** | | |
|---|---|---|---|---|---|---|
| | Clinical Transformer (ssGSEA-29) | Clinical Transformer (ssGSEA-43) | COMPASS + Clinical Transformer | Clinical Transformer (ssGSEA-29) | Clinical Transformer (ssGSEA-43) | COMPASS + Clinical Transformer |
| Leave-IMVigor210 (n=298) | 0.617±0.039 | 0.611±0.057 | 0.630±0.048 | 0.631±0.064 | 0.618±0.062 | **0.668±0.048** |
| Leave-Liu (n=107) | 0.626±0.051 | 0.620±0.035 | 0.624±0.039 | 0.638±0.042 | 0.639±0.034 | **0.648±0.036** |
| Leave-Ravi-1 (n=98) | 0.631±0.034 | 0.620±0.046 | 0.632±0.038 | 0.636±0.051 | 0.636±0.053 | **0.648±0.044** |
| Leave-Ravi-2 (n=25) | 0.615±0.034 | 0.614±0.046 | 0.615±0.049 | 0.624±0.041 | 0.610±0.044 | **0.632±0.041** |
| Leave-Rose (n=89) | 0.615±0.053 | 0.607±0.052 | 0.620±0.039 | 0.630±0.041 | 0.604±0.054 | **0.633±0.027** |
| Leave-Snyder (n=21) | 0.618±0.042 | 0.611±0.040 | 0.622±0.029 | 0.625±0.041 | 0.613±0.037 | **0.643±0.031** |
| Leave-Hugo (n=25) | 0.621±0.058 | 0.615±0.050 | 0.615±0.030 | 0.621±0.039 | 0.621±0.051 | **0.625±0.045** |
| Leave-Gide (n=73) | 0.617±0.040 | 0.611±0.033 | 0.615±0.038 | 0.626±0.049 | 0.594±0.025 | **0.635±0.036** |
| Leave-Riaz (n=51) | 0.614±0.040 | 0.626±0.034 | 0.616±0.040 | 0.631±0.031 | 0.614±0.029 | **0.633±0.021** |
| Leave-Allen (n=39) | 0.613±0.042 | 0.612±0.020 | 0.605±0.028 | **0.630±0.036** | 0.611±0.038 | 0.626±0.022 |
| Leave-MGH (n=34) | 0.615±0.042 | 0.615±0.029 | 0.617±0.036 | 0.625±0.035 | 0.617±0.021 | **0.629±0.018** |
| **Average** | 0.618±0.006 | 0.615±0.006 | 0.619±0.008 | 0.629±0.005 | 0.616±0.013 | **0.638±0.013** |

**Supplementary Table S12: Test cohort evaluation of transformer-based survival models under the leave-one-cohort-out setting**. Reported values are mean ± SD C-index scores across cohorts for baseline and transfer learning variants of the Clinical Transformer, TMB, the CSP transformer model, and the COMPASS-PFT response model. Bold values indicate the best-performing method within each row. *TMB: results were computed only for patients with available TMB data (IMVigor210, n=234; Liu, n=107; Rose, n=88; Ravi-1, n=51; Riaz, n=46; Allen, n=39; MGH, n=28; Hugo, n=25; Ravi-2, n=10). The COMPASS-PFT model was trained on the same data as the Clinical Transformer and used predicted non-response probabilities as risk scores for C-index calculation. Details of the methods and experimental setup are provided in **Supplementary Methods Method S4**

| **SurvivalModel** | **Baseline Setup** | | | **Transfer Learning Setup** | | | **TMB*** | cGAS-STING pathway (CSP) Transformer | COMPASS PFT model |
|---|---|---|---|---|---|---|---|---|---|
| | Clinical Transformer (ssGSEA-29) | Clinical Transformer (ssGSEA-43) | COMPASS + Clinical Transformer | Clinical Transformer (ssGSEA-29) | Clinical Transformer (ssGSEA-43) | COMPASS + Clinical Transformer | | | |
| IMVigor210 (n=298) | 0.569±0.001 | 0.568±0.002 | 0.583±0.004 | 0.579±0.008 | 0.574±0.007 | 0.589±0.008 | 0.586 | 0.512 | **0.610±0.017** |
| Liu (n=107) | 0.591±0.023 | 0.520±0.021 | 0.568±0.013 | 0.577±0.033 | 0.549±0.032 | 0.584±0.016 | 0.568 | 0.628 | **0.652±0.010** |
| Ravi-1 (n=98) | 0.588±0.016 | 0.610±0.011 | 0.574±0.038 | **0.675±0.039** | 0.614±0.017 | 0.631±0.004 | 0.588 | 0.613 | 0.619±0.008 |
| Rose (n=89) | **0.626±0.011** | 0.613±0.007 | 0.562±0.013 | 0.495±0.010 | 0.562±0.006 | 0.603±0.011 | 0.577 | 0.516 | 0.516±0.032 |
| Gide (n=73) | 0.616±0.057 | 0.673±0.007 | 0.693±0.020 | 0.670±0.010 | 0.638±0.017 | 0.677±0.014 | n.a. | 0.440 | **0.720±0.011** |
| Riaz (n=51) | 0.562±0.038 | 0.535±0.020 | 0.619±0.015 | 0.570±0.007 | 0.591±0.009 | 0.615±0.002 | 0.446 | **0.657** | 0.616±0.014 |
| Van Allen (n=39) | 0.671±0.006 | 0.612±0.026 | **0.702±0.008** | 0.520±0.034 | 0.640±0.035 | 0.686±0.020 | 0.590 | 0.452 | 0.624±0.035 |
| MGH (n=34) | 0.636±0.007 | 0.528±0.022 | 0.611±0.020 | 0.633±0.028 | 0.540±0.047 | 0.553±0.038 | **0.710** | 0.498 | 0.646±0.043 |
| Ravi-2 (n=25) | 0.646±0.034 | 0.568±0.053 | 0.669±0.052 | 0.617±0.035 | 0.603±0.020 | 0.642±0.051 | 0.375 | **0.704** | 0.568±0.022 |
| Hugo (n=25) | 0.432±0.030 | 0.545±0.048 | 0.523±0.053 | 0.543±0.013 | 0.457±0.019 | 0.574±0.019 | **0.663** | 0.512 | 0.548±0.027 |
| Snyder (n=21) | 0.502±0.077 | 0.476±0.016 | 0.546±0.061 | 0.542±0.042 | 0.537±0.033 | 0.540±0.042 | n.a. | 0.588 | **0.595±0.113** |
| **Average** | 0.585±0.069 | 0.568±0.055 | 0.604±0.060 | 0.589±0.065 | 0.577±0.052 | 0.605±0.053 | 0.567±0.102 | 0.556±0.086 | **0.610±0.055** |

### Supplementary Methods

#### Method S1 Overview of signature scoring approaches: ssGSEA and geometric mean

We compared the COMPASS models (pre-trained COMPASS-PT and fine-tuned COMPASS-PFT as feature extractors from transcriptomic data) with two signature scoring methods: single-sample gene set enrichment analysis (ssGSEA) and geometric averaging (referred to as "Average"). For both ssGSEA and Average, we used the full set of genes underlying each concept to compute either a normalized ssGSEA score (GSEApy v1.1.3[18], TPM as input) or the mean of $\log_2$(TPM+1) values.

In contrast to these scoring methods, COMPASS introduces three key differences. First, its encoder is a gene language model that captures gene–gene interactions through attention, whereas ssGSEA and Average treat genes independently. Second, COMPASS implements a hierarchical mapping from genes to granular gene-set concepts and then to high-level immune concepts, rather than the flat architecture of ssGSEA and Average. Third, in COMPASS concept scores are pre-trained on TCGA (COMPASS-PT) and further fine-tuned on clinical cohorts (COMPASS-PFT), improving alignment with specific datasets and enhancing predictive performance (**Extended Data Fig. 2a**).

Because COMPASS-PFT requires labeled data, we split the 1,133-patient dataset into a 20% test set ($n = 226$) and an 80% fine-tuning set ($n = 907$). Using the fine-tuning set, we computed concept scores for `Cytotoxic T Cell` and `Exhausted T Cell` with ssGSEA, Average, COMPASS-PT, and COMPASS-PFT. We examined the relationship between these concepts by comparing Pearson correlations across methods (**Extended Data Fig. 2b**) and assessed score consistency using pairwise Pearson's $r$ for selected concepts. COMPASS concept scores were highly correlated with ssGSEA and Average (**Extended Data Fig. 2c**).

To evaluate the predictive utility of T cell concepts relative to signature-based scores, we assessed all four methods on the fine-tuning set ($n = 907$) and the independent test set ($n = 226$) for their ability to distinguish responders from non-responders using `Cytotoxic T Cell`, `Exhausted T Cell`, and the negative control `Reference`. Statistical significance was assessed using the Mann–Whitney U test comparing responders and non-responders (**Supplementary Figure S19**). We also computed the point-biserial correlation (PBC) between concept scores and binary response labels using SciPy (v1.10.1)[19], where higher PBC values indicate greater predictive power. Together, these analyses show that COMPASS-PFT refines concept representations to improve prediction while reducing non-informative signal.

#### Method S2 Benchmarking patient representations against signature scoring approaches

To assess the added value of the COMPASS encoder–projector, we benchmarked it against the fixed scoring baselines defined in Section **Method** S1 (geometric mean and ssGSEA), deriving patient-level representations directly from the same gene sets(**Extended Data Fig. 3**).

**Approaches compared.** Baselines included geometric mean aggregation of TPM values and ssGSEA signature scores computed for both granular and high-level concepts, as described in the previous section. For COMPASS, we used the pre-trained encoder-projector to embed expression profiles into biologically structured representations, again for both granular (132 dimensions) and high-level (43 dimensions) concepts.

**Classification model, evaluation scheme, and statistical analysis.** For all approaches, patient representations were used as input features to a logistic regression classifier (TPM-Average-LGR, TPM-ssGSEA-LGR, TPM-COMPASS-LGR). We performed a grid search over the inverse regularization parameter $C \in [10^{-4}, 10^{1}]$ with 10-fold stratified cross-validation, applying z-score normalization before model fitting and selecting the best hyperparameter by maximizing AUROC on the training folds. Model evaluation followed a leave-one-cohort-out protocol across 16 independent ICI-treated cohorts, holding out one cohort for testing in each round while training on the remainder. Performance was quantified by AUROC (**Supplementary Table S9**) and PR-AUC (**Supplementary Table S10**). Method performance was compared using paired Wilcoxon signed-rank tests across cohorts, with $p$-values adjusted for multiple testing where appropriate and significance denoted as * $p < 0.05$, ** $p < 0.01$, *** $p < 0.001$.

##### Method S3 Benchmarking against genome-wide transcriptomic methods

We benchmarked COMPASS against three transcriptomic-based machine learning approaches for ICI response prediction: **ENLIGHT**[20], **EaSIeR**[21], and **IRnet**[22]. We compared performance across three independent cohorts spanning distinct cancer types: **IMvigor210** (BLCA; anti–PD-L1), **Gide** (SKCM; anti–PD-1), and **Kim** (STAD; anti–PD-1). These datasets were selected because they cover different tumor types and provide both raw count and TPM gene expression data, enabling consistent evaluation across methods that require either input format. For retrainable models, including COMPASS and EaSIeR, we applied a **leave-one-cohort-out** protocol, training on the remaining 15 cohorts and testing on the held-out cohort to prevent data leakage. For inference-only methods such as ENLIGHT and IRnet, predictions were generated using the authors' official implementations or web services. Model performance was compared using **AUROC** and **AUPRC** across all cohorts.

**ENLIGHT.** ENLIGHT is a transcriptomics framework that infers target-centered genetic interaction signals to score therapy suitability and predict response across cancers[23–25], extending the SELECT approach[20]. Because ENLIGHT does not provide training code, we generated **ENLIGHT Matching Scores (EMS)** using the official web server (https://ems.pangeabiomed.com/), which supports predictions for drugs described in the original publication with user-supplied RNA-seq profiles. Since ENLIGHT does not include a PD-L1 model, we did not evaluate the **IMvigor210** cohort with this model. We therefore requested test access via https://ems.pangeabiomed.com/request-form and submitted the **Gide** and **Kim** cohorts for analysis. The resulting EMS values were used as continuous predictors and evaluated by AUROC and AUPRC against the corresponding clinical response labels.

**EaSIeR.** EaSIeR[21] is an RNA-seq–based tool for predicting biomarker-driven immunotherapy response. We installed the **EaSIeR R package** (https://github.com/olapuentesantana/easier_manuscript) and computed the five defined view scores (*ccpairs*, *immunecells*, *lrpairs*, *pathways*, and *tfs*) for all 16 cohorts (1133 patients). The detailed pipeline is documented here: https://github.com/mims-harvard/COMPASS/blob/main/paper/04_model_performance/08_ML_benchmark/EASIER/01_run_EASIER_per_cohort.r. Following the published approach, the ensemble EaSIeR score was first computed as the average of the five view-specific scores. To ensure a fair comparison with COMPASS, we additionally trained a **leave-one-cohort-out logistic regression (LGR) model** using the five view scores as input features. The model-based approach achieved higher predictive performance than the simple averaging strategy. Therefore, all final EaSIeR results are reported based on the LGR model.

**IRnet.** IRNet[22] is a graph neural network–based framework for immunotherapy response prediction that incorporates pathway-level biological knowledge to transform gene-level features into pathway-level features. We installed IRNet from the official repository (https://github.com/yuexujiang/IRnet) and generated predictions for the three benchmark cohorts using the **gene expression counts** as input by executing the official prediction script (`IRnet/predict.py`).

##### Method S4 Benchmarking against transformer-based methods for survival prediction

We evaluated the COMPASS response model for survival outcome prediction, using response probabilities as predictors, against two recently published transformer-based approaches: the **Clinical Transformer**[26] and the **cGAS–STING pathway (CSP) transformer model**[27]. The benchmarking included 860 patients across 11 independent cohorts with available overall survival data

**Cohorts and data used for survival prediction benchmarking.** For benchmarking, we employed a leave-one-cohort-out approach to evaluate model performance on unseen patient cohorts. The dataset comprised 16 cohorts with a total of 1,133 ICI-treated patients with recorded response outcomes. Among these, 860 patients from 11 cohorts had available overall survival data: *IMvigor210* ($n = 298$), *Rose* ($n = 89$), *Snyder* ($n = 21$), *Ravi-1* ($n = 98$), *Hugo* ($n = 25$), *Gide* ($n = 73$), *Liu* ($n = 107$), *Riaz* ($n = 51$), *Van Allen*

($n = 39$), *MGH* ($n = 34$), and *Ravi-2* ($n = 25$). In addition, 628 patients from nine cohorts had available tumor mutational burden (TMB) data: *IMvigor210* ($n = 234$), *Liu* ($n = 107$), *Rose* ($n = 88$), *Ravi-1* ($n = 51$), *Riaz* ($n = 46$), *Van Allen* ($n = 39$), *MGH* ($n = 28$), *Hugo* ($n = 25$), and *Ravi-2* ($n = 10$). For each benchmarking round, model performance was evaluated only on patients from the held-out cohort with available TMB and survival data, while training used all remaining cohorts, incorporating both survival and response labels but excluding any data from the test cohort.

**cGAS–STING pathway transformer model.** The cGAS–STING pathway (CSP) transformer model employs pathway-informed transcriptomic representations for survival prediction in hepatocellular carcinoma. We used the pre-trained model checkpoint and pathway file provided in the authors' repository (https://github.com/mlwalker123/CSP_survival_model/tree/main) to compute risk scores directly. As this baseline does not require fine-tuning, it serves as an external reference for comparison with transfer learning–based transformer models.

**Clinical Transformer model.** The Clinical Transformer code was obtained from https://codeocean.com/capsule/2256163/tree/v1. The standard Clinical Transformer uses 29 immune-related signature scores derived by ssGSEA[18] from functional gene sets[28]. We followed the hyperparameter settings reported by the authors for both pretraining and fine-tuning stages. To compare the Clinical Transformer and COMPASS, we evaluated three configurations (**Supplementary Figure S29**):

(i) **Clinical Transformer (ssGSEA-29)**, which used the 29 ssGSEA-derived immune-related signatures as input features;

(ii) **Clinical Transformer (ssGSEA-43)**, which used the 43 COMPASS-defined immune concepts as gene sets, with ssGSEA-derived signature scores provided as model inputs; and

(iii) **COMPASS + Clinical Transformer**, which used concept scores generated by the pretrained COMPASS encoder–projector as input features for the Clinical Transformer. In this configuration, both the COMPASS encoder–projector and the Clinical Transformer backbone were pretrained

The Clinical Transformer architecture followed the design described by Arango-Argoty et al.[26], and our implementation was adapted from their official repository. All three configurations were assessed under both transfer learning (with TCGA pretraining) and baseline (without pretraining) conditions.

**Evaluation setting: Transfer learning.** For all three configurations, we pretrained models on TCGA data (10,184 patients) using their respective input features for the Clinical Transformer. During self-supervised pretraining with a masked-feature prediction task, 10% of the data was held out as a validation set. The model architecture consisted of 8 transformer layers, 2 attention heads, and an embedding dimension of 128, trained for up to 2000 epochs following the Clinical Transformer protocol. We selected the 20,000-step pretrained checkpoint for transfer learning, as it achieved the lowest validation loss and was also recommended in the original publication (see **Supplementary Figure S30**).

In the transfer learning stage, pretrained weights were loaded, and the training set was split with 10% held out for validation across 10 independent runs. Models were trained for 300 epochs, and the optimal epoch (median across runs) was selected based on the average concordance index (C-index) on the validation sets. After determining the optimal pretrained checkpoint (20,000 steps) and training epoch, a final model was retrained using the full training data. For each test cohort, the best training epoch was optimized. Model evaluation followed a leave-one-cohort-out strategy, in which each of the 11 cohorts was held out in turn as the test set.

**Evaluation setting: Baseline setup.** For the baseline setup, we used the same hyperparameters as in the transfer learning experiments, except that no pretrained checkpoints were loaded for fine-tuning. We used the official Clinical Transformer implementation with survival-specific loss (approximated C-index loss) and evaluation metric (sigmoid C-index), adopting default parameters from the original study. Training was performed from scratch, with 10% of the training set reserved for validation, and repeated across 10 independent runs. Each model was trained for 300 epochs, and the optimal epoch (median across runs) was selected based on the average validation C-index. A final model was then retrained on the full training data for the selected epoch. The validation set performance (C-index) is summarized in **Supplementary Table S11**, and test performance under the leave-one-cohort-out evaluation across 11 cohorts is reported in **Supplementary Table S12**.

##### Method S5 Further ablations and sensitivity analyses of COMPASS

**Ablation of the cancer-type token.** To assess whether model performance is driven by cancer-type-specific response prevalence encoded in the cancer-type token, we performed explicit ablation analyses during fine-tuning. We compared models trained with and without the cancer-type input. In the ablated setting, the cancer-type token was removed, such that no cancer-type information was provided during training or inference, while all other inputs and model components were unchanged. In the full model, cancer type was included as an additional categorical input and jointly embedded with transcriptomic features.

To evaluate robustness under distribution shift, we conducted two out-of-distribution generalization experiments. First, in cross-indication generalization, we used a leave-one-indication-out strategy, excluding all cohorts of a given cancer type during training and evaluating exclusively on that held-out indication. Second, in cross-target generalization, we used a leave-one-target-out strategy, excluding all cohorts associated with a specific treatment target during training and evaluating on those cohorts.

Ablated and full models were trained under identical configurations, including optimizer, learning rate, batch size, and early stopping. Performance was evaluated using PR-AUC and ROC curves, with fixed random seeds for reproducibility. This design isolates the contribution of the cancer-type token and tests whether predictive performance arises from prevalence signals or transcriptomic features learned by the model. Results are shown in **Supplementary Figure S10**.

**Sensitivity analysis of data augmentation during pretraining.** We evaluated the effect of perturbation strength in the contrastive pretraining framework. Each training instance consists of an anchor sample, a perturbed positive sample, and a negative sample. Perturbations were applied only to the positive sample, while the anchor remained unchanged.

We considered two perturbation mechanisms: feature masking and additive Gaussian noise. For masking, we varied the masking probability (`mask_p_prob`) from 0.1 to 0.9. For jitter, we varied the standard deviation of Gaussian noise (`jitter_p_std`) from 0.1 to 0.9. In each experiment, only one perturbation type was active, with the other set to zero.

Pretraining was performed on TCGA transcriptomic data using a subset of 2,475 genes (including all concept genes) to reduce computational cost. Data were split using a 90/10 stratified split by cancer type. All experiments were conducted using immuno-compass v2.0.5, with model architecture, optimizer settings, and training schedules held constant. Random seeds were fixed.

We evaluated (i) self-supervised loss during pretraining on training and held-out data, and (ii) downstream performance measured by PR-AUC. Downstream evaluation used the ITRP dataset under a leave-one-cohort-out (LOCO) protocol across 10 medium- and large-sized cohorts ($n > 30$). For each pretrained model, we evaluated both no fine-tuning (NFT) and post fine-tuning (PFT), using identical hyperparameters. Results are shown in **Supplementary Figure S36**.

**Sensitivity analysis of hard-negative sampling.** To examine the role of negative sample hardness in contrastive pretraining, we introduced a parameter $K$ controlling the size of the neighborhood used for negative sampling. Negative samples were selected using $k$-nearest neighbors (KNN) in gene expression space. The parameter $K \in [0.1, 1]$ defines the proportion of the dataset used as the candidate pool.

At $K = 0.1$, negatives are drawn from the closest 10% of samples (transcriptionally similar, hard negatives), whereas $K = 1$ allows sampling from all other samples (including distant, easy negatives). Smaller $K$ enforces local, hard negatives, while larger $K$ progressively introduces more distant samples.

We varied $K$ from 0.1 to 1.0 while holding all other pretraining parameters fixed. Pretraining used the same TCGA data, train–test split, and model configuration as above, with fixed random seeds. Performance was evaluated using (i) pretraining loss and (ii) downstream PR-AUC under both NFT and PFT settings. Downstream evaluation was performed on the ITRP dataset using the LOCO framework.

This design isolates the effect of negative sampling locality on representation learning. Results are shown in **Supplementary Figure S37**.
